# Analysing complex interventions using component network meta-analysis

**DOI:** 10.1101/2025.06.30.25330567

**Authors:** Areti Angeliki Veroniki, Dianna Wolfe, Brian Hutton, Guido Schwarzer, Daniel I McIsaac, Sharon E. Straus, Dan Jackson, Andrea C. Tricco

**Author notes:** **Corresponding Author** Dr. Areti Angeliki Veroniki, PhD, MSc Scientist, Knowledge Translation Program Assistant Professor, Institute for Health Policy, Management, and Evaluation, University of Toronto Li Ka Shing Knowledge Institute of St. Michael’s Hospital, Unity Health Toronto 209 Victoria Street, East Building, Toronto, Ontario, M5B 1T8, Canada.

## Abstract

Systematic reviews with network meta-analysis (NMA) frequently evaluate complex interventions combining multiple healthcare interventions (known as components). Components may act separately of each other or in conjunction with other components, synergistically or antagonistically. Component effect estimation is crucial to produce relevant and clinically meaningful evidence. However, standard NMA cannot quantify individual component effects of complex interventions. This study presents methods for modeling complex interventions and highlights the advantages and limitations of component NMA (CNMA). CNMA enables the estimation of individual component effects, whether additive or interactive. Interaction CNMA can be considered an extension of the additive CNMA model that includes interaction terms. We give practical guidance on how to carry out these analyses via empirical examples, which showcase both the strengths and limitations of CNMA. Implementing CNMA models is complex and requires the skills of a multidisciplinary team including clinicians, methodologists, and statisticians.

**Summary points:**

1. CNMA provides the opportunity to disentangle the effects of components of complex interventions and assess their efficacy or safety, accounting for potential interactions in component combinations.
2. Under the additivity assumption, CNMA assumes that the total effect of a complex intervention is the sum of its individual component effects (e.g., if component A lowers a symptom score by 2 points and B by 1 point, A+B is expected to lower it by 3 points), while interaction CNMA is used when there is evidence of violation of additivity and the combined effect differs due to synergy or antagonism.
3. Interaction CNMA can be considered a compromise between additive CNMA and standard NMA, but selecting interaction terms requires clinical, statistical, and methodological considerations.
4. Clinicians and other knowledge users should be engaged in the selection of interaction CNMA models to ensure biological plausibility.

## Introduction

Systematic reviews frequently evaluate complex interventions that combine multiple healthcare interventions—called components—to achieve important patient and health system outcomes.1, 2 Clinicians and patients may want to know the safest and most effective individual component and combination of components amongst many identifiable components and combinations to treat or prevent a given condition. For example, non-pharmacological interventions, such as fall prevention interventions,3 usually share common components that relate to the nature of the intervention (e.g., fall prevention devices, social engagement, cognitive behavioral therapy), the provider (e.g., clinician, nurse, layperson), the intensity (e.g., daily, weekly), setting (e.g., hospital, home, community) and the mode of delivery (e.g., virtually, in-person). Quantitative methods—such as component network meta-analysis (CNMA), an extension of standard network meta-analysis (NMA)—can compare complex interventions and their separate component effects in a single model.1, 2 Unless some combinations of interventions are treated as having the same efficacy, sometimes referred to lumping interventions (see section When is a component network meta-analysis useful?), a standard NMA estimates the effects of entire interventions (e.g., the fall prevention intervention combining calcium and vitamin D vs usual care). However, a CNMA estimates the individual effects of each component (e.g., calcium alone vs usual care and vitamin D alone vs usual care). This component-level insight supports more informed clinical decision-making and guideline development because it enables clinicians and policymakers to identify which components drive the effectiveness of interventions, alone or synergistically, and which combinations may be redundant or antagonistic.3

Tsokani et al.2 recently provided a description of the advantages and limitations of CNMA compared to standard NMA. In this paper, we present approaches to modelling complex interventions using real-life examples; elaborate on when a CNMA is needed; emphasize the importance, advantages, and limitations of CNMA; give practical guidance on how to carry out these analyses; and provide readily available R code for data modeling and visualization.

### Motivating examples

For our primary motivating example, we used data from 106 randomized controlled trials (RCTs) of 8,816 adults undergoing surgery that assessed prehabilitation interventions for the prevention of post-operative complications.4 In these RCTs, individuals were allocated to receive a prehabilitation intervention comprised of one or more of the following components: exercise (EXE), nutrition (NUT), cognitive (COG), or psychosocial (PSY); an active comparator intervention; or usual care (UC) (an inactive comparator; reference group). The outcome of interest was binary, defined as the occurrence of any post-operative medical or surgical complication during the initial hospital stay or within 30 days following surgery. As this was a negative outcome, odds ratios less than one indicated a beneficial effect of the intervention. Seven active interventions were compared to UC and formed a network of trials with one loop due to a single three-arm study (Figure 1b). In addition to the four single-component interventions (EXE, NUT, COG, PSY), three complex interventions were assessed in the eligible RCTs: EXE + NUT, EXE + PSY, EXE + NUT + PSY. Data are presented in Appendix1. The evidence graph and selection of components were informed by data available in the literature and input from the research team. Team-wide questionnaires were used to identify key prehabilitation components and prioritize critical outcomes for analysis, while a broader taxonomy was pre-specified at the protocol stage.^4, 5^

In the main text, we illustrate key CNMA concepts using the prehabilitation intervention example.^4^ Two additional examples, presented in Appendices 1-9, demonstrate real-world complexities of applying CNMA to complex interventions: one focusing on the prevention of fall-related fractures^3^ and the other on improving quality of life through knowledge translation strategies.^6^ The nature of these two additional examples is quite different, highlighting both the strengths and limitations of CNMA. The example of fall prevention interventions^3^ shows where CNMA is particularly helpful in a simplified analysis that performs well. In contrast, the example of knowledge translation interventions^6^ suggests that CNMA is less likely to offer improvements over NMA, given the complex interactions between components. In this case, higher-order interactions seem necessary, bringing the CNMA model closer to the standard NMA. While CNMA can streamline some analyses, it may struggle to outperform NMA in more complex scenarios.

### What is a component network meta-analysis?

The complexity of an intervention lies in its included components, which may act independently of or in conjunction with each other. Components reflecting the nature of the intervention may interact with each other or with other intervention characteristics, such as behaviours of those delivering (e.g., clinician, nurse, layperson) or receiving the intervention, groups targeted by or delivering the intervention, and the intensity (e.g., daily, weekly), setting (e.g., hospital, home, community), and mode of delivery (e.g., virtually, in-person) of the intervention.^1^ In our prehabilitation example, consider an RCT assessing the efficacy of a complex intervention consisting of two components, *EXE* and *NUT*, compared with *UC* in preventing post-operative complications. Although we can estimate the effect size of this complex intervention by considering it as a single treatment, a question of perhaps greater clinical interest may pertain to the relative merits of the individual components (*EXE* and *NUT*). A potential design to answer this question would be a factorial RCT, wherein participants are randomized to receive one of four potential options: *EXE, NUT*, both *EXE* and *NUT* (conventionally labeled “*EXE* + *NUT*”) or neither *EXE* nor *NUT*, i.e., *UC.* Unfortunately, this design is not commonly encountered due to the increased resources required, reducing feasibility.^7^

If multiple RCTs exist, we typically conduct a standard NMA to evaluate the relative efficacy of the interventions of interest.^8, 9^ However, standard NMA cannot quantify the effects of individual components from complex interventions. Consider a scenario in which an NMA of interventions including *EXE, NUT*, and *COG* for prevention of post-operative complications suggests *EXE* has a large effect, *EXE + NUT* has a small effect, and *EXE + NUT + COG* has a large effect, where the same components (*EXE* and *NUT*) are included in the most and least efficacious interventions. This increases the difficulty of relating components to efficacy. To develop tailored recommendations to optimise patient care and better allocate finite health care resources, it is vital to disentangle the effects of each component of the prehabilitation interventions using CNMA.^2^ CNMA can estimate a separate effect for each individual component of complex interventions.^10^ For example, the efficacy of the *EXE* and *NUT* components can be estimated separately and in combination, without requiring direct evidence from previous RCTs by borrowing information from trials sharing each of the components.

### When is a component network meta-analysis useful?

CNMA allows us to derive estimates of the effects of individual components, even when delivered in combination. Exploring intervention components in a network of trials can help with exploring clinical heterogeneity, b) gaining insights into how an intervention works and the drivers of its effect, c) identifying effective components, and d) informing the design of future studies, particularly regarding what components and their combinations may be most promising to assess.

A key question is: *how should we analyse complex interventions?* This depends on the research question. If the purpose of the systematic review is to address whether the underlying class of interventions works as a whole (e.g., ‘*do prehabilitation interventions as a whole work compared to usual care?*’), then lumping all interventions into a single group and performing analyses using a single-effect model is appropriate (i.e., a pairwise meta-analysis; **Figure 1a**).^1, 11, 12^ However, this model provides limited information for clinical practice or implementation decisions, since different interventions (and their underlying components) are ignored using this approach. A single effect model assumes that intervention (and component) effects are equivalent, and a single effect at the intervention class level is estimated. Lumping all interventions (and their components) in a single group may also increase or mask heterogeneity (e.g., combining strong and weak intervention effects can increase heterogeneity and hide valuable information).

**Figure 1.**
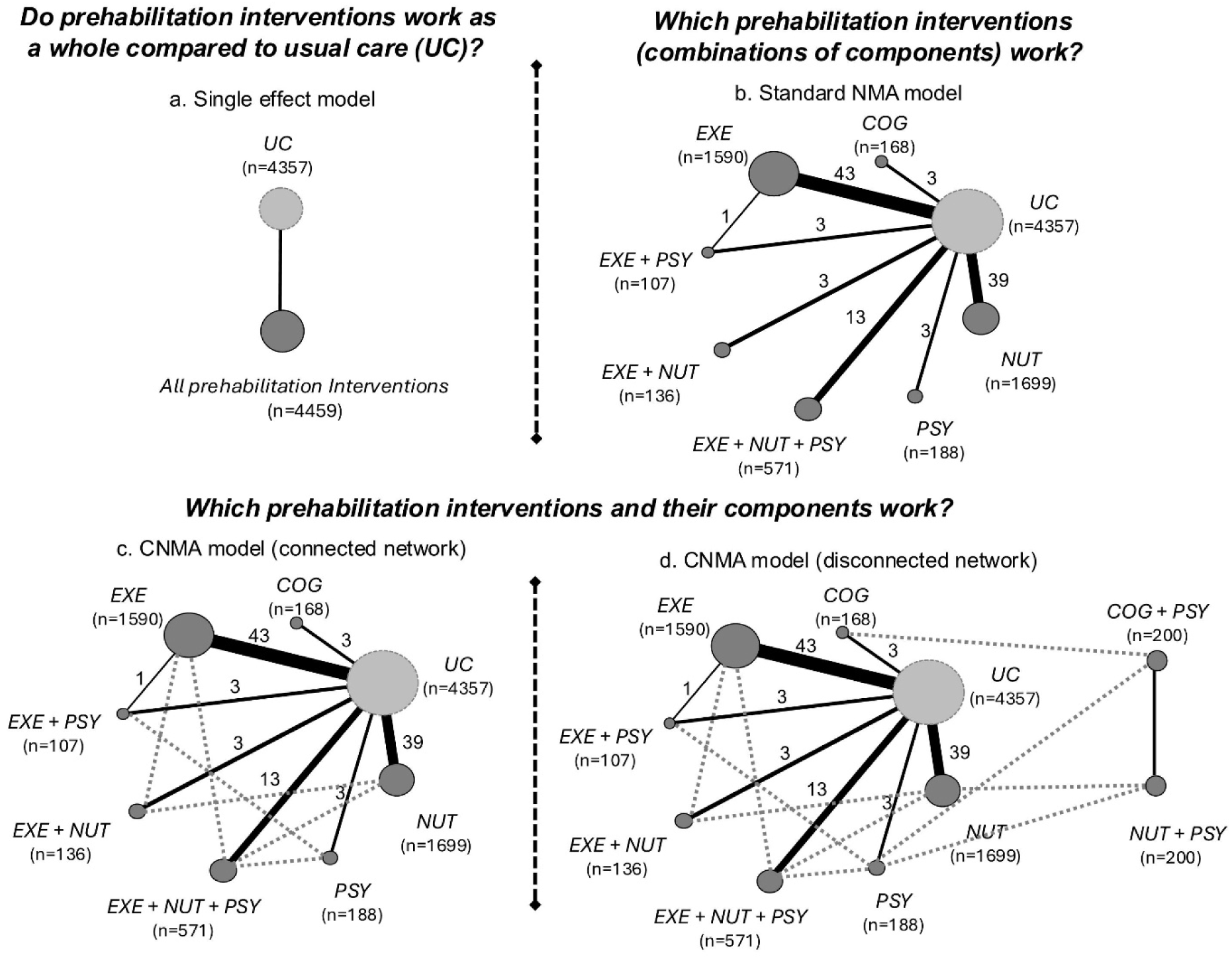
Network diagrams depicting different research questions on the prevention of post-operative complications. Nodes represent different combinations of components evaluated within each model and edges depicting studies comparing the different combinations of components they connect. Thickness of solid lines is proportional to the number of studies included in the group comparison, and node size is proportional to the number of patients included in the underlying group. Dashed lines represent joints added by component network meta-analysis model between different combinations of components sharing common components.

If the purpose of the systematic review is to investigate which types of interventions are most effective and how they compare to each other (e.g., ‘*which prehabilitation interventions [combinations of components] work?*’), then considering each unique combination of components as a separate node within the evidence network and performing a standard NMA might be more appropriate (**Figure 1b**).^12–15^ Applying standard NMA methods considers each unique combination of components as a separate intervention and thus estimates an intervention-level effect for each unique combination of components.^16, 17^ This is the standard analysis of a network of trials provided by most meta-analytical software (e.g., **netmeta** package in R^18^). Separating all unique combinations of components into distinct nodes can reduce heterogeneity in the network when compared to the single-effect model (i.e., pairwise meta-analysis) described above.^19, 20^ In standard NMA, unique combinations of components exhibit sufficiently noteworthy dissimilarities to justify separate categorization. However, standard NMA ignores any potential relationships between combinations of components (e.g., *EXE + NUT*) and the individual components (e.g., *EXE, NUT*) that are part of the combination. The individual components are considered unrelated; however, component effects may be additive or different sets of components may interact with each other to decrease or increase the effect. The standard NMA model cannot separate the effects of the different components included in a network of trials and cannot estimate their contribution to the network effects. Also, certain components may be effective in some combinations but not in others, which increases the difficulty in making inferences about component effects. When multiple components are assessed in unique combinations in the eligible trials, standard NMA may have a sparse network with limited data points per intervention comparison, an increased number of parameters in the model, and decreased statistical power. In contrast, CNMA can be an improvement over standard NMA in such situations by borrowing strength across studies with common components, reducing the number of parameters estimated, and enabling the estimation of component-specific effects— thereby increasing interpretability.

If the purpose of the systematic review is to address which individual components are more effective (e.g., ‘*which components of prehabilitation interventions work?*’) then CNMA is appropriate (**Figure 1c**).^11, 17, 21^ CNMA models are a generalization of NMA models, where complex interventions can be analysed by assuming *additive* or *interactive* components. Additivity in CNMA assumes that the effect of a complex intervention is the sum of the effects of its components (i.e., a complex intervention including *EXE* and *NUT* [i.e., *EXE + NUT*] has an intervention effect equal to the sum of the individual component effects of *EXE* and *NUT*). Additionally, CNMA can consider interactions to allow synergistic (positive interactions) or antagonistic (negative interactions) effects between components. The use of interactions allows us to relax the *additivity* assumption. A CNMA model with all possible interactions (i.e., all unique combinations of components with direct evidence), called the full interaction model, is equivalent to the standard NMA.^17^ These models are discussed further in the next section. CNMA allows us to explain heterogeneity by exploring differences in the effects of the intervention characteristics (e.g., nature, provider, intensity, setting, and mode of delivery of intervention). In standard NMA, intervention effects are derived only from the studies that evaluated the specific combinations of components. However, in CNMA, the component effects are derived from all studies that evaluated interventions that contain the components of the CNMA. This borrowing of information effectively increases the power to detect differences between components, and hence their combined effects (i.e., the intervention effects).^17^

CNMA models can also be performed to reconstruct disconnected networks,^22^ as long as complex interventions in each network share common individual components and there is strong clinical evidence of additivity. For example, if prior studies or expert consensus suggest that the effects of individual components (e.g., *EXE* and *NUT* in prehabilitation) are independent and expected to combine without interaction. Strong clinical evidence for additivity must exist because the additivity assumption cannot be assessed statistically in disconnected networks.^17^ Additivity can be assessed by comparison of the CNMA with the standard NMA, if the network is connected. However, the standard NMA cannot be performed on a disconnected network.^22^ As an example, let us consider a network of 10 unique combinations of components (including *UC*) presented in **Figure 1d**. This network includes 107 two-arm studies comparing combinations of components with *UC,* one two-arm study comparing two active combinations of components (*EXE* vs *EXE + PSY*), and one two-arm study comparing two unique combinations of components (*COG + PSY* vs *NUT + PSY*). The two-arm study *COG + PSY* vs *NUT + PSY* is not connected to the remaining network of trials, and hence two disconnected networks are formed. CNMA models can reconnect the disconnected networks, *as long as there are common components across the two networks*. Since the components *COG*, *PSY*, and *NUT* are common within the two networks, their efficacy can be estimated using the CNMA model (**Figure 1d**).

### Should we choose additivity or interaction component network meta-analysis?

Depending on how the effects of individual components are related to each other (e.g., additive or interactive), CNMA may be categorized as a) an additive CNMA or b) an interaction CNMA. Interaction CNMA can be considered an extension of the additive CNMA model that includes interaction terms. These models have been described both in Bayesian^11^ and frequentist settings.^17^

### Additive component network meta-analysis model

The additive CNMA model assumes that there is a separate and independent effect for each of the components of a complex intervention. Under the additivity assumption, the total intervention effect is the sum of the relevant component effects. That is, the effect of the combination of components *EXE* + *NUT* (including two components *EXE* and *NUT*) against *UC* is equal to the sum of the individual component effects of *EXE* versus *UC* plus the individual component effects of *NUT* versus *UC.*^11, 13, 15, 21, 23^ If the effect estimate is a log odds ratio (LOR), then LOR_EXE + NUT VS. UC_ = LOR_EXE VS. UC_ + LOR_NUT VS. UC_. This approach can readily be extended to combinations of components consisting of more than two components.

While this model can theoretically predict the effects of component combinations not evaluated in the included trials, such predictions should be viewed as exploratory and interpreted with caution.^17^

Under the additivity assumption, the comparative effects of common components in pairwise comparisons against the same component (e.g., *NUT*) cancel out.^12, 17^ Therefore, the effect of *EXE + NUT* vs. *NUT* is identical to the incremental effect of *EXE* alone (or simply the effect of *EXE* vs. *UC*). If desirable, it is possible to define a single component as *inactive* which means that adding it to any combination of active components does not affect the underlying intervention effect. For example, assuming that *UC* is an inactive component, the effect of *EXE + NUT* vs. *UC* is identical to the incremental effect of *EXE + NUT* because the effect of *UC* is assumed to be zero. If *UC* is considered an inactive component, intervention effects will be relative to *UC*, which is assumed to have no impact when combined with other interventions. However, if no inactive component (e.g., *UC*, control, placebo) is chosen or available in the RCTs, incremental intervention effects will be relative to no intervention. The choice of an inactive component is important as it impacts the parameterization of component effects, and it is assumed that the comparison of an inactive vs. an active component leads to the active component’s net effect.^17^ Whether an inactive component is considered in a CNMA is at the discretion of the review authors and should be guided by clinical expertise.

### Interaction component network meta-analysis model

Additive CNMA assumes that there is no interaction between components. However, clinically, components may interact with each other (i.e., the effect of a component may differ in the presence of another component), and additive CNMA may lead to biased estimates of component effects. If the additive CNMA does not fit the data well (i.e., the additivity assumption is not met), then relaxing the additivity assumption and assuming interactions between components in an interaction CNMA model may be appropriate to explore which sets of component effects are synergistic or antagonistic.

The interaction CNMA model is an extension of the additive CNMA model that includes additional terms for component interactions that may account for higher (synergistic) or lower (antagonistic) intervention effects than the sum of the component effects alone.^11–13, 23^ A two-way interaction CNMA model allows pairs of components within interventions to interact, resulting in potentially lower or higher component effects compared to the additive effects of the two components. In our illustrative example, a two-way interaction model answers the question: ‘*Are prehabilitation interventions containing specific pairs of components violating the additivity assumption?*’ For example, the effect of the combination of components *EXE + NUT*, where components *EXE* and *NUT* have an interaction effect, is: ^LOR^_EXE + NUT VS. UC_ ^= LOR^_EXE VS. UC_ ^+ LOR^_NUT VS. UC_ ^+ LOR^_EXE*NUT VS. UC_^. In other words,^ when exercise is implemented along with nutrition to prevent post-operative complications, their combined effect is amplified (or suppressed) by a specified amount (i.e., the effect of the interaction term). Overall, there are three possible situations regarding the relationship between *EXE* and *NUT* components for prevention of post-operative complications:

1. there is minimal or no interaction between *EXE* and *NUT,* leading to LOR_EXE*NUT_ being approximately zero (or OR_EXE*NUT_=1);
2. the interaction is positive (synergistic) between *EXE* and *NUT,* where LOR_EXE*NUT_ is negative (or OR_EXE*NUT_<1), suggesting the log-odds of post-operative complications are reduced, and hence *EXE* and *NUT* together are more protective than expected from their individual effects; or
3. the interaction is negative (antagonistic) between *EXE* and *NUT,* where LOR_EXE*NUT_ is positive (or OR_EXE*NUT_>1), suggesting the log-odds of post-operative complications are increased, and hence the combined effect is less protective than expected.

The direction of effect of the interaction would be the opposite for beneficial outcomes (e.g., quality of life). Specifically, LOR_EXE*NUT_ < 0 indicates a negative interaction, meaning *EXE* and *NUT* together are less beneficial than expected; e.g., their combined use reduces improvements in quality of life. However, LOR_EXE*NUT_ > 0 indicates a positive interaction, with *EXE* and *NUT* combination enhancing benefits more than either component alone.

In principle, the interaction CNMA model can incorporate interaction terms for each unique combination of components found in the dataset and, hence, can include interactions between two or more components. As explained above, including all estimable interactions of all orders found in the dataset would lead to the standard NMA model (i.e., the full-interaction model).^17^ However, there is a trade-off between model fit and estimation precision; as more interaction terms are added to the model, the number of parameters to be estimated increases, decreasing precision and eventually statistical power. Ideally, interaction terms should be defined a priori based on their clinical plausibility as determined by clinical expertise and the literature; an entirely data driven approach is discouraged (i.e., testing all possible interaction models). In a frequentist setting, adding interaction terms for component combinations that are missing from the dataset is not possible, since these are not estimable (i.e., the design matrix on the active components cannot be defined for a non-existent combination of components).^10^ These interactions cannot be validated since there is no direct evidence for examination.^17^ In a Bayesian setting, this could, in principle, be achieved by incorporating informative priors. However, careful consideration would be needed when selecting these priors, as they can be expected to strongly influence the posterior distribution, provided that there is no direct evidence from the data. The power to estimate and detect evidence of interactions will be limited by the availability of data on the unique combinations of components.

In the following, we focus on the inclusion of identifiable interaction terms, applicable to both frequentist and Bayesian settings. Overall, interaction CNMA is an extension of additive CNMA and a compromise between additive CNMA and standard NMA.

## How do we fit a component network meta-analysis?

### Model selection

Initially, an additive CNMA model is fitted and assessed to determine whether the additivity assumption is met. The additivity assumption can be assessed by comparing the effects of different combinations of component pairs: for example, does it appear that the effect of *EXE* vs. *NUT* equals the effect of *EXE + COG* vs. *NUT + COG* and the effect of *EXE + COG + PSY* vs. *NUT + COG + PSY*? A statistical test to assess the additivity assumption, proposed by Rücker et al,^17^ is based on comparing the Q heterogeneity statistics from the NMA and CNMA models. The difference between the Q statistic for the additive CNMA model (Q_Add_, which has fewer parameters) and the Q for the NMA model (Q_NMA_, which has more parameters) can be used to inform which model fits the data better^24^. This approach is analogous to a test of subgroup differences in pairwise meta-analysis; for instance, comparing the effect of *EXE* vs. *NUT* across three subgroups: a) no additional therapy, b) *COG* added, and c) *COG + PSY* added. A statistically significant difference in Q suggests that the additivity assumption is not met, and further model exploration with interaction terms should be considered. This test can only be applied to connected networks^17, 22^ (e.g., **Figure 1c**). When the additivity assumption is met, compared to standard NMA, the additive CNMA model can increase precision of estimated component and intervention effects, while also yielding results that are expected to be unbiased.^13^

When the additivity assumption is not met, suggesting the need for an interaction CNMA model, model selection can be guided by changes in the Q statistic in a frequentist setting or deviance information criterion in a Bayesian setting. In a frequentist setting, similar to the test of the additivity assumption, the difference between the Q statistics (i.e., Q for additive CNMA [Q_Add_] and the Q for the interaction CNMA [Q_Int_]) can be used to explore if adding *single* interaction terms improves the fit over the additive model. This corresponds to a likelihood ratio test and is related to the Akaike Information Criterion (AIC),^22^ which can be used for model selection. A significant improvement in model fit over the additive CNMA occurs when the Q statistic of the interaction CNMA is statistically significantly lower than that of the additive CNMA, using an adjusted p-value according to the difference in degrees of freedom of the two models (e.g., p < 0.157 for one degree of freedom)^24, 25^ to account for the lower power of the test to detect violations of additivity. However, this test is not well powered to identify minor violations of the additivity assumption.^22^ Similar to the Q test for heterogeneity, its statistical power is expected to be low in sparse networks with few studies across intervention comparisons.^26^ Heterogeneity should also be evaluated across models, a reduction of which may suggest that the interaction model explains some of the variability observed across studies.

The Q statistics of CNMA models with two or more interaction terms (called *multi-interaction* models) can also be compared to those of nested interaction CNMA models with fewer interaction terms (i.e., lower order). However, the process of determining the ‘best fitting’ model should be pre-specified and clearly explained and justified. For example, one approach could be to apply models with all *a priori* determined and clinically relevant two-way interaction terms, then all models with all *a priori* determined and clinically relevant three-way interaction terms, and subsequently combine the best fitting models to create more complex ones. Overall, there are various strategies, both in terms of the models considered and the statistical metrics (e.g., Q statistics, p-values, heterogeneity, deviance information criteria^27^) used to assess model fit, which can lead to different results. In our illustrative and empirical examples, we focused on the Q statistics and corresponding p-values. However, this is just one approach and others are possible. For example, in cases where notable differences in heterogeneity are observed across models, this might instead be used as a model selection criterion. Overall, these statistical approaches should be used in combination with clinical expertise regarding how the components are likely to work together. Different approaches and component taxonomies may yield different results, as they address distinct questions. As such, findings may not be generalizable across all populations or settings.

### Application to the prehabilitation interventions example

We demonstrate a model selection approach using the network of trials on prevention of post-operative complications (**Figure 1c**). For this network, there was no strong evidence that the additivity assumption was violated (Q_diff_ = 3.52, *df* = 3; p-value = 0.318; **Table 1**); however, for demonstration purposes, we will explore all interaction models. Interactions to be explored in statistical models should be selected using clinical judgement from the possible interaction terms identifiable in the available data. In particular, the final selection of interactions should be guided by clinical judgment, where clinicians assess their clinical plausibility *a priori* for inclusion in the models.

**Table 1.**
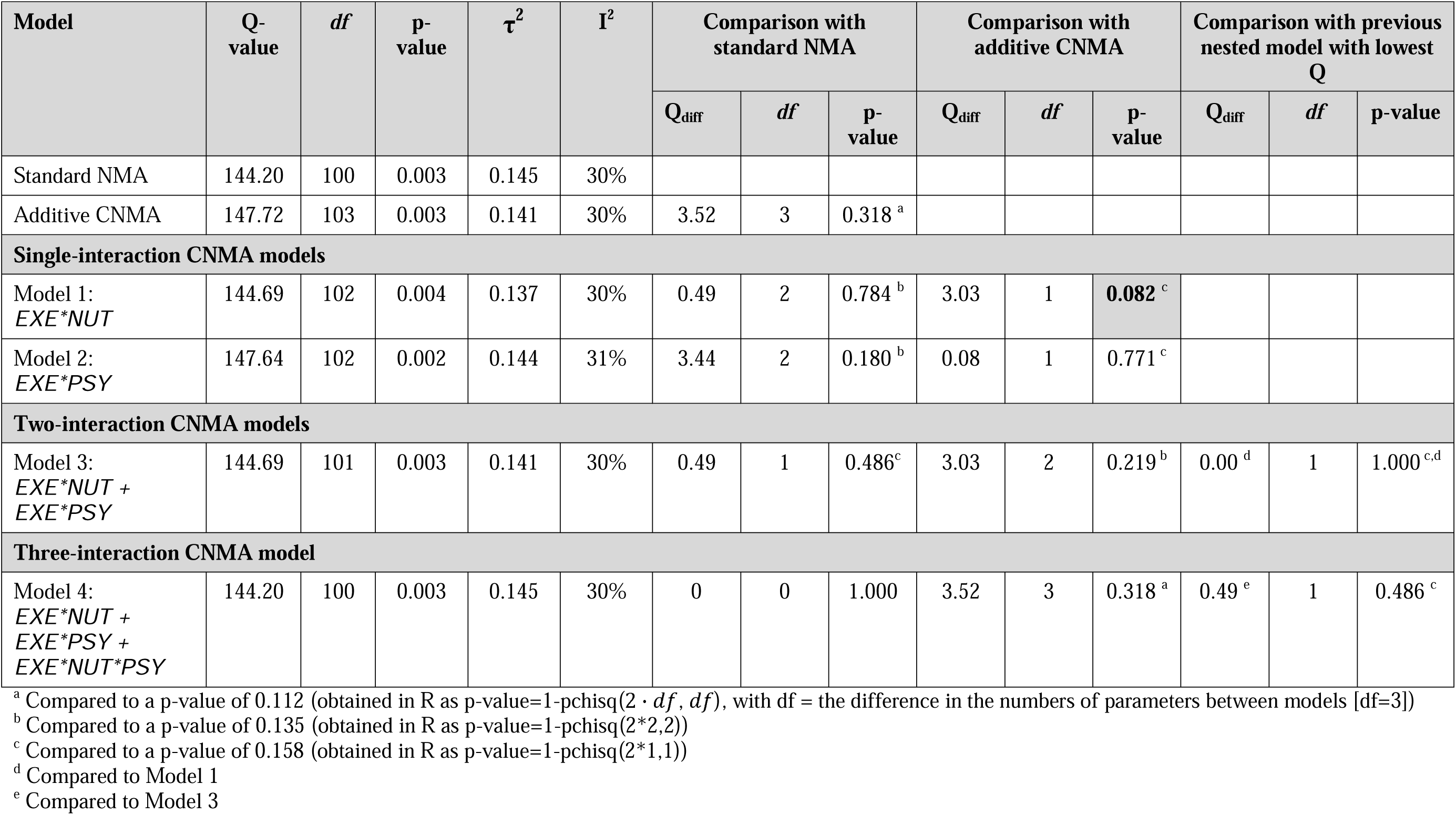
Model selection for a dataset with combinations of components *EXE, NUT, PSY, COG, EXE + NUT, EXE + PSY, EXE + NUT + PSY*, and *UC*. Statistically significant differences have been highlighted in bold text. Model 4 can be seen to be equivalent to the standard (i.e., full interaction) NMA.

| Model | Q-value | df | p-value | $\tau^2$ | I<sup>2</sup> | Comparison with standard NMA | | | Comparison with additive CNMA | | | Comparison with previous nested model with lowest Q | | |
| --- | --- | --- | --- | --- | --- | --- | --- | --- | --- | --- | --- | --- | --- | --- |
|  |  |  |  |  |  | Q<sub>diff</sub> | df | p-value | Q<sub>diff</sub> | df | p-value | Q<sub>diff</sub> | df | p-value |
| Standard NMA | 144.20 | 100 | 0.003 | 0.145 | 30% |  |  |  |  |  |  |  |  |  |
| Additive CNMA | 147.72 | 103 | 0.003 | 0.141 | 30% | 3.52 | 3 | 0.318<sup>a</sup> |  |  |  |  |  |  |
| **Single-interaction CNMA models** |  |  |  |  |  |  |  |  |  |  |  |  |  |  |
| Model 1:<br>*EXE\*NUT* | 144.69 | 102 | 0.004 | 0.137 | 30% | 0.49 | 2 | 0.784<sup>b</sup> | 3.03 | 1 | **0.082<sup>c</sup>** |  |  |  |
| Model 2:<br>*EXE\*PSY* | 147.64 | 102 | 0.002 | 0.144 | 31% | 3.44 | 2 | 0.180<sup>b</sup> | 0.08 | 1 | 0.771<sup>c</sup> |  |  |  |
| **Two-interaction CNMA models** |  |  |  |  |  |  |  |  |  |  |  |  |  |  |
| Model 3:<br>*EXE\*NUT + EXE\*PSY* | 144.69 | 101 | 0.003 | 0.141 | 30% | 0.49 | 1 | 0.486<sup>c</sup> | 3.03 | 2 | 0.219<sup>b</sup> | 0.00<sup>d</sup> | 1 | 1.000<sup>c,d</sup> |
| **Three-interaction CNMA model** |  |  |  |  |  |  |  |  |  |  |  |  |  |  |
| Model 4:<br>*EXE\*NUT + EXE\*PSY + EXE\*NUT\*PSY* | 144.20 | 100 | 0.003 | 0.145 | 30% | 0 | 0 | 1.000 | 3.52 | 3 | 0.318<sup>a</sup> | 0.49<sup>e</sup> | 1 | 0.486<sup>c</sup> |

<sup>a</sup> Compared to a p-value of 0.112 (obtained in R as p-value=1-pchisq(2 · df, df), with df = the difference in the numbers of parameters between models [df=3])
<sup>b</sup> Compared to a p-value of 0.135 (obtained in R as p-value=1-pchisq(2\*2,2))
<sup>c</sup> Compared to a p-value of 0.158 (obtained in R as p-value=1-pchisq(2\*1,1))
<sup>d</sup> Compared to Model 1
<sup>e</sup> Compared to Model 3

In this small network, there are two estimable two-way interactions — *EXE*NUT* and *EXE*PSY* — and one three-way interaction, *EXE*NUT*PSY*, and all were considered clinically meaningful. In more complex networks, developing interaction models for all identifiable interactions is not advisable. With entirely data driven approaches that make no use of clinical judgement, multiple testing increases the likelihood of finding a model that appears to perform well by chance alone, given the large number of comparisons with the additive model, the standard NMA model, and with other nested interaction models.

To determine if an interaction CNMA model improves upon all previously fitted models, its Q statistic is compared to that of 1) the standard NMA, 2) the additive CNMA, and 3) a previous nested ‘best-fitting’ interaction CNMA model, for multi-interaction models. The most parsimonious model with a Q statistic not significantly different from that of the standard NMA - was preferred in our example. A forward-selection model building approach was used, with all clinically relevant interactions individually added to the additive model, creating multiple separate models with one interaction term each (**Table 1**, Models 1–2). Note that (1) a single-interaction *NUT*PSY* model is missing because no studies evaluated *NUT + PSY*, and (2) a model with the three-way interaction term alone (*EXE*NUT*PSY*) was not generated because the identifiable two lower-order interaction terms *EXE*NUT* and *EXE*PSY* should also be added to the model; in this small network, this was equivalent to the full-interaction (standard) NMA (Model 4). Without including lower-order interactions, if the *EXE*NUT*PSY* interaction was found to have a beneficial or adverse effect, we would not know if the effect was driven by just two of the three components. Identifiable lower-order interactions allow us to disentangle the effects and understand the contributions of each component. Also, the financial cost of performing all three components, rather than just two, is an important factor to consider. Therefore, when higher-order interactions are included in the model, our position is that it is good practice to also incorporate their corresponding lower-order interactions, whenever possible.

Although the additivity assumption held, for demonstration purposes interaction models were fit. When compared to the additive CNMA model, the single-interaction model *EXE*NUT* had a better fit (**Table 1**, Model 1: *df* = 1, p-value = 0.082 < 0.158). In light of the additivity assumption being met, caution should be made in interpreting this finding as multiple testing may spuriously identify statistically ‘better’ models. In the context of this example, the effect estimate of the interaction term was found to be opposite to currently held clinical expertise (i.e., the *EXE* and *NUT* components were found in modelling to be antagonistic, while clinically there is strong belief that the effects of exercise and nutrition are either additive or synergistic).^4^ Thus, given that the additivity assumption held, the significant finding for interaction Model 1 was deemed to be spurious. Model-fit testing using the Q statistic demonstrated that neither single-interaction model fit better than the standard NMA model (**Table 1**, Model 1: *df* = 2, p-value = 0.784 > 0.135; Model 2: *df* = 2, p-value = 0.180 > 0.135).

Again, for demonstration purposes, we also fit all possible models with two identifiable interaction terms, of which there was one model (Model 3), and we compared its model fit against the standard NMA, the additive CNMA, and Model 2 (nested single-interaction CNMA model with the lowest Q statistic). No gains in model fit were observed. For completeness, the full-interaction model has been reported (Model 4), demonstrating its equivalence to the standard NMA.

## How do we examine transitivity, consistency, and homogeneity in component network meta-analysis?

### Additional model assumptions

The fundamental NMA assumptions of transitivity and consistency (both corresponding to the assumption of exchangeability^16^) are also required assumptions for CNMA models^9^. Transitivity in a network assumes that the distribution of any effect modifiers (i.e., any variables that may change the true effect of an intervention) is similar on average across intervention comparisons. Consistency is considered the statistical equivalent of transitivity, and in a network, it assumes that the different sources of evidence (i.e., direct and indirect evidence) agree. Numerous approaches have been developed for evaluating transitivity and consistency in NMA,^9, 28–32^ but these approaches have not been extended to CNMA.

Heterogeneity remains a critical concern in CNMA, as in standard NMA, yet current methods to estimate and interpret heterogeneity in component effects are still limited. The use of prediction intervals around component estimates (e.g., assuming a normal distribution with means equal to the component effects and variance equal to the between-study heterogeneity)^33, 34^ may provide insight on the extent of heterogeneity.

Overall, moving from a single-effect model to an additive CNMA, then an interaction CNMA, and finally a standard NMA (full-interaction model) increases model complexity and reduces assumptions, but does not negate important considerations, such as inconsistency and heterogeneity in the network. While this progression may reduce precision of intervention effects, it offers a more detailed understanding of how individual components or combinations contribute to overall effects.

### Application to the prehabilitation interventions example

Transitivity was evaluated using a multi-faceted approach, including review of study and patient characteristics based on evidence tables along with graphical approaches, such as inspection of box plots and bar plots, to examine characteristics of treatment comparisons within the evidence network. As reported in the original publication, no evidence of intransitivity was identified when mean age, proportion female, control group risk, year of publication, and surgery type were assessed as potential effect modifiers (see also Supplementary File, Appendix 14 of original publication).^4^

The design-by-treatment interaction model^28^ as an extension of the standard NMA model and assuming unique combinations of components as different nodes, suggested no statistical evidence for inconsistency (Q_between-designs_ = 3.41, *df* = 2, p-value = 0.18, τ^2^ = 0.14). However, local assessment of inconsistency using the back-calculation method^29^ indicated that the direct evidence in *EXE* vs *US* was borderline inconsistent with the remaining network at the 5% significance level (p-value = 0.07). Exploration of evidence of inconsistency was conducted through network meta-regression for control group risk and surgery type (a summary of these explorations is included in Supplementary File, Appendix 15 of original publication). The certainty rating for that comparison was downgraded due to inconsistency. Assessments of both transitivity and consistency were informed by team discussions, incorporating clinical considerations relevant to prehabilitation. These discussions also guided the interpretation of results once the analyses were complete.

## How do we report results in component network meta-analysis?

### Methods and presentation of findings

When reporting the findings from a CNMA, it is essential to ensure that the results are aligned with the review question and relevant to clinicians, policymakers, and patient partners. The selection of the primary analysis model (e.g., additive CNMA, interaction CNMA, or standard NMA) should be guided by both methodological considerations (e.g., model fit, transitivity, heterogeneity) and knowledge user priorities, including the clinical plausibility of additivity and component interactions.

For the presentation of CNMA results, typically a reference intervention is defined as the comparator. The reference intervention could be the current, standard intervention or inactive placebo/*UC* in settings where no standard intervention has been established. Graphical approaches can be used to present and compare the findings of intervention and component effects across NMA and CNMA models.^35, 36^

### Application to the prehabilitation interventions example

In this example, we applied random-effects additive and interaction CNMA models. We also fitted standard NMA models for completeness. The common-within network heterogeneity was estimated using the DerSimonian and Laird method.^37^ Results were expressed as odds ratios with corresponding 95% CIs for each model. We calculated P-scores^38^ to rank interventions and used a rank-heat plot^39^ for their presentation across the different models. Additional information on the individual components as obtained from the NMA model are presented in Appendix 7. All analyses, and across all empirical examples, were conducted in a frequentist setting using R packages **netmeta**^18^ and **viscomp.**^40^

The publication of the prehabilitation interventions example^4^ reported both standard NMA and CNMA findings, but prioritized NMA because patient partners and clinical collaborators felt strongly that clinically meaningful interactions between components made standard NMA more appropriate for capturing the distinct effects of each intervention combination.

**Figure 2** presents a forest plot comparing findings from standard NMA, additive CNMA, and interaction CNMA, using *UC* as the reference group. The interaction CNMA shown corresponds to Model 1 from **Table 1**, which includes a single interaction term, *EXE*NUT*, and yielded a likely spuriously significant reduction in the Q statistic. Based on these results, had this interaction model been selected over the additive model, the *EXE*NUT* interaction term (OR_EXE*NUT_=1.52, 95% CI 0.86 to 2.70) would have acted to decrease the combined impact of *EXE* and *NUT* in the *EXE* + *NUT* intervention (i.e., *EXE* and *NUT* acted antagonistically, so that the *EXE* + *NUT* combination then appears less effective than in the additive model; CNMA OR_EXE+NUT_=0.35, 95% CI 0.26 to 0.46; interaction CNMA OR_EXE+NUT_=0.48, 95% CI 0.29 to 0.80), see Figure 2. This finding would have been counter to current clinical thinking, potentially reinforcing the preference for the additive model. However, given that the assumption of additivity held, the estimated effects of combinations of components could be calculated from the incremental effects (OR_EXE_=0.53, 95% CI 0.42 to 0.66; OR_NUT_=0.66, 95% CI 0.54 to 0.81), after converting odds ratios to LORs. After consulting with the team and incorporating clinical and patient partner input, parsimony was prioritized over marginal decreases in the Q statistic. This decision was guided by the understanding that component effects estimated from an interaction model cannot be generalized beyond the specific combinations observed in the dataset.

**Figure 2.**
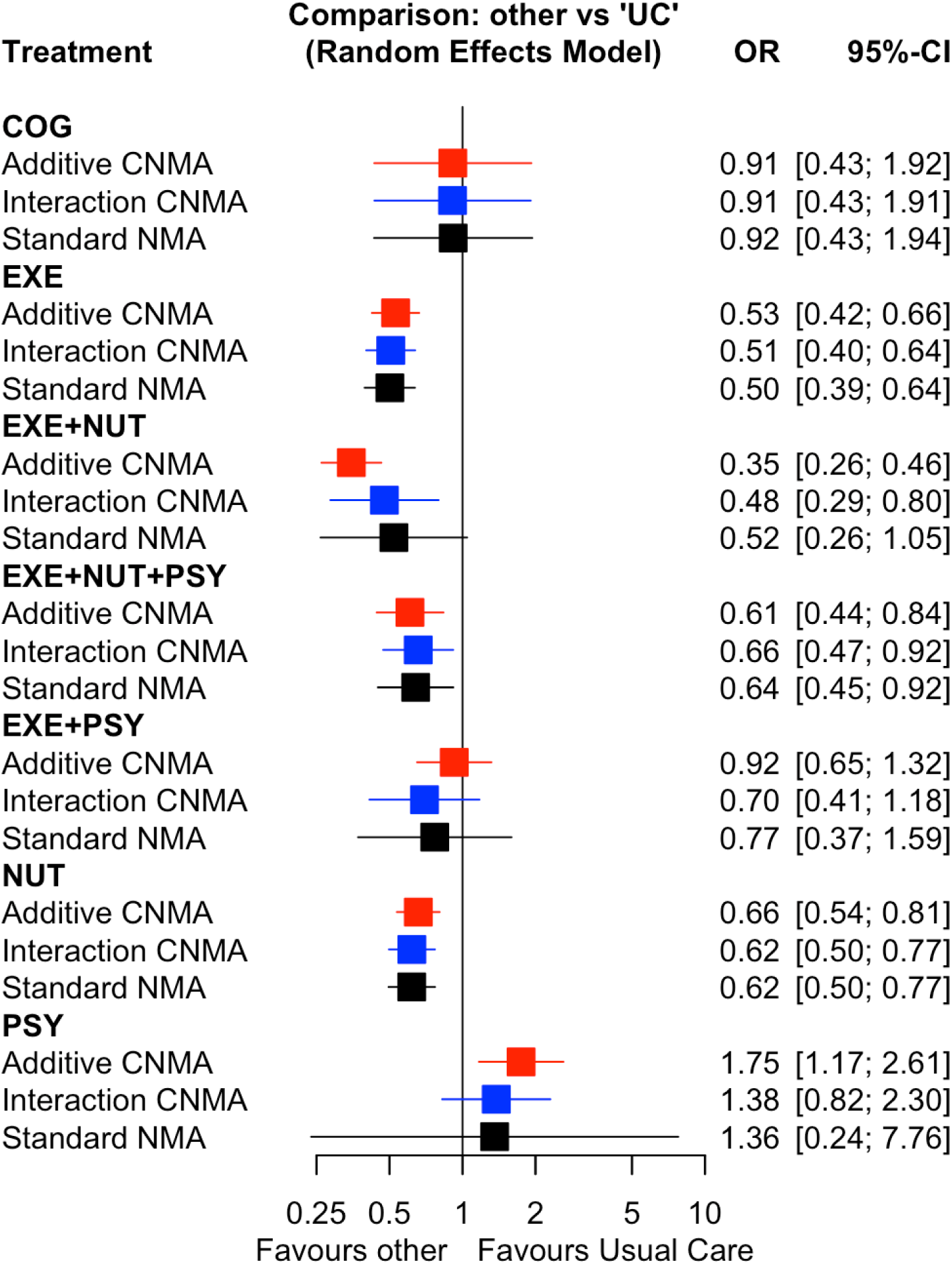
Interval plot of intervention effects (odds ratios) with corresponding 95% confidence intervals for the prevention of post-operative complications. Results are shown for three models: standard network meta-analysis (NMA, black font), additive component network meta-analysis (CNMA, red font), and interaction CNMA with *EXE*NUT* interaction term (Model 1, blue font).

## Discussion

Complex interventions are increasingly evaluated in healthcare trials with growing methodological interest. The comparative efficacy of intervention components should be determined in consideration of their potential interactions and contextual factors. CNMA allows for the assessment and disentanglement of individual component effects: the additive CNMA model assumes full additivity of component effects. Extending the additive CNMA to include terms that account for interactions between components leads to the interaction CNMA model. The full interaction model, considering all possible interactions between components, is the standard NMA model.^16^

Under the additivity assumption, the additive CNMA model improves power and precision compared to standard NMA. When additivity is violated, the interaction CNMA is suggested to generate less-biased component effects. Interaction CNMA provides a compromise between additive CNMA and standard NMA, especially when component effects may be synergistic or antagonistic. However, interactions can be hard to identify. As also noted in our empirical examples, usually the amount of data in a network is too sparse to test for many interactions, and *a priori* selection of clinically relevant interactions should be considered for modelling.

CNMA has several benefits over standard NMA: it estimates fewer parameters, borrows strength from studies sharing components, can provide more powerful and precise results,^17^ and can connect disconnected networks with common components, when there is strong clinical evidence that additivity can be assumed.^22^ However, challenges remain which may affect the validity, interpretability, and certainty of CNMA results. These include decisions on component definition and selection, coding strategies to build a network graph, data availability, sparse data (i.e., few studies against component combinations), differences in usual care across studies, heterogeneity, and inconsistency in the network. Component selection in CNMA is analogous to node-making in standard NMA and represents a critical methodological decision that requires clear justification and transparency.^41–44^ Key considerations include the process for identifying components, consistency of component reporting in the primary literature, and the number of components selected (e.g., too many components may result in sparse data). Available structured frameworks can help guide consistent and replicable component classification.^45–47^

Currently, frameworks such as CINeMA^48^ (Confidence in Network Meta-Analysis) and GRADE^49^ (Grading of Recommendations, Assessment, Development, and Evaluations), which assess the certainty of evidence in standard NMAs, have not yet been formally extended to CNMA, and can be applied only to overall intervention effects derived from standard NMA rather than individual component estimates. The certainty of evidence for component effects remains an open methodological question, as does the assessment of transitivity and consistency in the network. Future advancements in CNMA may address current limitations, including advancements in the assessment of component consistency (e.g., between additive effects and direct comparisons), ranking of components, component contribution to intervention effects, and component effect modelling in multivariate CNMA, along with inherent challenges these bring. For instance, intervention rankings in NMA can vary depending on the ranking metric used, as each metric answers a different type of ranking question.^50^ Also, current ranking approaches do not incorporate assessments of the certainty of evidence, which is particularly important in cases where an intervention with a small sample size or high risk of bias ranks highest, potentially leading to misleading or overly confident interpretations of treatment hierarchies.^51, 52^ We anticipate that similar challenges apply to CNMA, and future developments should aim to integrate certainty assessments into both component and intervention rankings. Future simulation studies should provide guidance on the minimum number of studies required for reliable CNMA estimation across varying numbers of components and under different modeling assumptions (e.g., additivity or interaction).

Although the use of NMA has increased in the last two decades,^16, 53^ CNMA has not been widely used. Provided that there is an increased clinical interest in the assessment of the efficacy of individual components in complex interventions,^21, 54^ and given the freely available R package **netmeta,**^18^ we expect a rise in the use of CNMA models. CNMA should be considered when interventions share clearly defined components and there is clinical plausibility for assuming additive or interactive effects among components. CNMA may not be feasible in networks with sparse data (e.g., few studies per component or combination), poorly reported components, or when strong interactions between components violate the additivity assumption without sufficient data to model interactions. Both CNMA and standard NMA rely on key assumptions that require clinical considerations to inform their assessment. Ultimately, the choice between the models depends on clinical relevance, the structure of the evidence base, model complexity, and data sufficiency. Thus, to enhance the adoption and familiarity of CNMA approaches, there is an urgent need to train researchers involved in evidence synthesis in all aspects of CNMA, from planning and protocol development to implementation,^55^ model selection,^22^ and presentation of results.^35, 36^ Trialists should also provide more information about complex interventions, such as using the TIDIER (template for intervention description and replication)^56^ framework for reporting interventions, which can help inform CNMA.

## Conclusions

Component effect estimation is important to produce relevant and clinically meaningful evidence synthesis results for knowledge users. Implementing CNMA requires multidisciplinary expertise and careful clinical, statistical and methodological planning. Different CNMA approaches and decisions (e.g., about interactions) may generate important variations in results so, when possible, decisions in CNMA model development should be made *a priori* at the protocol stage. Future research to guide selection of CNMA models is critical in developing a consensus-based approach and advancing evidence synthesis methods incorporating CNMA.

## Supporting information

R code example

Data set

R code examples

## Data Sharing

The datasets of the empirical examples and statistical code are available in the supplementary material.

## Acknowledgements

We thank Brahmleen Kaur for their support in formatting the paper.

## Dissemination

We will disseminate our results to relevant knowledge users (e.g., researchers, trainees and healthcare providers).

## Ethics Approval

Not applicable.

## Patient and Public Involvement Statement

This study did not involve patient or public participation in its design, conduct, reporting, or dissemination, as it was not considered relevant or applicable to the aims of this methodological study.

## Transparency Statement

The lead author, AAV, (the manuscript’s guarantor) affirms that the manuscript is an honest, accurate, and transparent account of the study being reported; that no important aspects of the study have been omitted; and that any discrepancies from the study as planned (and, if relevant, registered) have been explained.

## Contributors and sources

Dr. Areti Angeliki Veroniki (AAV) is a statistician with network meta-analysis expertise. She is co-chair of the Methods Executive and co-Convenor of the Statistical Methods Group in Cochrane. Dr. Guido Schwarzer (GS) and Dr. Dan Jackson (DJ) are statisticians with expertise in developing and applying network meta-analysis models. Dr. Dianna Wolfe (DW) is an epidemiologist, Dr. Brian Hutton (BH) and Andrea C. Tricco (ACT) are methodologists, Dr. Daniel I McIsaac (DIM) is an anesthesiologist, and Dr. Sharon E. Straus (SES) is a geriatrician – all have expertise and experience in conducting systematic reviews with network meta-analysis to support clinical and policy decision-making.

AAV developed the initial R code, completed analyses, drafted the initial manuscript, and integrated co-author feedback. DW and GS contributed to revising the R code and conducted data analyses. AAV conceived the study idea, and all authors contributed to the conception and design of this study. AAV drafted the first version of the manuscript, and all authors contributed to the manuscript’s revision and interpretation of findings. AAV is the guarantor of this article.

We used empirical data from three published systematic reviews.^3, 4, 6^

