## Appendices for "Analysing complex interventions using component network meta-analysis"

### Appendix 1. Outcome data for prevention of post-operative complications, falls-related fractures and quality of life examples.

| ***Post-operative complications*** | | | | | | | | | | | | |
| --- | --- | --- | --- | --- | --- | --- | --- | --- | --- | --- | --- | --- |
| **Author-Year** | | **Events/ Sample size Group 1** | | **Events/ Sample size Group 2** | | **Events/ Sample size Group 3** | | **Treatment Group 1** | | **Treatment Group 2** | **Treatment Group 3** | |
| Lloréns 2014^1^ | | 1/21 | | 0/23 | |  | | *UC* | | *EXE* |  | |
| Birch 2020^2^ | | 1/29 | | 1/31 | |  | | *UC* | | *PSY* |  | |
| Moug 2018^3^ | | 12/19 | | 12/17 | |  | | *UC* | | *EXE + PSY* |  | |
| Kikuchi 2016^4^ | | 17/38 | | 12/39 | |  | | *UC* | | *NUT* |  | |
| Swaminathan 2020^5^ | | 14/29 | | 5/29 | |  | | *UC* | | *EXE* |  | |
| Hollis 2019^6^ | | 2/14 | | 0/20 | |  | | *UC* | | *NUT* |  | |
| Northgraves 2020^7^ | | 4/11 | | 3/10 | |  | | *UC* | | *EXE* |  | |
| Jantharapatta 2020^8^ | | 5/31 | | 4/31 | |  | | *UC* | | *NUT* |  | |
| O'Gara 2020^9^ | | 3/20 | | 5/20 | |  | | *UC* | | *COG* |  | |
| Liu 2020^10^ | | 5/36 | | 4/37 | |  | | *UC* | | *EXE + NUT + PSY* |  | |
| Karlsson 2019^11^ | | 2/11 | | 6/10 | |  | | *UC* | | *EXE* |  | |
| Ausania 2019^12^ | | 12/22 | | 6/18 | |  | | *UC* | | *EXE + NUT* |  | |
| Vlisides 2019^13^ | | 5/29 | | 6/23 | |  | | *UC* | | *COG* |  | |
| Chakravartty 2019^14^ | | 2/10 | | 1/10 | |  | | *UC* | | *NUT* |  | |
| Kanekiyo 2019^15^ | | 11/20 | | 4/20 | |  | | *UC* | | *NUT* |  | |
| Valkenet 2018^16^ | | 43/121 | | 47/120 | |  | | *UC* | | *EXE* |  | |
| Ruiz-Tovar 2019^17^ | | 1/20 | | 0/20 | |  | | *UC* | | *NUT* |  | |
| Kong 2018^18^ | | 23/62 | | 19/65 | |  | | *UC* | | *NUT* |  | |
| Mudge 2018^19^ | | 32/66 | | 34/65 | |  | | *UC* | | *NUT* |  | |
| Grundmann 2018^20^ | | 19/40 | | 15/36 | |  | | *UC* | | *NUT* |  | |
| Palma-Milla 2016^21^ | | 10/16 | | 8/17 | |  | | *UC* | | *NUT* |  | |
| Banerjee 2018^22^ | | 7/28 | | 6/27 | |  | | *UC* | | *EXE* |  | |
| Barberan-Garcia 2018^23^ | | 39/63 | | 19/62 | |  | | *UC* | | *EXE + NUT + PSY* |  | |
| Lai 2017^24^ | | 14/50 | | 5/51 | |  | | *UC* | | *EXE* |  | |
| Ida 2017^25^ | | 8/60 | | 9/63 | |  | | *UC* | | *NUT* |  | |
| Licker 2016^26^ | | 37/77 | | 25/74 | |  | | *UC* | | *EXE* |  | |
| Lai 2019^27^ | | 12/34 | | 4/34 | |  | | *UC* | | *EXE* |  | |
| Huang 2017^28^ | | 12/30 | | 5/30 | | 8/30 | | *UC* | | *EXE + PSY* | *EXE* | |
| Burnand 2016^29^ | | 2/25 | | 1/21 | |  | | *UC* | | *NUT* |  | |
| Dunne 2016^30^ | | 7/15 | | 8/19 | |  | | *UC* | | *EXE* |  | |
| Kaya 2016^31^ | | 12/27 | | 6/31 | |  | | *UC* | | *NUT* |  | |
| Kalarchian 2016^32^ | | 0/72 | | 1/71 | |  | | *UC* | | *PSY* |  | |
| Ruiz-Tovar 2015^33^ | | 1/20 | | 1/40 | |  | | *UC* | | *NUT* |  | |
| Kabata 2014^34^ | | 17/48 | | 8/54 | |  | | *UC* | | *NUT* |  | |
| Bergin 2014^35^ | | 3/56 | | 1/50 | |  | | *UC* | | *EXE* |  | |
| Falewee 2014^36^ | | 35/64 | | 33/73 | |  | | *UC* | | *NUT* |  | |
| Oosting 2012^37^ | | 10/14 | | 7/12 | |  | | *UC* | | *EXE* |  | |
| Pehlivan 2011^38^ | | 5/30 | | 1/30 | |  | | *UC* | | *EXE* |  | |
| Burden 2011^39^ | | 26/62 | | 24/54 | |  | | *UC* | | *NUT* |  | |
| Benzo 2011^40^ | | 5/8 | | 3/9 | |  | | *UC* | | *EXE* |  | |
| Bitterli 2011^41^ | | 0/37 | | 3/36 | |  | | *UC* | | *EXE* |  | |
| Rosenfeldt 2011^42^ | | 19/57 | | 22/60 | |  | | *UC* | | *EXE + PSY* |  | |
| Ishikawa 2010^43^ | | 3/13 | | 2/11 | |  | | *UC* | | *NUT* |  | |
| Kulkarni 2010^44^ | | 2/19 | | 1/57 | |  | | *UC* | | *EXE* |  | |
| Hoogeboom 2010^45^ | | 0/10 | | 1/10 | |  | | *UC* | | *EXE* |  | |
| Okamoto 2009^46^ | | 4/30 | | 4/30 | |  | | *UC* | | *NUT* |  | |
| Ferreira 2009^47^ | | 0/15 | | 3/15 | |  | | *UC* | | *EXE* |  | |
| Furze 2009^48^ | | 1/90 | | 1/86 | |  | | *UC* | | *PSY* |  | |
| Nielsen 2008^49^ | | 8/32 | | 8/28 | |  | | *UC* | | *EXE* |  | |
| Dronkers 2007^50^ | | 8/10 | | 3/10 | |  | | *UC* | | *EXE* |  | |
| Hulzebos 2006a^51^ | | 48/137 | | 25/139 | |  | | *UC* | | *EXE* |  | |
| Rooks 2006^52^ | | 8/39 | | 3/39 | |  | | *UC* | | *EXE* |  | |
| Xu 2006^53^ | | 8/30 | | 2/30 | |  | | *UC* | | *NUT* |  | |
| Gocen 2004^54^ | | 1/30 | | 1/29 | |  | | *UC* | | *EXE* |  | |
| Beaupre 2003^55^ | | 6/60 | | 3/55 | |  | | *UC* | | *EXE* |  | |
| Schueren 2001^56^ | | 7/15 | | 10/17 | |  | | *UC* | | *NUT* |  | |
| Braga 1999^57^ | | 21/86 | | 9/85 | |  | | *UC* | | *NUT* |  | |
| Roukema 1988^58^ | | 50/84 | | 13/69 | |  | | *UC* | | *EXE* |  | |
| Argunova 2018^59^ | | 3/18 | | 2/20 | |  | | *UC* | | *EXE* |  | |
| Fulop 2021^60^ | | 16/72 | | 17/77 | |  | | *UC* | | *EXE + NUT + PSY* |  | |
| Da Cunha 2013^61^ | | 4/10 | | 4/12 | |  | | *UC* | | *EXE* |  | |
| Moya 2016^62^ | | 43/122 | | 28/122 | |  | | *UC* | | *NUT* |  | |
| Gloor 2022^63^ | | 2/53 | | 7/54 | |  | | *UC* | | *EXE* |  | |
| Lee 2021^64^ | | 24/82 | | 25/79 | |  | | *UC* | | *NUT* |  | |
| Berkel 2022^65^ | | 21/29 | | 12/28 | |  | | *UC* | | *EXE* |  | |
| Bhattacharyya 2021^66^ | | 8/40 | | 7/40 | |  | | *UC* | | *NUT* |  | |
| Steffens 2021^67^ | | 7/11 | | 10/11 | |  | | *UC* | | *EXE* |  | |
| Lopez-Rodriguez-Arias 2021^68^ | | 5/10 | | 2/10 | |  | | *UC* | | *EXE + NUT + PSY* |  | |
| Sittitrai 2021^69^ | | 14/56 | | 10/60 | |  | | *UC* | | *NUT* |  | |
| IJmker-Hemink 2020^70^ | | 10/49 | | 9/48 | |  | | *UC* | | *NUT* |  | |
| Chen 2017^71^ | | 11/37 | | 3/31 | |  | | *UC* | | *NUT* |  | |
| Sawatzky 2014^72^ | | 4/7 | | 2/8 | |  | | *UC* | | *EXE + NUT + PSY* |  | |
| Soares 2013^73^ | | 11/16 | | 5/16 | |  | | *UC* | | *EXE* |  | |
| Gillis 2016^74^ | | 9/21 | | 8/22 | |  | | *UC* | | *NUT* |  | |
| Carli 2020^75^ | | 25/55 | | 25/55 | |  | | *UC* | | *EXE + NUT + PSY* |  | |
| van Nieuwenhove 2011^76^ | | 18/136 | | 8/137 | |  | | *UC* | | *NUT* |  | |
| Minnella 2018^77^ | | 18/25 | | 14/24 | |  | | *UC* | | *EXE + NUT* |  | |
| Liang 2018^78^ | | 6/34 | | 3/44 | |  | | *UC* | | *EXE + NUT + PSY* |  | |
| Barth 2018^79^ | | 5/27 | | 6/30 | |  | | *UC* | | *NUT* |  | |
| Gillis 2014^80^ | | 17/39 | | 12/38 | |  | | *UC* | | *EXE + NUT + PSY* |  | |
| Humeidan 2020^81^ | | 29/126 | | 18/125 | |  | | *UC* | | *COG* |  | |
| Bousquet-Dion 2018^82^ | | 8/26 | | 14/37 | |  | | *UC* | | *EXE + NUT + PSY* |  | |
| Santa Mina 2018^83^ | | 14/40 | | 18/42 | |  | | *UC* | | *EXE* |  | |
| De Luis 2012^84^ | | 1/20 | | 1/20 | |  | | *UC* | | *NUT* |  | |
| Ismail 2016^85^ | | 6/26 | | 2/24 | |  | | *UC* | | *EXE* |  | |
| Abdelaal 2017^86^ | | 15/24 | | 7/26 | |  | | *UC* | | *EXE* |  | |
| D'Lima 1996^87^ | | 2/10 | | 4/20 | |  | | *UC* | | *EXE* |  | |
| Yegen 2020^88^ | | 23/39 | | 15/39 | |  | | *UC* | | *NUT* |  | |
| Lai 2016^89^ | | 5/24 | | 2/24 | |  | | *UC* | | *EXE* |  | |
| Gade 2016^90^ | | 15/16 | | 15/19 | |  | | *UC* | | *NUT* |  | |
| Dronkers 2010^91^ | | 9/20 | | 8/21 | |  | | *UC* | | *EXE* |  | |
| McCarthy 2008^92^ | | 6/7 | | 6/8 | |  | | *UC* | | *NUT* |  | |
| Hulzebos 2006b^93^ | | 6/12 | | 2/14 | |  | | *UC* | | *EXE* |  | |
| Allen 2021^94^ | | 18/23 | | 13/22 | |  | | *UC* | | *EXE + NUT + PSY* |  | |
| Diaz-Feijoo 2022^95^ | | 12/19 | | 6/15 | |  | | *UC* | | *EXE + NUT + PSY* |  | |
| Labuschagne 2022^96^ | | 4/6 | | 2/6 | |  | | *UC* | | *EXE* |  | |
| McIsaac 2022^97^ | | 52/88 | | 42/94 | |  | | *UC* | | *EXE + NUT* |  | |
| Sykes 2022^98^ | | 16/25 | | 15/24 | |  | | *UC* | | *NUT* |  | |
| Tesar 2022^99^ | | 19/60 | | 22/60 | |  | | *UC* | | *NUT* |  | |
| Xia 2022^100^ | | 34/78 | | 8/78 | |  | | *UC* | | *EXE* |  | |
| Atoui 2023^101^ | | 7/46 | | 10/43 | |  | | *UC* | | *EXE + NUT + PSY* |  | |
| Chen 2023a^102^ | | 13/109 | | 4/109 | |  | | *UC* | | *EXE* |  | |
| Molenaar 2023^103^ | | 54/128 | | 39/123 | |  | | *UC* | | *EXE + NUT + PSY* |  | |
| Chen 2023b^104^ | | 34/119 | | 9/120 | |  | | *UC* | | *NUT* |  | |
| Zhang 2023^105^ | | 7/70 | | 5/70 | |  | | *UC* | | *EXE* |  | |
| Wall 2023^106^ | | 2/5 | | 1/8 | |  | | *UC* | | *NUT* |  | |
| ***Falls-related fractures*** | | | | | | | | | | | | |
| **Author-Year** | **Events/ Sample size**  **Group 1** | | **Events/ Sample size**  **Group 2** | | **Events/ Sample size**  **Group 3** | | **Treatment Group 1** | | **Treatment Group 2** | | | **Treatment Group 3** |
| Vind 2009^107^ | 24/196 | | 16/196 | |  | | *cl-qi + mf* | | *uc* | | |  |
| Sakamoto 2013^108^ | 11/455 | | 4/410 | |  | | *uc* | | *ex* | | |  |
| Sambrook 2012^109^ | 13/207 | | 18/205 | | 19/190 | | *ca + vi-d* | | *pa-qi* | | | *vi-d* |
| Perula 2012^110^ | 1/133 | | 2/271 | |  | | *cl-qi + ex + mf + pa-qi* | | *pa-qi* | | |  |
| Glendenning 2012^111^ | 10/353 | | 10/333 | |  | | *pa-qi + vi-d* | | *pa-qi* | | |  |
| Pekkarinen 2013^112^ | 95/1174 | | 59/1004 | |  | | *uc* | | *cl-qi + mf + pa-qi* | | |  |
| Neelemaat 2012^113^ | 1/105 | | 0/105 | |  | | *uc* | | *ca + cl-qi + di + pa-qi + vi-d* | | |  |
| Palvanen 2014^114^ | 42/653 | | 33/661 | |  | | *pa-qi* | | *cl-qi + mf + pa-qi* | | |  |
| Suman 2011^115^ | 3/123 | | 2/96 | |  | | *cl-qi + mf* | | *cl-qi + mf + pa-qi* | | |  |
| Dangour 2011^116^ | 5/389 | | 6/440 | | 10/392 | | *uc* | | *di + ex* | | | *ex* |
| Spink 2011^117^ | 1/153 | | 7/152 | |  | | *bf + cp + ex + pa-qi* | | *bf* | | |  |
| Cameron 2011b^118^ | 1/101 | | 1/97 | |  | | *pa-qi* | | *cl-qi + de-hp* | | |  |
| Haines 2011^119^ | 2/381 | | 1/401 | |  | | *bf + cl-qi* | | *bf + cl-qi + pa-qi* | | |  |
| Drahota 2013^120^ | 0/210 | | 2/208 | |  | | *fl* | | *uc* | | |  |
| Sanders 2010^121^ | 135/1125 | | 171/1131 | |  | | *uc* | | *vi-d* | | |  |
| Iliffe 2014^122^ | 4/323 | | 1/273 | | 1/289 | | *uc* | | *ex* | | | *ex + pa-qi* |
| de Vries 2010^123^ | 5/111 | | 5/106 | |  | | *uc* | | *cl-qi + mf* | | |  |
| Meyer 2009^124^ | 40/551 | | 42/574 | |  | | *cl-qi* | | *bf + cl-qi* | | |  |
| Koike 2009^125^ | 36/345 | | 67/327 | |  | | *de-hp + pa-qi* | | *pa-qi* | | |  |
| Sahota 2014^126^ | 3/857 | | 2/863 | |  | | *uc* | | *de-al* | | |  |
| Salminen 2009^127^ | 28/298 | | 27/293 | |  | | *pa-qi* | | *cl-qi + mf + pa-qi + so* | | |  |
| Spice 2009^128^ | 27/120 | | 22/103 | | 30/159 | | *bf* | | *cl-qi + mf* | | | *mf* |
| Rosendahl 2008^129^ | 3/55 | | 5/64 | |  | | *ex* | | *so* | | |  |
| Cumming 2008^130^ | 2/1836 | | 3/1751 | |  | | *cl-qi + de-al + ex + mf + pa-qi* | | *uc* | | |  |
| Prince 2008^131^ | 4/151 | | 3/151 | |  | | *ca + vi-d* | | *ca* | | |  |
| Berggren 2008^132^ | 11/97 | | 7/102 | |  | | *uc* | | *ca + cl-qi + mf + vi-d* | | |  |
| Burleigh 2007^133^ | 3/103 | | 1/100 | |  | | *ca* | | *ca + vi-d* | | |  |
| Smith 2007^134^ | 306/4727 | | 279/4713 | |  | | *vi-d* | | *uc* | | |  |
| Cumming 2007^135^ | 18/307 | | 31/309 | |  | | *uc* | | *ea + va* | | |  |
| Korpelainen 2006^136^ | 6/84 | | 16/76 | |  | | *ex* | | *uc* | | |  |
| Foss 2006^137^ | 3/103 | | 5/115 | |  | | *uc* | | *su-ey* | | |  |
| Law 2006^138^ | 53/1955 | | 66/1762 | |  | | *uc* | | *vi-d* | | |  |
| Harwood 2005^139^ | 12/152 | | 4/154 | |  | | *uc* | | *su-ey* | | |  |
| Grant 2005^140^ | 212/1343 | | 184/1306 | |  | | *vi-d* | | *ca + vi-d* | | |  |
| Davison 2005^141^ | 7/159 | | 13/154 | |  | | *ea + ex + mf* | | *uc* | | |  |
| Ray 2005^142^ | 241/5064 | | 218/4439 | |  | | *uc* | | *cl-qi + mf + pa-qi* | | |  |
| Barr 2005^143^ | 85/1276 | | 24/599 | |  | | *uc* | | *bf* | | |  |
| Flicker 2005^144^ | 35/312 | | 25/313 | |  | | *ca* | | *ca + vi-d* | | |  |
| Sato 2005b^145^ | 27/230 | | 8/231 | |  | | *ca + vi-d* | | *ca + op-tx + vi-d* | | |  |
| Dyer 2004^146^ | 3/60 | | 2/56 | |  | | *cl-qi + ex + mf* | | *uc* | | |  |
| Cameron 2003^147^ | 49/298 | | 52/302 | |  | | *uc* | | *cl-qi + de-hp* | | |  |
| Becker 2003^148^ | 31/485 | | 33/450 | |  | | *de-hp + ea + ex + pa-qi* | | *uc* | | |  |
| Nikolaus 2003^149^ | 4/181 | | 3/179 | |  | | *cl-qi + mf* | | *mf* | | |  |
| Birks 2003^150^ | 24/182 | | 16/184 | |  | | *de-hp + pa-qi* | | *pa-qi* | | |  |
| Hogan 2001^151^ | 3/79 | | 5/84 | |  | | *mf* | | *so* | | |  |
| Kenny 2001^152^ | 3/84 | | 4/87 | |  | | *bf + su-pm* | | *bf* | | |  |
| Cameron 2001^153^ | 13/88 | | 14/86 | |  | | *uc* | | *cl-qi + de-hp* | | |  |
| McMurdo 2000^154^ | 1/71 | | 3/52 | |  | | *ex + mf* | | *so* | | |  |
| Fairhall 2014^155^ | 12/121 | | 13/120 | |  | | *uc* | | *cl-qi + ex + mf* | | |  |
| Gianoudis 2014^156^ | 1/81 | | 0/81 | |  | | *cl-qi + ex + pa-qi* | | *pa-qi* | | |  |
| Kim 2014^157^ | 1/52 | | 2/53 | |  | | *ex* | | *pa-qi* | | |  |
| Pitkala 2013^158^ | 4/65 | | 5/61 | |  | | *pa-qi* | | *ex* | | |  |
| Sherrington 2014^159^ | 15/169 | | 14/171 | |  | | *pa-qi* | | *ex + pa-qi* | | |  |
| Sjoberg 2013^160^ | 15/99 | | 16/100 | |  | | *uc* | | *bf* | | |  |
| Larsen 2005^161^ | 309/4813 | | 325/4916 | | 162/2054 | | *ca + pa-qi + vi-d* | | *ea + pa-qi* | | | *uc* |
| Lips 1996^162^ | 122/1287 | | 135/1291 | |  | | *ca* | | *ca + vi-d* | | |  |
| Patil 2015^163^ | 12/204 | | 8/205 | |  | | *uc* | | *ex* | | |  |
| Gleeson 2015^164^ | 1/60 | | 2/60 | |  | | *ex* | | *uc* | | |  |
| Parry 2016^165^ | 3/210 | | 7/205 | |  | | *cbt + cl-qi + pa-qi* | | *uc* | | |  |
| Cockayne 2017^166^ | 14/507 | | 19/493 | |  | | *pa-qi* | | *cp + de-ft + pa-qi* | | |  |
| Chapuy 1994^167^ | 301/1176 | | 368/1127 | |  | | *ca + vi-d* | | *uc* | | |  |
| Chapuy 2002^168^ | 97/393 | | 55/190 | |  | | *ca + vi-d* | | *uc* | | |  |
| Gallagher 2001^169^ | 10/122 | | 8/122 | | 16/122 | | *uc* | | *op-tx + vi-d* | | | *op-tx* |
| Ferrer 2014^170^ | 8/127 | | 2/98 | |  | | *mf* | | *uc* | | |  |
| Pfeifer 2000^171^ | 6/67 | | 3/70 | |  | | *ca* | | *ca + vi-d* | | |  |
| Reid 2006^172^ | 204/732 | | 221/739 | |  | | *ca* | | *uc* | | |  |
| Trivedi 2003^173^ | 149/1341 | | 119/1345 | |  | | *uc* | | *vi-d* | | |  |
| Vetter 1992^174^ | 14/324 | | 16/350 | |  | | *cl-qi* | | *cl-qi + ex + mf* | | |  |
| ***Quality of life*** | | | | | | | | | | | | |
| **Author-Year** | **Mean (SD), Sample size**  **Group 1** | | **Mean, (SD), Sample size Group 2** | | **Mean, (SD), Sample size**  **Group 3** | | **Treatment Group 1** | | **Treatment Group 2** | | | **Treatment Group 3** |
| Bucknall 2012^175^ | -1.38 (11.33), 232 | | 2.99 (12.56), 232 | |  | | *UC* | | *c-M + s-M* | | |  |
| Chi 2012^176^ | -34.53 (14.85), 87 | | -21.11 (18.30), 84 | |  | | *UC* | | *s-M* | | |  |
| Community Pharmacy Medicines Management Project Evaluation Team 2007^177^ | 0.58 (0.07), 422 | | 0.58 (0.07), 810 | |  | | *UC* | | *te + px_educ* | | |  |
| Fan 2012^178^ | -51.80 (13.30), 217 | | -51.30 (13.30), 209 | |  | | *UC* | | *c-M + s-M* | | |  |
| Freund 2016^179^ | 35.50 (10.20), 711 | | 36.50 (10.60), 777 | |  | | *UC* | | *te + staff + finan + c-M + s-M + px_educ* | | |  |
| Hernández 2015^180^ | -49.00 (22.00), 84 | | -43.00 (20.00), 71 | |  | | *UC* | | *c-M + s-M* | | |  |
| Kennedy 2013^181^ | 0.60 (0.30), 409 | | 0.60 (0.30), 277 | |  | | *UC* | | *staff + s-M* | | |  |
| Nguyen 2013^182^ | 35.20 (11.27), 41 | | 38.90 (11.21), 43 | | 35.80 (12.00), 41 | | *px_educ* | | *te + c-M + s-M + px_re + px_educ* | | | *te + c-M + s-M + px_educ + mot_int* |
| Pinnock 2013^183^ | -67.30 (17.30), 128 | | -68.20 (16.30), 128 | |  | | *UC* | | *relay + s-M + px_educ* | | |  |
| van de Ven 2013^184^ | 0.36 (0.22), 79 | | 0.35 (0.26), 49 | |  | | *UC* | | *staff* | | |  |
| Xi 2015^185^ | -63.66 (3.6), 30 | | -59.60 (5.40), 30 | |  | | *px_educ* | | *s-M + px_educ* | | |  |
| Bosanquet 2017^186^ | 35.1 (12.11), 236 | | 34.00 (13.51), 249 | |  | | *UC* | | *te + EPR + relay + c-M + s-M* | | |  |
| Forster 2015^187^ | 0.56 (0.48), 128 | | 0.51 (0.46), 134 | |  | | *UC* | | *te + c-M + s-M + px_educ* | | |  |
| Khdour 2009^188^ | -65.3 (18.62), 87 | | -61.80 (16.55), 86 | |  | | *UC* | | *c-M + s-M* | | |  |
| Markle-Reid 2011^189^ | 47.81 (18.96), 39 | | 48.29 (23.23), 43 | |  | | *UC* | | *te + staff + c-M + s-M + px_educ + px_finan* | | |  |
| Murphy 2009^190^ | 38.8 (11.1), 173 | | 40.50 (11.10), 159 | |  | | *UC* | | *staff + s-M* | | |  |
| Ortiz-Bautista 2017^191^ | -19.8 (23.3), 43 | | -23.46 (22.10), 84 | |  | | *UC* | | *te + c-M + px_educ* | | |  |
| Peters-Klimm 2010^192^ | 38 (8.6), 100 | | 38.30 (8.60), 97 | |  | | *UC* | | *te + EPR + relay + AF + c-M + s-M + px_educ* | | |  |
| Sackley 2015^193^ | 0.18 (0.31), 123 | | 0.20 (0.36), 160 | |  | | *UC* | | *staff + c-M + s-M + px_educ* | | |  |
| Schrader 2005^194^ | 37.6 (10.78), 237 | | 36.70 (7.35), 213 | |  | | *UC* | | *te + staff + c-M + s-M + px_educ* | | |  |
| Van der Aa 2015^195^ | 0.7 (0.3), 134 | | 0.70 (0.30), 131 | |  | | *UC* | | *c-M + s-M + px_educ* | | |  |
| Xiao 2016^196^ | 41.6 (8.7), 30 | | 41.10 (7.70), 31 | |  | | *UC* | | *staff + c-M + s-M* | | |  |
| Zwar 2016^197^ | -17.06 (14.9), 104 | | -16.85 (15.20), 136 | |  | | *UC* | | *EPR + staff* | | |  |
| Schafer 2017^198^ | 0.7 (0.28), 87 | | 0.68 (0.32), 86 | |  | | *UC* | | *staff + s-M + px_educ* | | |  |
| Vaillant-Roussel 2016^199^ | 51.6 (25.5), 83 | | 52.80 (23.80), 76 | |  | | *UC* | | *staff + c-M + s-M + px_educ + mot_int* | | |  |
| Stewart 2015^200^ | 36.6 (12.1), 112 | | 38.10 (10.80), 116 | |  | | *UC* | | *c-M + s-M + px_educ* | | |  |
| Walters 2013^201^ | 38.5 (9.4), 62 | | 38.50 (10.30), 58 | |  | | *UC* | | *staff + c-M + s-M + px_educ* | | |  |
| Wagenaar 2019^202^ | -26.5 (32.5), 150 | | -25.50 (27.80), 150 | | -28.3 (31), 150 | | *UC* | | *EPR + relay + c-M + px_re + px_educ* | | | *s-M + px_re* |
| Liu 2008^203^ | 30.9 (10.78), 24 | | 47.90 (7.35), 24 | |  | | *UC* | | *relay + s-M + px_re + px_educ* | | |  |
| McManus 2014^204^ | 0.806 (0.26), 276 | | 0.82 (0.25), 276 | |  | | *UC* | | *relay + s-M + px_educ* | | |  |
| Galbreath 2004^205^ | 33.4 (13.4), 359 | | 32.10 (12.40), 354 | |  | | *UC* | | *te + staff + c-M + s-M + px_educ* | | |  |
| de la Porte 2007^206^ | -34.5 (23.3), 122 | | -30.20 (22.10), 118 | |  | | *UC* | | *te + c-M + s-M + px_educ* | | |  |
| Eccles 2007^207^ | 30.1 (15.3), 360 | | 29.70 (15.60), 311 | |  | | *UC* | | *EPR + staff + clin_re + px_re* | | |  |
| Lopez Cabezas 2006^208^ | 0.606 (0.18), 64 | | 0.64 (0.15), 70 | |  | | *UC* | | *c-M + px_educ* | | |  |
| Ojeda 2005^209^ | -37.8 (21), 77 | | -35.70 (18.90), 76 | |  | | *UC* | | *s-M + px_educ* | | |  |

**Abbreviations:** *AF*, Audit & feedback; *BF,* Basic falls risk assessment*; CA,* Calcium*; CBT,* Cognitive behavioral therapy*; CLIN_RE*, Clinician reminders*; CL-QI,* Clinic-level quality improvement*; C-M*, Case management*; COG*, Cognitive; *CP,* Comprehensive podiatry assessment and treatment*; DE-AL,* Device—alarm*; DE-FT,* Multifaceted podiatry; *DE-HP,* Device—hip protector*; DI,* Dietary modifications*; EA,* Environmental assessment and modification*; EPR*, Electronic patient registry*; EXE,* Exercise*; FIN*, Financial incentives*; FL,* Flooring*; MF,* Multifactorial assessment and treatment*; MOT_INT*, Motivational interview*; NUT*, Nutrition; *OP-TX,* Osteoporosis treatment*; PA-QI,* Patient-level quality improvement*; PSY*, Psychosocial; *PX_ED*, Patient education*; PX_FIN*, Financial incentives*; PX_RE*, Patient reminders*; RE*, Facilitated relay of info to clinicians*; SD*, Standard Deviation; *S-M*, Promotion of self-management*; SO,* Social engagement*; STA*, Staff education*; SU-EY,* Surgery—cataract*; SU-PM,* Surgery—pacemaker*; TE*, Team changes*; UC,* Usual Care*; VA,* Vision assessment and treatment*; VI-D,* Vitamin D

#### Study References in Appendix 1

1. Llorens J, Rovira L, Ballester M, et al. Preoperative inspiratory muscular training to prevent postoperative hypoxemia in morbidly obese patients undergoing laparoscopic bariatric surgery. A randomized clinical trial. *Obes Surg*. Jun 2015;25(6):1003-9. doi:10.1007/s11695-014-1487-4
2. Birch S, Stilling M, Mechlenburg I, Hansen TB. No effect of cognitive behavioral patient education for patients with pain catastrophizing before total knee arthroplasty: a randomized controlled trial. *Acta Orthop*. Feb 2020;91(1):98-103. doi:10.1080/17453674.2019.1694312
3. Moug SJ, Mutrie N, Barry SJE, et al. Prehabilitation is feasible in patients with rectal cancer undergoing neoadjuvant chemoradiotherapy and may minimize physical deterioration: results from the REx trial. *Colorectal Dis*. May 2019;21(5):548-562. doi:10.1111/codi.14560
4. Kikuchi Y, Hiroshima Y, Matsuo K, et al. A Randomized Clinical Trial of Preoperative Administration of Branched-Chain Amino Acids to Prevent Postoperative Ascites in Patients with Liver Resection for Hepatocellular Carcinoma. *Ann Surg Oncol*. Oct 2016;23(11):3727-3735. doi:10.1245/s10434-016-5348-3
5. Swaminathan N, Kundra P, Ravi R, Kate V. ERAS protocol with respiratory prehabilitation versus conventional perioperative protocol in elective gastrectomy- a randomized controlled trial. *Int J Surg*. Sep 2020;81:149-157. doi:10.1016/j.ijsu.2020.07.027
6. Hollis G, Franz R, Bauer J, Bell J. Implementation of a very low calorie diet program into the pre-operative model of care for obese general elective surgery patients: Outcomes of a feasibility randomised control trial. *Nutr Diet*. Nov 2020;77(5):490-498. doi:10.1111/1747-0080.12601
7. Northgraves MJ, Arunachalam L, Madden LA, et al. Feasibility of a novel exercise prehabilitation programme in patients scheduled for elective colorectal surgery: a feasibility randomised controlled trial. *Support Care Cancer*. Jul 2020;28(7):3197-3206. doi:10.1007/s00520-019-05098-0
8. Jantharapattana K, Orapipatpong O. Efficacy of EPA-enriched supplement compared with standard formula on body weight changes in malnourished patients with head and neck cancer undergone surgery: a randomized study. *Head Neck*. Feb 2020;42(2):188-197. doi:10.1002/hed.25987
9. O'Gara BP, Mueller A, Gasangwa DVI, et al. Prevention of Early Postoperative Decline: A Randomized, Controlled Feasibility Trial of Perioperative Cognitive Training. *Anesth Analg*. Mar 2020;130(3):586-595. doi:10.1213/ANE.0000000000004469
10. Liu Z, Qiu T, Pei L, et al. Two-Week Multimodal Prehabilitation Program Improves Perioperative Functional Capability in Patients Undergoing Thoracoscopic Lobectomy for Lung Cancer: A Randomized Controlled Trial. *Anesth Analg*. Sep 2020;131(3):840-849. doi:10.1213/ANE.0000000000004342
11. Karlsson E, Farahnak P, Franzen E, et al. Feasibility of preoperative supervised home-based exercise in older adults undergoing colorectal cancer surgery - A randomized controlled design. *PLoS One*. 2019;14(7):e0219158. doi:10.1371/journal.pone.0219158
12. Ausania F, Senra P, Melendez R, Caballeiro R, Ouvina R, Casal-Nunez E. Prehabilitation in patients undergoing pancreaticoduodenectomy: a randomized controlled trial. *Rev Esp Enferm Dig*. Aug 2019;111(8):603-608. doi:10.17235/reed.2019.6182/2019
13. Vlisides PE, Das AR, Thompson AM, et al. Home-based Cognitive Prehabilitation in Older Surgical Patients: A Feasibility Study. *J Neurosurg Anesthesiol*. Apr 2019;31(2):212-217. doi:10.1097/ANA.0000000000000569
14. Chakravartty S, Vivian G, Mullholland N, et al. Preoperative liver shrinking diet for bariatric surgery may impact wound healing: a randomized controlled trial. *Surg Obes Relat Dis*. Jan 2019;15(1):117-125. doi:10.1016/j.soard.2018.10.001
15. Kanekiyo S, Takeda S, Iida M, et al. Efficacy of perioperative immunonutrition in esophageal cancer patients undergoing esophagectomy. *Nutrition*. Mar 2019;59:96-102. doi:10.1016/j.nut.2018.08.006
16. Guinan EM, Forde C, O'Neill L, et al. Effect of preoperative inspiratory muscle training on physical functioning following esophagectomy. *Dis Esophagus*. Feb 1 2019;32(2)doi:10.1093/dote/doy091
17. Ruiz-Tovar J, Blanca M, Garcia A, et al. Preoperative administration of Omega-3 fatty acids on postoperative pain and acute-phase reactants in patients undergoing Roux-en-Y gastric bypass: A randomized clinical trial. *Clin Nutr*. Aug 2019;38(4):1588-1593. doi:10.1016/j.clnu.2018.07.026
18. Kong SH, Lee HJ, Na JR, et al. Effect of perioperative oral nutritional supplementation in malnourished patients who undergo gastrectomy: A prospective randomized trial. *Surgery*. Dec 2018;164(6):1263-1270. doi:10.1016/j.surg.2018.05.017
19. Mudge LA, Watson DI, Smithers BM, et al. Multicentre factorial randomized clinical trial of perioperative immunonutrition versus standard nutrition for patients undergoing surgical resection of oesophageal cancer. *Br J Surg*. Sep 2018;105(10):1262-1272. doi:10.1002/bjs.10923
20. Grundmann F, Muller RU, Reppenhorst A, et al. Preoperative Short-Term Calorie Restriction for Prevention of Acute Kidney Injury After Cardiac Surgery: A Randomized, Controlled, Open-Label, Pilot Trial. *J Am Heart Assoc*. Mar 13 2018;7(6)doi:10.1161/JAHA.117.008181
21. Palma-Milla S, Lopez-Plaza B, Santamaria B, de Arriba-Sanchez A, Bermejo LM, Gomez-Candela C. New, Immunomodulatory, Oral Nutrition Formula for Use Prior to Surgery in Patients With Head and Neck Cancer: An Exploratory Study. *JPEN J Parenter Enteral Nutr*. Feb 2018;42(2):371-379. doi:10.1177/0148607116676839
22. Banerjee S, Manley K, Shaw B, et al. Vigorous intensity aerobic interval exercise in bladder cancer patients prior to radical cystectomy: a feasibility randomised controlled trial. *Support Care Cancer*. May 2018;26(5):1515-1523. doi:10.1007/s00520-017-3991-2
23. Barberan-Garcia A, Ubre M, Roca J, et al. Personalised Prehabilitation in High-risk Patients Undergoing Elective Major Abdominal Surgery: A Randomized Blinded Controlled Trial. *Ann Surg*. Jan 2018;267(1):50-56. doi:10.1097/SLA.0000000000002293
24. Lai Y, Su J, Qiu P, et al. Systematic short-term pulmonary rehabilitation before lung cancer lobectomy: a randomized trial. *Interact Cardiovasc Thorac Surg*. Sep 1 2017;25(3):476-483. doi:10.1093/icvts/ivx141
25. Ida S, Hiki N, Cho H, et al. Randomized clinical trial comparing standard diet with perioperative oral immunonutrition in total gastrectomy for gastric cancer. *Br J Surg*. Mar 2017;104(4):377-383. doi:10.1002/bjs.10417
26. Licker M, Karenovics W, Diaper J, et al. Short-Term Preoperative High-Intensity Interval Training in Patients Awaiting Lung Cancer Surgery: A Randomized Controlled Trial. *J Thorac Oncol*. Feb 2017;12(2):323-333. doi:10.1016/j.jtho.2016.09.125
27. Lai Y, Wang X, Zhou K, Su J, Che G. Impact of one-week preoperative physical training on clinical outcomes of surgical lung cancer patients with limited lung function: a randomized trial. *Ann Transl Med*. Oct 2019;7(20):544. doi:10.21037/atm.2019.09.151
28. Huang J, Lai Y, Zhou X, et al. Short-term high-intensity rehabilitation in radically treated lung cancer: a three-armed randomized controlled trial. *J Thorac Dis*. Jul 2017;9(7):1919-1929. doi:10.21037/jtd.2017.06.15
29. Burnand KM, Lahiri RP, Burr N, Jansen van Rensburg L, Lewis MP. A randomised, single blinded trial, assessing the effect of a two week preoperative very low calorie diet on laparoscopic cholecystectomy in obese patients. *HPB (Oxford)*. May 2016;18(5):456-61. doi:10.1016/j.hpb.2016.01.545
30. Dunne DF, Jack S, Jones RP, et al. Randomized clinical trial of prehabilitation before planned liver resection. *Br J Surg*. Apr 2016;103(5):504-12. doi:10.1002/bjs.10096
31. Kaya SO, Akcam TI, Ceylan KC, Samancilar O, Ozturk O, Usluer O. Is preoperative protein-rich nutrition effective on postoperative outcome in non-small cell lung cancer surgery? A prospective randomized study. *J Cardiothorac Surg*. Jan 19 2016;11:14. doi:10.1186/s13019-016-0407-1
32. Kalarchian MA, Marcus MD, Courcoulas AP, Cheng Y, Levine MD. Preoperative lifestyle intervention in bariatric surgery: a randomized clinical trial. *Surg Obes Relat Dis*. Jan 2016;12(1):180-7. doi:10.1016/j.soard.2015.05.004
33. Ruiz-Tovar J, Zubiaga L, Diez M, et al. Preoperative Regular Diet of 900 kcal/day vs Balanced Energy High-Protein Formula vs Immunonutrition Formula: Effect on Preoperative Weight Loss and Postoperative Pain, Complications and Analytical Acute Phase Reactants After Laparoscopic Sleeve Gastrectomy. *Obes Surg*. Jun 2016;26(6):1221-7. doi:10.1007/s11695-015-1880-7
34. Kabata P, Jastrzebski T, Kakol M, et al. Preoperative nutritional support in cancer patients with no clinical signs of malnutrition--prospective randomized controlled trial. *Support Care Cancer*. Feb 2015;23(2):365-70. doi:10.1007/s00520-014-2363-4
35. Bergin C, Speroni KG, Travis T, et al. Effect of preoperative incentive spirometry patient education on patient outcomes in the knee and hip joint replacement population. *J Perianesth Nurs*. Feb 2014;29(1):20-7. doi:10.1016/j.jopan.2013.01.009
36. Falewee MN, Schilf A, Boufflers E, et al. Reduced infections with perioperative immunonutrition in head and neck cancer: exploratory results of a multicenter, prospective, randomized, double-blind study. *Clin Nutr*. Oct 2014;33(5):776-84. doi:10.1016/j.clnu.2013.10.006
37. Oosting E, Jans MP, Dronkers JJ, et al. Preoperative home-based physical therapy versus usual care to improve functional health of frail older adults scheduled for elective total hip arthroplasty: a pilot randomized controlled trial. *Arch Phys Med Rehabil*. Apr 2012;93(4):610-6. doi:10.1016/j.apmr.2011.11.006
38. Pehlivan E, Turna A, Gurses A, Gurses HN. The effects of preoperative short-term intense physical therapy in lung cancer patients: a randomized controlled trial. *Ann Thorac Cardiovasc Surg*. 2011;17(5):461-8. doi:10.5761/atcs.oa.11.01663
39. Burden ST, Hill J, Shaffer JL, Campbell M, Todd C. An unblinded randomised controlled trial of preoperative oral supplements in colorectal cancer patients. *J Hum Nutr Diet*. Oct 2011;24(5):441-8. doi:10.1111/j.1365-277X.2011.01188.x
40. Benzo R, Wigle D, Novotny P, et al. Preoperative pulmonary rehabilitation before lung cancer resection: results from two randomized studies. *Lung Cancer*. Dec 2011;74(3):441-5. doi:10.1016/j.lungcan.2011.05.011
41. Bitterli R, Sieben JM, Hartmann M, de Bruin ED. Pre-surgical sensorimotor training for patients undergoing total hip replacement: a randomised controlled trial. *Int J Sports Med*. Sep 2011;32(9):725-32. doi:10.1055/s-0031-1271696
42. Rosenfeldt F, Braun L, Spitzer O, et al. Physical conditioning and mental stress reduction--a randomised trial in patients undergoing cardiac surgery. *BMC Complement Altern Med*. Mar 9 2011;11:20. doi:10.1186/1472-6882-11-20
43. Ishikawa Y, Yoshida H, Mamada Y, et al. Prospective randomized controlled study of short-term perioperative oral nutrition with branched chain amino acids in patients undergoing liver surgery. *Hepatogastroenterology*. May-Jun 2010;57(99-100):583-90.
44. Kulkarni SR, Fletcher E, McConnell AK, Poskitt KR, Whyman MR. Pre-operative inspiratory muscle training preserves postoperative inspiratory muscle strength following major abdominal surgery - a randomised pilot study. *Ann R Coll Surg Engl*. Nov 2010;92(8):700-7. doi:10.1308/003588410X12771863936648
45. Hoogeboom TJ, Dronkers JJ, van den Ende CH, Oosting E, van Meeteren NL. Preoperative therapeutic exercise in frail elderly scheduled for total hip replacement: a randomized pilot trial. *Clin Rehabil*. Oct 2010;24(10):901-10. doi:10.1177/0269215510371427
46. Okamoto Y, Okano K, Izuishi K, Usuki H, Wakabayashi H, Suzuki Y. Attenuation of the systemic inflammatory response and infectious complications after gastrectomy with preoperative oral arginine and omega-3 fatty acids supplemented immunonutrition. *World J Surg*. Sep 2009;33(9):1815-21. doi:10.1007/s00268-009-0140-1
47. Ferreira PE, Rodrigues AJ, Evora PR. Effects of an inspiratory muscle rehabilitation program in the postoperative period of cardiac surgery. *Arq Bras Cardiol*. Apr 2009;92(4):275-82. doi:10.1590/s0066-782x2009000400005
48. Furze G, Dumville JC, Miles JN, Irvine K, Thompson DR, Lewin RJ. "Prehabilitation" prior to CABG surgery improves physical functioning and depression. *Int J Cardiol*. Feb 6 2009;132(1):51-8. doi:10.1016/j.ijcard.2008.06.001
49. Nielsen PR, Andreasen J, Asmussen M, Tonnesen H. Costs and quality of life for prehabilitation and early rehabilitation after surgery of the lumbar spine. *BMC Health Serv Res*. Oct 9 2008;8:209. doi:10.1186/1472-6963-8-209
50. Dronkers J, Veldman A, Hoberg E, van der Waal C, van Meeteren N. Prevention of pulmonary complications after upper abdominal surgery by preoperative intensive inspiratory muscle training: a randomized controlled pilot study. *Clin Rehabil*. Feb 2008;22(2):134-42. doi:10.1177/0269215507081574
51. Hulzebos EH, Helders PJ, Favie NJ, De Bie RA, Brutel de la Riviere A, Van Meeteren NL. Preoperative intensive inspiratory muscle training to prevent postoperative pulmonary complications in high-risk patients undergoing CABG surgery: a randomized clinical trial. *JAMA*. Oct 18 2006;296(15):1851-7. doi:10.1001/jama.296.15.1851
52. Rooks DS, Huang J, Bierbaum BE, et al. Effect of preoperative exercise on measures of functional status in men and women undergoing total hip and knee arthroplasty. *Arthritis Rheum*. Oct 15 2006;55(5):700-8. doi:10.1002/art.22223
53. Xu J, Zhong Y, Jing D, Wu Z. Preoperative enteral immunonutrition improves postoperative outcome in patients with gastrointestinal cancer. *World J Surg*. Jul 2006;30(7):1284-9. doi:10.1007/s00268-005-0756-8
54. Gocen Z, Sen A, Unver B, Karatosun V, Gunal I. The effect of preoperative physiotherapy and education on the outcome of total hip replacement: a prospective randomized controlled trial. *Clin Rehabil*. Jun 2004;18(4):353-8. doi:10.1191/0269215504cr758oa
55. Beaupre LA, Lier D, Davies DM, Johnston DB. The effect of a preoperative exercise and education program on functional recovery, health related quality of life, and health service utilization following primary total knee arthroplasty. *J Rheumatol*. Jun 2004;31(6):1166-73.
56. van Bokhorst-De Van Der Schueren MA, Quak JJ, von Blomberg-van der Flier BM, et al. Effect of perioperative nutrition, with and without arginine supplementation, on nutritional status, immune function, postoperative morbidity, and survival in severely malnourished head and neck cancer patients. *Am J Clin Nutr*. Feb 2001;73(2):323-32. doi:10.1093/ajcn/73.2.323
57. Braga M, Gianotti L, Radaelli G, et al. Perioperative immunonutrition in patients undergoing cancer surgery: results of a randomized double-blind phase 3 trial. *Arch Surg*. Apr 1999;134(4):428-33. doi:10.1001/archsurg.134.4.428
58. Roukema JA, Carol EJ, Prins JG. The prevention of pulmonary complications after upper abdominal surgery in patients with noncompromised pulmonary status. *Arch Surg*. Jan 1988;123(1):30-4. doi:10.1001/archsurg.1988.01400250032004
59. Argunova Y, Korotkevich A, Pomeshkina s, Kokov A, Inozemtseva A, Barbarash O. Efficacy of physical trainings as cardioprotection method for coronary bypass surgery. *Russian Journal of Cardiology*. 07/11 2018;23:159-165. doi:10.15829/1560-4071-2018-6-159-165
60. Fulop A, Lakatos L, Susztak N, Szijarto A, Banky B. The effect of trimodal prehabilitation on the physical and psychological health of patients undergoing colorectal surgery: a randomised clinical trial. *Anaesthesia*. Jan 2021;76(1):82-90. doi:10.1111/anae.15215
61. Da Cunha F, Ruas G, Fanan JMV, et al. Effects of preoperative respiratory muscle training on early and late postoperative outcome of patients undergoing esophageal surgery. Intensive Care Medicine. 2013;39:S369.
62. Moya P, Soriano-Irigaray L, Ramirez JM, et al. Perioperative Standard Oral Nutrition Supplements Versus Immunonutrition in Patients Undergoing Colorectal Resection in an Enhanced Recovery (ERAS) Protocol: A Multicenter Randomized Clinical Trial (SONVI Study). *Medicine (Baltimore)*. May 2016;95(21):e3704. doi:10.1097/MD.0000000000003704
63. Gloor S, Misirlic M, Frei-Lanter C, et al. Prehabilitation in patients undergoing colorectal surgery fails to confer reduction in overall morbidity: results of a single-center, blinded, randomized controlled trial. *Langenbecks Arch Surg*. May 2022;407(3):897-907. doi:10.1007/s00423-022-02449-0
64. Lee SY, Lee J, Park HM, Kim CH, Kim HR. Impact of Preoperative Immunonutrition on the Outcomes of Colon Cancer Surgery: Results from a Randomized Controlled Trial. *Ann Surg*. Mar 1 2023;277(3):381-386. doi:10.1097/SLA.0000000000005140
65. Berkel AEM, Bongers BC, Kotte H, et al. Effects of Community-based Exercise Prehabilitation for Patients Scheduled for Colorectal Surgery With High Risk for Postoperative Complications: Results of a Randomized Clinical Trial. *Ann Surg*. Feb 1 2022;275(2):e299-e306. doi:10.1097/SLA.0000000000004702
66. Bhattacharyya A, Ramamoorthy L, Pottakkat B. Effect of Pre-operative Nutritional Protocol Implementation on Postoperative Outcomes Following Gastrointestinal Surgeries: A Randomized Clinical Trial. *J Caring Sci*. Nov 2021;10(4):177-183. doi:10.34172/jcs.2021.030
67. Steffens D, Young J, Beckenkamp PR, et al. Feasibility and acceptability of a preoperative exercise program for patients undergoing major cancer surgery: results from a pilot randomized controlled trial. *Pilot Feasibility Stud*. Jan 13 2021;7(1):27. doi:10.1186/s40814-021-00765-8
68. Lopez-Rodriguez-Arias F, Sanchez-Guillen L, Aranaz-Ostariz V, et al. Effect of home-based prehabilitation in an enhanced recovery after surgery program for patients undergoing colorectal cancer surgery during the COVID-19 pandemic. *Support Care Cancer*. Dec 2021;29(12):7785-7791. doi:10.1007/s00520-021-06343-1
69. Sittitrai P, Ruenmarkkaew D, Booyaprapa S, Kasempitakpong B. Effect of a perioperative immune-enhancing diet in clean-contaminated head and neck cancer surgery: A randomized controlled trial. *Int J Surg*. Sep 2021;93:106051. doi:10.1016/j.ijsu.2021.106051
70. VE IJ-H, Wanten GJA, de Nes LCF, van den Berg MGA. Effect of a Preoperative Home-Delivered, Protein-Rich Meal Service to Improve Protein Intake in Surgical Patients: A Randomized Controlled Trial. *JPEN J Parenter Enteral Nutr*. Mar 2021;45(3):479-489. doi:10.1002/jpen.2015
71. Chen ZH, Lin SY, Dai QB, Hua J, Chen SQ. The Effects of Pre-Operative Enteral Nutrition from Nasal Feeding Tubes on Gastric Outlet Obstruction. *Nutrients*. Apr 10 2017;9(4)doi:10.3390/nu9040373
72. Sawatzky JA, Kehler DS, Ready AE, et al. Prehabilitation program for elective coronary artery bypass graft surgery patients: a pilot randomized controlled study. *Clin Rehabil*. Jul 2014;28(7):648-57. doi:10.1177/0269215513516475
73. Soares SM, Nucci LB, da Silva MM, Campacci TC. Pulmonary function and physical performance outcomes with preoperative physical therapy in upper abdominal surgery: a randomized controlled trial. *Clin Rehabil*. Jul 2013;27(7):616-27. doi:10.1177/0269215512471063
74. Gillis C, Loiselle SE, Fiore JF, Jr., et al. Prehabilitation with Whey Protein Supplementation on Perioperative Functional Exercise Capacity in Patients Undergoing Colorectal Resection for Cancer: A Pilot Double-Blinded Randomized Placebo-Controlled Trial. *J Acad Nutr Diet*. May 2016;116(5):802-12. doi:10.1016/j.jand.2015.06.007
75. Carli F, Bousquet-Dion G, Awasthi R, et al. Effect of Multimodal Prehabilitation vs Postoperative Rehabilitation on 30-Day Postoperative Complications for Frail Patients Undergoing Resection of Colorectal Cancer: A Randomized Clinical Trial. *JAMA Surg*. Mar 1 2020;155(3):233-242. doi:10.1001/jamasurg.2019.5474
76. Van Nieuwenhove Y, Dambrauskas Z, Campillo-Soto A, et al. Preoperative very low-calorie diet and operative outcome after laparoscopic gastric bypass: a randomized multicenter study. *Arch Surg*. Nov 2011;146(11):1300-5. doi:10.1001/archsurg.2011.273
77. Minnella EM, Awasthi R, Loiselle SE, Agnihotram RV, Ferri LE, Carli F. Effect of Exercise and Nutrition Prehabilitation on Functional Capacity in Esophagogastric Cancer Surgery: A Randomized Clinical Trial. *JAMA Surg*. Dec 1 2018;153(12):1081-1089. doi:10.1001/jamasurg.2018.1645
78. Liang MK, Bernardi K, Holihan JL, et al. Modifying Risks in Ventral Hernia Patients With Prehabilitation: A Randomized Controlled Trial. *Ann Surg*. Oct 2018;268(4):674-680. doi:10.1097/SLA.0000000000002961
79. Barth RJ, Jr., Mills JB, Suriawinata AA, et al. Short-term Preoperative Diet Decreases Bleeding After Partial Hepatectomy: Results From a Multi-institutional Randomized Controlled Trial. *Ann Surg*. Jan 2019;269(1):48-52. doi:10.1097/SLA.0000000000002709
80. Gillis C, Li C, Lee L, et al. Prehabilitation versus rehabilitation: a randomized control trial in patients undergoing colorectal resection for cancer. *Anesthesiology*. Nov 2014;121(5):937-47. doi:10.1097/ALN.0000000000000393
81. Humeidan ML, Reyes JC, Mavarez-Martinez A, et al. Effect of Cognitive Prehabilitation on the Incidence of Postoperative Delirium Among Older Adults Undergoing Major Noncardiac Surgery: The Neurobics Randomized Clinical Trial. *JAMA Surg*. Feb 1 2021;156(2):148-156. doi:10.1001/jamasurg.2020.4371
82. Bousquet-Dion G, Awasthi R, Loiselle SE, et al. Evaluation of supervised multimodal prehabilitation programme in cancer patients undergoing colorectal resection: a randomized control trial. *Acta Oncol*. Jun 2018;57(6):849-859. doi:10.1080/0284186X.2017.1423180
83. Santa Mina D, Hilton WJ, Matthew AG, et al. Prehabilitation for radical prostatectomy: A multicentre randomized controlled trial. *Surg Oncol*. Jun 2018;27(2):289-298. doi:10.1016/j.suronc.2018.05.010
84. de Luis DA, Izaola O, Garcia Alonso M, Aller R, Cabezas G, de la Fuente B. Effect of a commercial hypocaloric diet in weight loss and post surgical morbidities in obese patients with chronic arthropathy, a randomized clinical trial. *Eur Rev Med Pharmacol Sci*. Nov 2012;16(13):1814-20.
85. Mat Eil Ismail MS, Sharifudin MA, Shokri AA, Ab Rahman S. Preoperative physiotherapy and short-term functional outcomes of primary total knee arthroplasty. *Singapore Med J*. Mar 2016;57(3):138-43. doi:10.11622/smedj.2016055
86. Abdelaal G, Eldahdouh S, Abdelsamie M, Labeeb A. Effect of preoperative physical and respiratory therapy on postoperative pulmonary functions and complications after laparoscopic upper abdominal surgery in obese patients. *Egyptian Journal of Chest Diseases and Tuberculosis*. 10/01 2017;66doi:10.1016/j.ejcdt.2017.10.012
87. D'Lima DD, Colwell CW, Jr., Morris BA, Hardwick ME, Kozin F. The effect of preoperative exercise on total knee replacement outcomes. *Clin Orthop Relat Res*. May 1996;(326):174-82. doi:10.1097/00003086-199605000-00020
88. Yeğen S, Kafadar MT, Gok M. Comparison of Perioperative Standard and Immunomodulating Enteral Nutrition in Patients Received Major Abdominal Cancer Surgery: a Prospective, Randomized, Controlled Clinical Trial. *Indian Journal of Surgery*. 03/25 2020;82doi:10.1007/s12262-020-02114-0
89. Lai Y, Su J, Yang M, Zhou K, Che G. [Impact and Effect of Preoperative Short-term Pulmonary Rehabilitation Training on Lung Cancer Patients with Mild to Moderate Chronic Obstructive Pulmonary Disease: A Randomized Trial]. *Zhongguo Fei Ai Za Zhi*. Nov 20 2016;19(11):746-753. doi:10.3779/j.issn.1009-3419.2016.11.05
90. Gade J, Levring T, Hillingso J, Hansen CP, Andersen JR. The Effect of Preoperative Oral Immunonutrition on Complications and Length of Hospital Stay After Elective Surgery for Pancreatic Cancer--A Randomized Controlled Trial. *Nutr Cancer*. 2016;68(2):225-33. doi:10.1080/01635581.2016.1142586
91. Dronkers JJ, Lamberts H, Reutelingsperger IM, et al. Preoperative therapeutic programme for elderly patients scheduled for elective abdominal oncological surgery: a randomized controlled pilot study. *Clin Rehabil*. Jul 2010;24(7):614-22. doi:10.1177/0269215509358941
92. Sorensen D, McCarthy M, Baumgartner B, Demars S. Perioperative immunonutrition in head and neck cancer. *Laryngoscope*. Jul 2009;119(7):1358-64. doi:10.1002/lary.20494
93. Hulzebos EH, van Meeteren NL, van den Buijs BJ, de Bie RA, Brutel de la Riviere A, Helders PJ. Feasibility of preoperative inspiratory muscle training in patients undergoing coronary artery bypass surgery with a high risk of postoperative pulmonary complications: a randomized controlled pilot study. *Clin Rehabil*. Nov 2006;20(11):949-59. doi:10.1177/0269215506070691
94. Allen SK, Brown V, White D, et al. Multimodal Prehabilitation During Neoadjuvant Therapy Prior to Esophagogastric Cancer Resection: Effect on Cardiopulmonary Exercise Test Performance, Muscle Mass and Quality of Life-A Pilot Randomized Clinical Trial. *Ann Surg Oncol*. Mar 2022;29(3):1839-1850. doi:10.1245/s10434-021-11002-0
95. Diaz-Feijoo B, Agusti-Garcia N, Sebio R, et al. Feasibility of a Multimodal Prehabilitation Programme in Patients Undergoing Cytoreductive Surgery for Advanced Ovarian Cancer: A Pilot Study. *Cancers (Basel)*. Mar 23 2022;14(7)doi:10.3390/cancers14071635
96. Labuschagne R, Roos R. Pre-operative physiotherapy for elderly patients undergoing abdominal surgery. *S Afr J Physiother*. 2022;78(1):1782. doi:10.4102/sajp.v78i1.1782
97. McIsaac DI, Hladkowicz E, Bryson GL, et al. Home-based prehabilitation with exercise to improve postoperative recovery for older adults with frailty having cancer surgery: the PREHAB randomised clinical trial. *Br J Anaesth*. Jul 2022;129(1):41-48. doi:10.1016/j.bja.2022.04.006
98. Sykes KJ, Gibbs H, Farrokhian N, et al. Pilot randomized, controlled, preoperative intervention for nutrition trial in head and neck cancer. *Head Neck*. Jan 2023;45(1):156-166. doi:10.1002/hed.27220
99. Tesar M, Kozusnikova V, Martinek L, Durdik S, Ihnat P. Preoperative nutritional support for patients undergoing elective colorectal cancer surgery - does it really work? *Biomed Pap Med Fac Univ Palacky Olomouc Czech Repub*. Jun 2023;167(2):145-151. doi:10.5507/bp.2022.009
100. Xia X, Ding G, Shi L, Wang M, Tian J. Effects of preoperative walking on bowel function recovery for patients undergoing gynecological malignancy laparoscopy. *Precision Medical Sciences*. 07/19 2022;11doi:10.1002/prm2.12071
101. Atoui S, Carli F, Bernard P, et al. Does a multimodal prehabilitation program improve sleep quality and duration in patients undergoing colorectal resection for cancer? Pilot randomized control trial. *J Behav Med*. Feb 2024;47(1):43-61. doi:10.1007/s10865-023-00437-3
102. Chen B, Yan X, Wang X, Mao Y. Effectiveness of precise and quantitative rapid pulmonary rehabilitation nursing program for elderly patients with lung cancer during the perioperative period: A randomized controlled trial. *Pak J Med Sci*. Mar-Apr 2023;39(2):572-577. doi:10.12669/pjms.39.2.7103
103. Molenaar CJL, Minnella EM, Coca-Martinez M, et al. Effect of Multimodal Prehabilitation on Reducing Postoperative Complications and Enhancing Functional Capacity Following Colorectal Cancer Surgery: The PREHAB Randomized Clinical Trial. *JAMA Surg*. Jun 1 2023;158(6):572-581. doi:10.1001/jamasurg.2023.0198
104. Chen J, Luo AL, Yang L, Wang W, Zhou X, Yang M. Nutrition management by a multidisciplinary team for prevention of nutritional deficits and morbidity following esophagectomy. *Braz J Med Biol Res*. 2023;56:e12421. doi:10.1590/1414-431X2023e12421
105. Zhang J, Sun W, Wang T, Li J, Yu M. Effect of preoperative functional exercise on cephalic vein diameter, anastomotic blood flow, and postoperative complications in patients with arteriovenous internal fistuloplasty. *J Pak Med Assoc*. Mar 2023;73(3):653-655. doi:10.47391/JPMA.6224
106. Wall C, Glyn T, Rowbotham D, et al. P769 Randomised feasibility study of preoperative medical nutrition therapy in adults undergoing surgery for Crohn’s disease. *Journal of Crohn's and Colitis*. 2023;17(Supplement_1):i899-i899. doi:10.1093/ecco-jcc/jjac190.0899
107. Vind AB, Andersen HE, Pedersen KD, Jorgensen T, Schwarz P. An outpatient multifactorial falls prevention intervention does not reduce falls in high-risk elderly Danes. *J Am Geriatr Soc*. Jun 2009;57(6):971-7. doi:10.1111/j.1532-5415.2009.02270.x
108. Sakamoto K, Endo N, Harada A, et al. Why not use your own body weight to prevent falls? A randomized, controlled trial of balance therapy to prevent falls and fractures for elderly people who can stand on one leg for </=15 s. *J Orthop Sci*. Jan 2013;18(1):110-20. doi:10.1007/s00776-012-0328-3
109. Sambrook PN, Cameron ID, Chen JS, et al. Does increased sunlight exposure work as a strategy to improve vitamin D status in the elderly: a cluster randomised controlled trial. *Osteoporos Int*. Feb 2012;23(2):615-24. doi:10.1007/s00198-011-1590-5
110. Perula LA, Varas-Fabra F, Rodriguez V, et al. Effectiveness of a multifactorial intervention program to reduce falls incidence among community-living older adults: a randomized controlled trial. *Arch Phys Med Rehabil*. Oct 2012;93(10):1677-84. doi:10.1016/j.apmr.2012.03.035
111. Glendenning P, Zhu K, Inderjeeth C, Howat P, Lewis JR, Prince RL. Effects of three-monthly oral 150,000 IU cholecalciferol supplementation on falls, mobility, and muscle strength in older postmenopausal women: a randomized controlled trial. *J Bone Miner Res*. Jan 2012;27(1):170-6. doi:10.1002/jbmr.524
112. Pekkarinen T, Loyttyniemi E, Valimaki M. Hip fracture prevention with a multifactorial educational program in elderly community-dwelling Finnish women. *Osteoporos Int*. Dec 2013;24(12):2983-92. doi:10.1007/s00198-013-2381-y
113. Neelemaat F, Lips P, Bosmans JE, Thijs A, Seidell JC, van Bokhorst-de van der Schueren MA. Short-term oral nutritional intervention with protein and vitamin D decreases falls in malnourished older adults. *J Am Geriatr Soc*. Apr 2012;60(4):691-9. doi:10.1111/j.1532-5415.2011.03888.x
114. Palvanen M, Kannus P, Piirtola M, Niemi S, Parkkari J, Jarvinen M. Effectiveness of the Chaos Falls Clinic in preventing falls and injuries of home-dwelling older adults: a randomised controlled trial. *Injury*. Jan 2014;45(1):265-71. doi:10.1016/j.injury.2013.03.010
115. Suman S, Myint PK, Clark A, Das P, Ring L, Trepte NJ. Community-based fall assessment compared with hospital-based assessment in community-dwelling older people over 65 at high risk of falling: a randomized study. *Aging Clin Exp Res*. Feb 2011;23(1):35-41. doi:10.1007/BF03324951
116. Dangour AD, Albala C, Allen E, et al. Effect of a nutrition supplement and physical activity program on pneumonia and walking capacity in Chilean older people: a factorial cluster randomized trial. *PLoS Med*. Apr 2011;8(4):e1001023. doi:10.1371/journal.pmed.1001023
117. Spink MJ, Menz HB, Fotoohabadi MR, et al. Effectiveness of a multifaceted podiatry intervention to prevent falls in community dwelling older people with disabling foot pain: randomised controlled trial. *BMJ*. Jun 16 2011;342:d3411. doi:10.1136/bmj.d3411
118. Cameron ID, Kurrle S, Quine S, et al. Increasing adherence with the use of hip protectors for older people living in the community. *Osteoporos Int*. Feb 2011;22(2):617-26. doi:10.1007/s00198-010-1334-y
119. Haines TP, Hill AM, Hill KD, et al. Patient education to prevent falls among older hospital inpatients: a randomized controlled trial. *Arch Intern Med*. Mar 28 2011;171(6):516-24. doi:10.1001/archinternmed.2010.444
120. Drahota AK, Ward D, Udell JE, et al. Pilot cluster randomised controlled trial of flooring to reduce injuries from falls in wards for older people. *Age Ageing*. Sep 2013;42(5):633-40. doi:10.1093/ageing/aft067
121. Sanders KM, Stuart AL, Williamson EJ, et al. Annual high-dose oral vitamin D and falls and fractures in older women: a randomized controlled trial. *JAMA*. May 12 2010;303(18):1815-22. doi:10.1001/jama.2010.594
122. Iliffe S, Kendrick D, Morris R, et al. Multicentre cluster randomised trial comparing a community group exercise programme and home-based exercise with usual care for people aged 65 years and over in primary care. *Health Technol Assess*. Aug 2014;18(49):vii-xxvii, 1-105. doi:10.3310/hta18490
123. de Vries OJ, Peeters GM, Elders PJ, et al. Multifactorial intervention to reduce falls in older people at high risk of recurrent falls: a randomized controlled trial. *Arch Intern Med*. Jul 12 2010;170(13):1110-7. doi:10.1001/archinternmed.2010.169
124. Meyer G, Kopke S, Haastert B, Muhlhauser I. Comparison of a fall risk assessment tool with nurses' judgement alone: a cluster-randomised controlled trial. *Age Ageing*. Jul 2009;38(4):417-23. doi:10.1093/ageing/afp049
125. Koike T, Orito Y, Toyoda H, et al. External hip protectors are effective for the elderly with higher-than-average risk factors for hip fractures. *Osteoporos Int*. Sep 2009;20(9):1613-20. doi:10.1007/s00198-008-0824-7
126. Salminen M, Vahlberg T, Kivela SL. The long-term effect of a multifactorial fall prevention programme on the incidence of falls requiring medical treatment. *Public Health*. Dec 2009;123(12):809-13. doi:10.1016/j.puhe.2009.10.018
127. Spice CL, Morotti W, George S, et al. The Winchester falls project: a randomised controlled trial of secondary prevention of falls in older people. *Age Ageing*. Jan 2009;38(1):33-40. doi:10.1093/ageing/afn192
128. Rosendahl E, Gustafson Y, Nordin E, Lundin-Olsson L, Nyberg L. A randomized controlled trial of fall prevention by a high-intensity functional exercise program for older people living in residential care facilities. *Aging Clin Exp Res*. Feb 2008;20(1):67-75. doi:10.1007/BF03324750
129. Cumming RG, Sherrington C, Lord SR, et al. Cluster randomised trial of a targeted multifactorial intervention to prevent falls among older people in hospital. *BMJ*. Apr 5 2008;336(7647):758-60. doi:10.1136/bmj.39499.546030.BE
130. Prince RL, Austin N, Devine A, Dick IM, Bruce D, Zhu K. Effects of ergocalciferol added to calcium on the risk of falls in elderly high-risk women. *Arch Intern Med*. Jan 14 2008;168(1):103-8. doi:10.1001/archinternmed.2007.31
131. Berggren M, Stenvall M, Olofsson B, Gustafson Y. Evaluation of a fall-prevention program in older people after femoral neck fracture: a one-year follow-up. *Osteoporos Int*. Jun 2008;19(6):801-9. doi:10.1007/s00198-007-0507-9
132. Burleigh E, McColl J, Potter J. Does vitamin D stop inpatients falling? A randomised controlled trial. *Age Ageing*. Sep 2007;36(5):507-13. doi:10.1093/ageing/afm087
133. Smith H, Anderson F, Raphael H, Maslin P, Crozier S, Cooper C. Effect of annual intramuscular vitamin D on fracture risk in elderly men and women--a population-based, randomized, double-blind, placebo-controlled trial. *Rheumatology (Oxford)*. Dec 2007;46(12):1852-7. doi:10.1093/rheumatology/kem240
134. Cumming RG, Ivers R, Clemson L, et al. Improving vision to prevent falls in frail older people: a randomized trial. *J Am Geriatr Soc*. Feb 2007;55(2):175-81. doi:10.1111/j.1532-5415.2007.01046.x
135. Korpelainen R, Keinanen-Kiukaanniemi S, Heikkinen J, Vaananen K, Korpelainen J. Effect of impact exercise on bone mineral density in elderly women with low BMD: a population-based randomized controlled 30-month intervention. *Osteoporos Int*. Jan 2006;17(1):109-18. doi:10.1007/s00198-005-1924-2
136. Foss AJ, Harwood RH, Osborn F, Gregson RM, Zaman A, Masud T. Falls and health status in elderly women following second eye cataract surgery: a randomised controlled trial. *Age Ageing*. Jan 2006;35(1):66-71. doi:10.1093/ageing/afj005
137. Law M, Withers H, Morris J, Anderson F. Vitamin D supplementation and the prevention of fractures and falls: results of a randomised trial in elderly people in residential accommodation. *Age Ageing*. Sep 2006;35(5):482-6. doi:10.1093/ageing/afj080
138. Harwood RH, Foss AJ, Osborn F, Gregson RM, Zaman A, Masud T. Falls and health status in elderly women following first eye cataract surgery: a randomised controlled trial. *Br J Ophthalmol*. Jan 2005;89(1):53-9. doi:10.1136/bjo.2004.049478
139. Grant AM, Avenell A, Campbell MK, et al. Oral vitamin D3 and calcium for secondary prevention of low-trauma fractures in elderly people (Randomised Evaluation of Calcium Or vitamin D, RECORD): a randomised placebo-controlled trial. *Lancet*. May 7-13 2005;365(9471):1621-8. doi:10.1016/S0140-6736(05)63013-9
140. Davison J, Bond J, Dawson P, Steen IN, Kenny RA. Patients with recurrent falls attending Accident & Emergency benefit from multifactorial intervention--a randomised controlled trial. *Age Ageing*. Mar 2005;34(2):162-8. doi:10.1093/ageing/afi053
141. Ray WA, Taylor JA, Brown AK, et al. Prevention of fall-related injuries in long-term care: a randomized controlled trial of staff education. *Arch Intern Med*. Oct 24 2005;165(19):2293-8. doi:10.1001/archinte.165.19.2293
142. Barr RJ, Stewart A, Torgerson DJ, Seymour DG, Reid DM. Screening elderly women for risk of future fractures--participation rates and impact on incidence of falls and fractures. *Calcif Tissue Int*. Apr 2005;76(4):243-8. doi:10.1007/s00223-004-0101-5
143. Flicker L, MacInnis RJ, Stein MS, et al. Should older people in residential care receive vitamin D to prevent falls? Results of a randomized trial. *J Am Geriatr Soc*. Nov 2005;53(11):1881-8. doi:10.1111/j.1532-5415.2005.00468.x
144. Sato Y, Kanoko T, Satoh K, Iwamoto J. The prevention of hip fracture with risedronate and ergocalciferol plus calcium supplementation in elderly women with Alzheimer disease: a randomized controlled trial. *Arch Intern Med*. Aug 8-22 2005;165(15):1737-42. doi:10.1001/archinte.165.15.1737
145. Dyer CA, Taylor GJ, Reed M, Dyer CA, Robertson DR, Harrington R. Falls prevention in residential care homes: a randomised controlled trial. *Age Ageing*. Nov 2004;33(6):596-602. doi:10.1093/ageing/afh204
146. Cameron ID, Cumming RG, Kurrle SE, et al. A randomised trial of hip protector use by frail older women living in their own homes. *Inj Prev*. Jun 2003;9(2):138-41. doi:10.1136/ip.9.2.138
147. Becker C, Kron M, Lindemann U, et al. Effectiveness of a multifaceted intervention on falls in nursing home residents. *J Am Geriatr Soc*. Mar 2003;51(3):306-13. doi:10.1046/j.1532-5415.2003.51103.x
148. Nikolaus T, Bach M. Preventing falls in community-dwelling frail older people using a home intervention team (HIT): results from the randomized Falls-HIT trial. *J Am Geriatr Soc*. Mar 2003;51(3):300-5. doi:10.1046/j.1532-5415.2003.51102.x
149. Birks YF, Hildreth R, Campbell P, Sharpe C, Torgerson DJ, Watt I. Randomised controlled trial of hip protectors for the prevention of second hip fractures. *Age Ageing*. Jul 2003;32(4):442-4. doi:10.1093/ageing/32.4.442
150. Hogan DB, MacDonald FA, Betts J, et al. A randomized controlled trial of a community-based consultation service to prevent falls. *CMAJ*. Sep 4 2001;165(5):537-43.
151. Kenny RA, Richardson DA, Steen N, Bexton RS, Shaw FE, Bond J. Carotid sinus syndrome: a modifiable risk factor for nonaccidental falls in older adults (SAFE PACE). *J Am Coll Cardiol*. Nov 1 2001;38(5):1491-6. doi:10.1016/s0735-1097(01)01537-6
152. Cameron ID, Venman J, Kurrle SE, et al. Hip protectors in aged-care facilities: a randomized trial of use by individual higher-risk residents. *Age Ageing*. Nov 2001;30(6):477-81. doi:10.1093/ageing/30.6.477
153. McMurdo ME, Millar AM, Daly F. A randomized controlled trial of fall prevention strategies in old peoples' homes. *Gerontology*. Mar-Apr 2000;46(2):83-7. doi:10.1159/000022139
154. Fairhall N, Sherrington C, Lord SR, et al. Effect of a multifactorial, interdisciplinary intervention on risk factors for falls and fall rate in frail older people: a randomised controlled trial. *Age Ageing*. Sep 2014;43(5):616-22. doi:10.1093/ageing/aft204
155. Gianoudis J, Bailey CA, Ebeling PR, et al. Effects of a targeted multimodal exercise program incorporating high-speed power training on falls and fracture risk factors in older adults: a community-based randomized controlled trial. *J Bone Miner Res*. Jan 2014;29(1):182-91. doi:10.1002/jbmr.2014
156. Kim H, Yoshida H, Suzuki T. Falls and fractures in participants and excluded non-participants of a fall prevention exercise program for elderly women with a history of falls: 1-year follow-up study. *Geriatr Gerontol Int*. Apr 2014;14(2):285-92. doi:10.1111/ggi.12095
157. Pitkala KH, Poysti MM, Laakkonen ML, et al. Effects of the Finnish Alzheimer disease exercise trial (FINALEX): a randomized controlled trial. *JAMA Intern Med*. May 27 2013;173(10):894-901. doi:10.1001/jamainternmed.2013.359
158. Sherrington C, Lord SR, Vogler CM, et al. A post-hospital home exercise program improved mobility but increased falls in older people: a randomised controlled trial. *PLoS One*. 2014;9(9):e104412. doi:10.1371/journal.pone.0104412
159. Sjoberg C, Wallerstedt SM. Effects of medication reviews performed by a physician on treatment with fracture-preventing and fall-risk-increasing drugs in older adults with hip fracture-a randomized controlled study. *J Am Geriatr Soc*. Sep 2013;61(9):1464-72. doi:10.1111/jgs.12412
160. Larsen ER, Mosekilde L, Foldspang A. Vitamin D and calcium supplementation prevents severe falls in elderly community-dwelling women: a pragmatic population-based 3-year intervention study. *Aging Clin Exp Res*. Apr 2005;17(2):125-32. doi:10.1007/BF03324585
161. Lips P, Graafmans WC, Ooms ME, Bezemer PD, Bouter LM. Vitamin D supplementation and fracture incidence in elderly persons. A randomized, placebo-controlled clinical trial. *Ann Intern Med*. Feb 15 1996;124(4):400-6. doi:10.7326/0003-4819-124-4-199602150-00003
162. Patil R, Uusi-Rasi K, Tokola K, Karinkanta S, Kannus P, Sievanen H. Effects of a Multimodal Exercise Program on Physical Function, Falls, and Injuries in Older Women: A 2-Year Community-Based, Randomized Controlled Trial. *J Am Geriatr Soc*. Jul 2015;63(7):1306-13. doi:10.1111/jgs.13489
163. Chapuy MC, Arlot ME, Delmas PD, Meunier PJ. Effect of calcium and cholecalciferol treatment for three years on hip fractures in elderly women. *BMJ*. Apr 23 1994;308(6936):1081-2. doi:10.1136/bmj.308.6936.1081
164. Chapuy MC, Pamphile R, Paris E, et al. Combined calcium and vitamin D3 supplementation in elderly women: confirmation of reversal of secondary hyperparathyroidism and hip fracture risk: the Decalyos II study. *Osteoporos Int*. Mar 2002;13(3):257-64. doi:10.1007/s001980200023
165. Gallagher JC, Fowler SE, Detter JR, Sherman SS. Combination treatment with estrogen and calcitriol in the prevention of age-related bone loss. *J Clin Endocrinol Metab*. Aug 2001;86(8):3618-28. doi:10.1210/jcem.86.8.7703
166. Ferrer A, Formiga F, Sanz H, et al. Multifactorial assessment and targeted intervention to reduce falls among the oldest-old: a randomized controlled trial. *Clin Interv Aging*. 2014;9:383-93. doi:10.2147/CIA.S57580
167. Pfeifer M, Begerow B, Minne HW, Abrams C, Nachtigall D, Hansen C. Effects of a short-term vitamin D and calcium supplementation on body sway and secondary hyperparathyroidism in elderly women. *J Bone Miner Res*. Jun 2000;15(6):1113-8. doi:10.1359/jbmr.2000.15.6.1113
168. Reid IR, Mason B, Horne A, et al. Randomized controlled trial of calcium in healthy older women. *Am J Med*. Sep 2006;119(9):777-85. doi:10.1016/j.amjmed.2006.02.038
169. Trivedi DP, Doll R, Khaw KT. Effect of four monthly oral vitamin D3 (cholecalciferol) supplementation on fractures and mortality in men and women living in the community: randomised double blind controlled trial. *BMJ*. Mar 1 2003;326(7387):469. doi:10.1136/bmj.326.7387.469
170. Vetter NJ, Lewis PA, Ford D. Can health visitors prevent fractures in elderly people? *BMJ*. Apr 4 1992;304(6831):888-90. doi:10.1136/bmj.304.6831.888
171. Bucknall C, Miller G, Lloyd S, et al. Glasgow supported self-management trial (GSuST) for patients with moderate to severe COPD: randomised controlled trial. *BMJ*. 2012;344
172. Chi C, Chen H. DAILY-BASED SELF-MANAGEMENT FOR NON-HOSPITALISED HEART FAILURE PATIENTS IMPROVE PROGNOSIS. *Heart*. 2012;98(Suppl 2):E231-E232.
173. Fan VS, Gaziano JM, Lew R, et al. A comprehensive care management program to prevent chronic obstructive pulmonary disease hospitalizations: a randomized, controlled trial. *Ann Intern Med*. 2012;156(10):673-683.
174. Freund T, Peters-Klimm F, Boyd CM, et al. Medical Assistant-Based Care Management for High-Risk Patients in Small Primary Care Practices: A Cluster Randomized Clinical Trial. *Ann Intern Med*. Mar 1 2016;164(5):323-30. doi:10.7326/M14-2403
175. Hernández C, Alonso A, Garcia-Aymerich J, et al. Effectiveness of community-based integrated care in frail COPD patients: a randomised controlled trial. *NPJ Prim Care Respir Med*. 2015;25(1):1-6.
176. Kennedy A, Bower P, Reeves D, et al. Implementation of self management support for long term conditions in routine primary care settings: cluster randomised controlled trial. *BMJ*. 2013;346
177. Nguyen HQ, Donesky D, Reinke LF, et al. Internet-based dyspnea self-management support for patients with chronic obstructive pulmonary disease. *J Pain Symptom Manag*. 2013;46(1):43-55.
178. Pinnock H, Hanley J, McCloughan L, et al. Effectiveness of telemonitoring integrated into existing clinical services on hospital admission for exacerbation of chronic obstructive pulmonary disease: researcher blind, multicentre, randomised controlled trial. *BMJ*. 2013;347
179. van de Ven G, Draskovic I, Adang EM, et al. Effects of dementia-care mapping on residents and staff of care homes: a pragmatic cluster-randomised controlled trial. *PloS One*. 2013;8(7):e67325.
180. Xi F, Wang Z, Qi Y, et al. Long-term effect of respiratory training for chronic obstructive pulmonary disease patients at an outpatient clinic: a randomised controlled trial. *Clin Transl Med*. 2015;4(1):1-7.
181. Bosanquet K, Adamson J, Atherton K, et al. CollAborative care for Screen-Positive EldeRs with major depression (CASPER plus): a multicentred randomised controlled trial of clinical effectiveness and cost-effectiveness. *Health Technol Assess*. 2017;21(67):1.
182. Forster A, Young J, Chapman K, et al. Cluster randomized controlled trial: clinical and cost-effectiveness of a system of longer-term stroke care. *Stroke*. 2015;46(8):2212-2219.
183. Khdour MR, Kidney JC, Smyth BM, McElnay JC. Clinical pharmacy‐led disease and medicine management programme for patients with COPD. *Br J Clin Pharmacol*. 2009;68(4):588-598.
184. Markle-Reid M, Orridge C, Weir R, et al. Interprofessional stroke rehabilitation for stroke survivors using home care. *Can J Neurol Sci*. 2011;38(2):317-334.
185. Murphy AW, Cupples M, Smith S, Byrne M, Byrne M, Newell J. Effect of tailored practice and patient care plans on secondary prevention of heart disease in general practice: cluster randomised controlled trial. *BMJ*. 2009;339
186. Ortiz-Bautista C, Diaz M, Delgado-Nicolas M, et al. Evaluation of a nurse-led cross intervention program in heart failure. WILEY 111 RIVER ST, HOBOKEN 07030-5774, NJ USA; 2017:306-307.
187. Peters-Klimm F, Campbell S, Hermann K, Kunz CU, Müller-Tasch T, Szecsenyi J. Case management for patients with chronic systolic heart failure in primary care: the HICMan exploratory randomised controlled trial. *Trials*. 2010;11(1):1-14.
188. Sackley CM, Walker MF, Burton CR, et al. An occupational therapy intervention for residents with stroke related disabilities in UK care homes (OTCH): cluster randomised controlled trial. *BMJ*. 2015;350
189. Schrader G, Cheok F, Hordacre AL, Marker J, Wade V. Effect of psychiatry liaison with general practitioners on depression severity in recently hospitalised cardiac patients: a randomised controlled trial. *Med J Aust*. 2005;182(6):272-276.
190. van der Aa HP, van Rens GH, Comijs HC, et al. Stepped care for depression and anxiety in visually impaired older adults: multicentre randomised controlled trial. *BMJ*. 2015;351
191. Xiao LD, De Bellis A, Kyriazopoulos H, Draper B, Ullah S. The Effect of a Personalized Dementia Care Intervention for Caregivers From Australian Minority Groups. *Am J Alzheimers Dis Other Demen*. Feb 2016;31(1):57-67. doi:10.1177/1533317515578256
192. Zwar NA, Bunker JM, Reddel HK, et al. Early intervention for chronic obstructive pulmonary disease by practice nurse and GP teams: a cluster randomized trial. *Fam Pract*. Dec 2016;33(6):663-670. doi:10.1093/fampra/cmw077
193. Schäfer I, Kaduszkiewicz H, Mellert C, et al. Narrative medicine-based intervention in primary care to reduce polypharmacy: results from the cluster-randomised controlled trial MultiCare AGENDA. *BMJ Open*. 2018;8(1):e017653.
194. Vaillant-Roussel H, Laporte C, Pereira B, et al. Impact of patient education on chronic heart failure in primary care (ETIC): a cluster randomised trial. *BMC Fam Pract*. 2016;17(1):1-13.
195. Stewart S, Ball J, Horowitz JD, et al. Standard versus atrial fibrillation-specific management strategy (SAFETY) to reduce recurrent admission and prolong survival: pragmatic, multicentre, randomised controlled trial. *Lancet*. 2015;385(9970):775-784.
196. Walters J, Cameron-Tucker H, Wills K, et al. Effects of telephone health mentoring in community-recruited chronic obstructive pulmonary disease on self-management capacity, quality of life and psychological morbidity: a randomised controlled trial. *Bmj Open*. 2013;3(9):e003097.
197. Wagenaar KP, Broekhuizen BD, Jaarsma T, et al. Effectiveness of the European Society of Cardiology/Heart Failure Association website ‘heartfailurematters. org’and an e‐health adjusted care pathway in patients with stable heart failure: results of the ‘e‐Vita HF’randomized controlled trial. *European Journal of Heart Failure*. 2019;21(2):238-246.
198. Liu WT, Wang CH, Lin HC, et al. Efficacy of a cell phone-based exercise programme for COPD. *Eur Respir J*. Sep 2008;32(3):651-9. doi:10.1183/09031936.00104407
199. McManus RJ, Mant J, Haque MS, et al. Effect of self-monitoring and medication self-titration on systolic blood pressure in hypertensive patients at high risk of cardiovascular disease: the TASMIN-SR randomized clinical trial. *Jama*. 2014;312(8):799-808.
200. Galbreath AD, Krasuski RA, Smith B, et al. Long-term healthcare and cost outcomes of disease management in a large, randomized, community-based population with heart failure. *Circulation*. 2004;110(23):3518-3526.
201. de la Porte PW, Lok DJ, van Veldhuisen DJ, et al. Added value of a physician-and-nurse-directed heart failure clinic: results from the Deventer–Alkmaar heart failure study. *Heart*. 2007;93(7):819-825.
202. Eccles MP, Whitty PM, Speed C, et al. A pragmatic cluster randomised controlled trial of a Diabetes REcall And Management system: the DREAM trial. *Implementation science : IS*. 2007;2(1):1-12.
203. López Cabezas C, Falces Salvador C, Cubí Quadrada D, et al. Randomized clinical trial of a postdischarge pharmaceutical care program vs regular follow-up in patients with heart failure. *Farmacia Hospitalaria*. 2006;30(6):328-342.
204. Ojeda S, Anguita M, Delgado M, et al. Short‐and long‐term results of a programme for the prevention of readmissions and mortality in patients with heart failure: are effects maintained after stopping the programme? *Eur J Heart Fail*. 2005;7(5):921-926.
205. Veroniki AA, Soobiah C, Nincic V, et al. Efficacy of sustained knowledge translation (KT) interventions in chronic disease management in older adults: systematic review and meta-analysis of complex interventions. *BMC Med*. Jul 24 2023;21(1):269. doi:10.1186/s12916-023-02966-9
206. Deeks J, Higgins J, Altman D. Chapter 10: Analysing data and undertaking meta-analyses. . In: Higgins J, Thomas J, Chandler J, et al, eds. *Cochrane Handbook for Systematic Reviews of Interventions*. version 6.3 (updated February 2022) ed. Cochrane; 2022.

### Appendix 2. Frequency of components included in single or multiple combinations of components in a) post-operative complications, b) falls-related fractures and c) quality of life examples.

| **Components** | **1-component combination** | **2-component combinations** | **3-component combinations** | **4-component combinations** | **5-component combinations** | **6-component combinations** | **7-component combinations** | **Total*** |
| --- | --- | --- | --- | --- | --- | --- | --- | --- |
| **Post-operative complications** | | | | | | | | |
| *UC* | 107 |  |  |  |  |  |  | **107** |
| *COG* | 3 |  |  |  |  |  |  | **3** |
| *EXE* | 44 | 7 | 13 |  |  |  |  | **64** |
| *NUT* | 39 | 3 | 13 |  |  |  |  | **55** |
| *PSY* | 3 | 4 | 13 |  |  |  |  | **20** |
| **Falls-related fractures** | | | | | | | | |
| *UC* | 37 |  |  |  |  |  |  | **37** |
| *bf* | 5 | 3 | 1 | 1 |  |  |  | **10** |
| *ca* | 6 | 10 | 2 | 2 | 1 |  |  | **21** |
| *cbt* |  |  | 1 |  |  |  |  | **1** |
| *cl-qi* | 2 | 10 | 10 | 3 | 2 |  |  | **27** |
| *cp* |  |  | 1 | 1 |  |  |  | **2** |
| *de-al* | 1 |  |  |  | 1 |  |  | **2** |
| *de-hp* |  | 5 |  | 1 |  |  |  | **6** |
| *de-ft* |  |  | 1 |  |  |  |  | **1** |
| *di* | 1 | 1 |  |  | 1 |  |  | **3** |
| *ea* |  | 2 | 1 | 2 |  |  |  | **5** |
| *ex* | 9 | 4 | 5 | 3 | 1 |  |  | **22** |
| *fl* | 1 |  |  |  |  |  |  | **1** |
| *mf* | 4 | 6 | 8 | 3 | 1 |  |  | **22** |
| *op-tx* | 1 | 1 | 1 |  |  |  |  | **3** |
| *pa-qi* | 13 | 6 | 9 | 5 | 2 |  |  | **35** |
| *so* | 3 |  |  | 1 |  |  |  | **4** |
| *su-ey* | 2 |  |  |  |  |  |  | **2** |
| *su-pm* |  | 1 |  |  |  |  |  | **1** |
| *va* |  | 2 |  |  |  |  |  | **1** |
| *vi-d* | 7 | 12 | 2 | 2 | 1 |  |  | **24** |
| **Quality of life** | | | | | | | | |
| *UC* | 33 |  |  |  |  |  |  | **33** |
| *AF* |  |  |  |  |  |  | 1 | **1** |
| *sta* | 1 | 3 | 2 | 3 | 3 | 2 |  | **14** |
| *clin_re* |  |  |  | 1 |  |  |  | **1** |
| *EPR* |  | 1 |  | 1 | 2 |  | 1 | **5** |
| *px_ed* | 2 | 4 | 6 | 5 | 6 | 2 | 1 | **26** |
| *px_re* |  | 1 |  | 2 | 2 |  |  | **5** |
| *te* |  | 1 | 1 | 2 | 5 | 2 | 1 | **12** |
| *c-M* |  | 5 | 4 | 4 | 7 | 2 | 1 | **23** |
| *s-M* | 1 | 9 | 6 | 5 | 6 | 2 | 1 | **30** |
| *re* |  |  | 2 | 1 | 2 |  | 1 | **6** |
| *fin* |  |  |  |  |  | 1 |  | **1** |
| *mot_int* |  |  |  |  | 2 |  |  | **2** |
| *px_fin* |  |  |  |  |  | 1 |  | **1** |

* 193 total active components and 37 UC inactive components were assessed across all studies in falls-related fractures; 127 total active components and 33 UC inactive components were assessed across all studies in quality of life.

*Abbreviations: AF*, Audit & feedback; *BF,* Basic falls risk assessment*; CA,* Calcium*; CBT,* Cognitive behavioral therapy*; CLIN_RE*, Clinician reminders*; CL-QI,* Clinic-level quality improvement*; C-M*, Case management*; COG*, Cognitive; *CP,* Comprehensive podiatry assessment and treatment*; DE-AL,* Device—alarm*; DE-FT,* Multifaceted podiatry; *DE-HP,* Device—hip protector*; DI,* Dietary modifications*; EA,* Environmental assessment and modification*; EPR*, Electronic patient registry*; EXE,* Exercise*; FIN*, Financial incentives*; FL,* Flooring*; MF,* Multifactorial assessment and treatment*; MOT_INT*, Motivational interview*; NUT*, Nutrition; *OP-TX,* Osteoporosis treatment*; PA-QI,* Patient-level quality improvement*; PSY*, Psychosocial; *PX_ED*, Patient education*; PX_FIN*, Financial incentives*; PX_RE*, Patient reminders*; RE*, Facilitated relay of info to clinicians*; SD*, Standard Deviation; *S-M*, Promotion of self-management*; SO,* Social engagement*; STA*, Staff education*; SU-EY,* Surgery—cataract*; SU-PM,* Surgery—pacemaker*; TE*, Team changes*; UC,* Usual Care*; VA,* Vision assessment and treatment*; VI-D,* Vitamin D

### Appendix 3. Additional empirical examples with complex interventions

#### Prevention of falls-related fractures

We explored the efficacy of complex interventions for the prevention of falls-related fractures in older adults using CNMA approaches.^210^ The network included binary outcome data of 68 RCTs (86,491 participants; 62 two-arm, three three-arm, three four-arm RCTs), 43 different combinations of components, including 20 individual active components, plus *UC* (inactive intervention) (**Appendix** **Table 1**, **Appendix** **Figure 1**). Data are presented in **Appendix 1**. Thirty-seven RCTs compared 28 different combinations of components with *UC* (65,904 participants; 38,061 participants in falls-prevention interventions and 27,843 in *UC*). Active combinations of components contained one to five components, with patient-level quality improvement (*pa-qi*) being the most frequent component included in the combinations of components (35 in 230 total active components were assessed across all studies, 18%) (**Appendix 2**). Overall, *PA-QI* appeared in 18 out of 43 active combinations of components (42%) and was evaluated in 24 (35%) studies.

**Appendix Table 1. Component Abbreviations in the falls-related fractures**

| **Component** | **Abbreviation** |
| --- | --- |
| Usual Care | *UC* |
| Basic falls risk assessment | *BF* |
| Calcium | *CA* |
| Cognitive behavioral therapy | *CBT* |
| Clinic-level quality improvement | *CL-QI* |
| Comprehensive podiatry assessment and treatment | *CP* |
| Device—alarm | *DE-AL* |
| Device—hip protector | *DE-HP* |
| Multifaceted podiatry | *DE-FT* |
| Dietary modifications | *DI* |
| Environmental assessment and modification | *EA* |
| Exercise | *EX* |
| Flooring | *FL* |
| Multifactorial assessment and treatment | *MF* |
| Osteoporosis treatment | *OP-TX* |
| Patient-level quality improvement | *PA-QI* |
| Social engagement | *SO* |
| Surgery—cataract | *SU-EY* |
| Surgery—pacemaker | *SU-PM* |
| Vision assessment and treatment | *VA* |
| Vitamin D | *VI-D* |

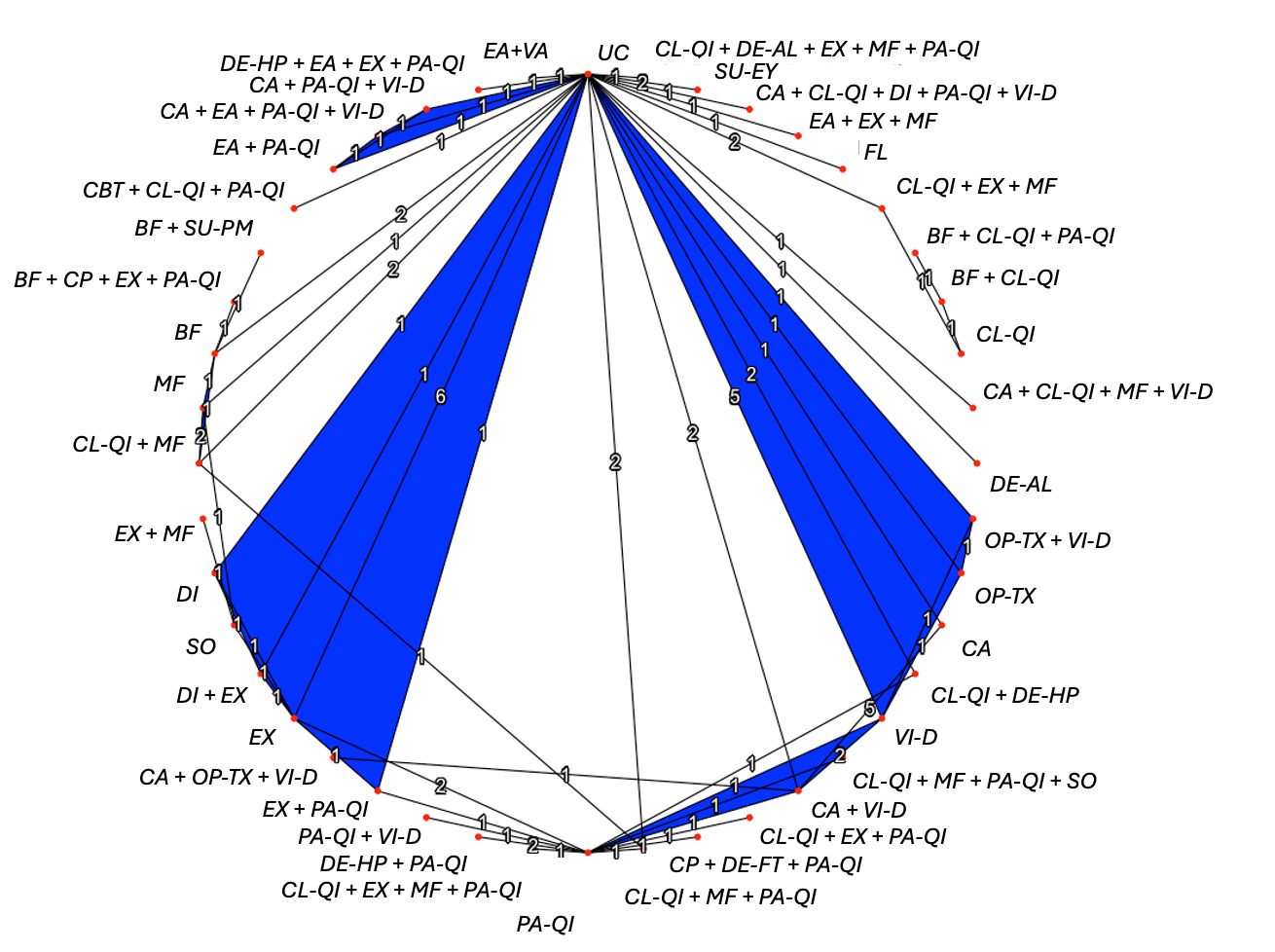

**Appendix Figure 1. Network diagram depicting network connectedness of complex interventions for falls-related fractures. Nodes represent complex interventions, as combinations of individual components. Thickness of solid lines is proportional to the number of studies included in the group comparison, and node size is proportional to the number of patients included in the underlying group.**

The design-by-treatment interaction model^211^ as an extension of the standard NMA model and assuming combinations of components as different nodes, suggested no statistical evidence for inconsistency (Q_between-designs_ = 12.40, *df* = 15, p-value = 0.648, τ^2^ = 0.22). Overall, comparing the NMA and additive CNMA effect estimates, CNMA provided more precise and more conservative effect estimates closer to null (LOR = 0). Heterogeneity in the CNMA was comparable with the standard NMA (NMA: 𝐼^2^ = 38%, τ^2^ = 0.04, Q_NMA_ = 54.62, *df* = 34, p-value = 0.014; Additive CNMA: 𝐼^2^ = 30%, τ^2^ = 0.03, Q_add_ = 81.59, *df* = 57, p-value = 0.018). The Q test did not indicate a statistically significant difference between the NMA and CNMA,^212^ suggesting that there was no statistical evidence that the additivity assumption is violated (Q_diff_ = 26.98, *df* = 23; p-value = 0.257). Since the additive CNMA appeared adequate, we did not proceed to applying an interaction CNMA model.

In the additive CNMA model, we explored the individual component effects and noted large and imprecise effects in the components: *cbt* LOR = -1.00 (95% CI -2.46 to 0.46), *cp* LOR = -1.78 (95% CI -3.94 to 0.39), and *de-ft* LOR = 2.12 (95% CI -0.18 to 4.42) vs *UC*. **Appendix 8** presents the mean intervention effects for the falls-related fractures network estimated in the NMA and additive CNMA models. These results were mainly influenced by the available data, that is a single three-component intervention informing *cbt* (i.e., *cbt + cl-qi + pa-qi*) assessed in a single study in the network, a single three-component intervention informing *de-ft* (i.e., *cp + de-ft + pa-qi*) in a single study in the network, and two multi-component combinations (i.e., *bf + cp + ex + pa-qi* and *cp + de-ft + pa-qi*) informing *cp* in two studies in the network. Similarly, in NMA the largest effect was observed for *bf + cp + ex + pa-qi* vs *UC* with LOR = -2.14 (95% -4.32 to 0.04) and P-score = 91%. However, *bf + cp + ex + pa-qi* was compared in a single study with a small number of events in the entire network (i.e., *bf + cp + ex + pa-qi*: events/sample size = 1/153 vs *bf*: events/sample size = 7/152). The second largest effect in NMA was estimated in the comparison *fl* vs *UC* with LOR = -1.63 (95% -4.69 to 1.44) and P-score = 78%. Again, *fl* was compared in a single study with rare events in the entire network (i.e., *fl:* 0/210 vs *UC*: 2/208). Sparse data in the network may suggest differences in the estimation of intervention effects between NMA and CNMA. For example, the intervention effect for *op-tx* vs *UC* was estimated as LOR = 0.56 (95% -0.22 to 1.33) in NMA, and as LOR = -0.33 (95% -0.88 to 0.22) in CNMA. Considering that *op-tx* was compared in one multi-arm study both as a single and multi-component intervention (i.e., *op-tx*: 16/122 vs *op-tx + vi-d*: 8/122 vs *UC:* 10/122) and in another study in a different multi-component intervention (i.e., *ca + op-tx + vi-d*: 8/231 vs *ca + vi-d*: 27/230), this led to differences in this component estimation across NMA and CNMA models.

#### Quality of life using knowledge translation interventions

We demonstrate CNMA approaches by modelling a network of sustained KT interventions supporting chronic disease management that were assessed in RCTs of older adults (> 65 years old) with chronic disease(s), their caregivers, health and/or policy-decision makers.^213^ Study participants received a KT intervention/*UC* supporting chronic disease management for a minimum of 12 months. The outcome of interest was health-related QoL (continuous outcome). Data are presented in **Appendix 1**.

These data are reported in a published systematic review with meta-analysis^213^ that included 157 RCTs comprising 329 KT interventions, involving patients (43.2%), healthcare providers (20.7%) or both (10.9%). The Cochrane Effective Practice and Organisation of Care (EPOC) (**Appendix** **Table 2**) and the Behaviour Change Technique (*BCT*) were used to classify characteristics of the KT interventions identified by the systematic review. Examples of categories include patient/healthcare provider education, reminders, and financial incentives in EPOC and problem solving, action planning, and social support in *BCT*. There were 104 combinations of EPOC-derived components, and 190 combinations of *BCT*-derived components identified in the included studies. Despite the large number of studies, the original review could not explain the observed heterogeneity through a pairwise meta-analysis.

For the QoL outcome, 50 RCTs in the original publication reported data on final scores and change from baseline, using seven different measurement scales. These data were synthesized in pairwise meta-analyses for each measurement scale using the mean difference, with each meta-analysis including a range of nine to 16 KT interventions in a maximum of 15 RCTs. Due to the limited number of studies per measurement scale, we decided to restrict to the pairwise meta-analysis, since there was an insufficient number of data points to estimate the multiple parameters of the NMA model. However, in this study, we consider standardized scales and the standardized mean difference (SMD) effect measure. When using SMD, variations in the standard deviation (SD) indicate differences in the measurement reliability rather than variations in the measurement scale.^214^ Therefore, in this study, we restricted the analysis to RCTs assessing QoL using final scores only. In the present study, we also restrict components to the EPOC taxonomy to demonstrate the CNMA models. The control group was *UC*, which reflects not receiving a KT intervention for a patient or healthcare provider.

The QoL evidence included 35 RCTs (11,789 participants; 33 two-arm and two three-arm RCTs) with continuous data measured on multiple scales, comparing 29 different combinations of components (comprised of 15 individual components; **Appendix** **Table 2**) in a connected network (**Appendix** **Figure 2**). Thirty-three RCTs compared 24 different combinations of components with the inactive intervention *UC* (11,604 participants; 6,025 participants in KT interventions and 5,579 in *UC*). Active combinations of components contained one to seven components, with promotion of self-management (*s-M*) being the most frequent component included in the different combinations of components (30 in 127 total active components were assessed across all studies, 24%) (**Appendix 2**). Overall, *s-M* appeared in 20 out of 28 active combinations of components (71%) and was evaluated in 29 (83%) studies. Disentangling the effect of each KT component and examining its interactions is core to understanding what works well in KT interventions.

In Appendices 4-6 we also present ‘leave one component out’ scatter plots to explore component inclusion/exclusion on intervention efficacy, violin plots to explore the distribution of effects of individual components and component heat plots to explore the efficacy of component pairs in different combinations of components.

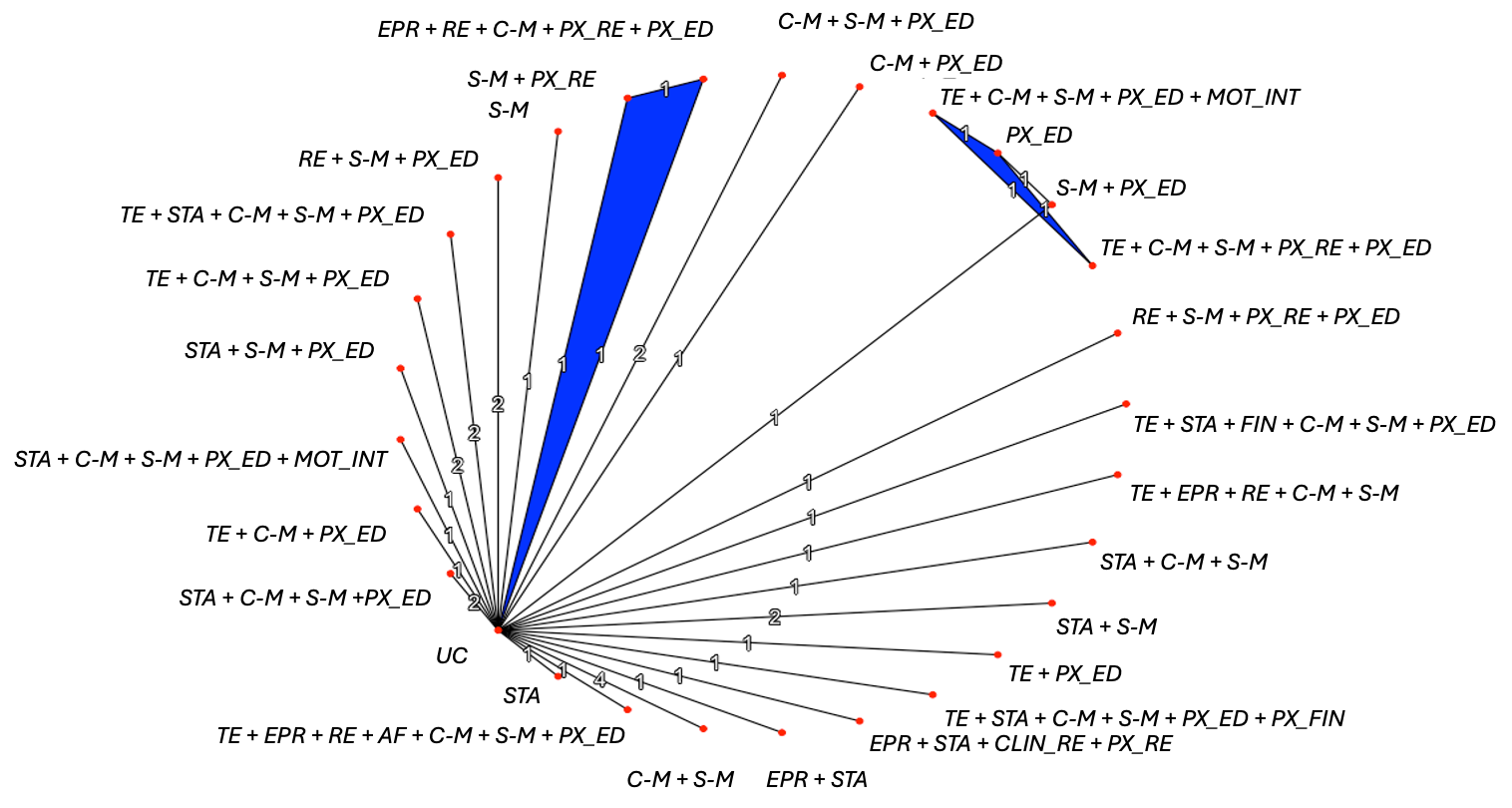

**Appendix Figure 2. Network diagram depicting network connectedness of complex interventions for quality of life. Nodes represent complex interventions, as combinations of individual components. Thickness of solid lines is proportional to the number of studies included in the group comparison, and node size is proportional to the number of patients included in the underlying group.**

**Appendix Table 2. Component abbreviations in quality of life** **with knowledge translation interventions**

| **Component in QoL with KT Interventions^1^** | **Abbreviation** |
| --- | --- |
| Usual Care | *UC* |
| Case management | *C-M* |
| Electronic patient registry | *EPR* |
| Patient education | *PX_ED* |
| Facilitated relay of info to clinicians | *RE* |
| Promotion of self-management | *S-M* |
| Staff education | *STA* |
| Team changes | *TE* |
| Patient reminders | *PX_RE* |
| Clinician reminders | *CLIN_RE* |
| Financial incentives | *FIN* |
| Motivational interview | *MOT_INT* |
| Financial incentives | *PX_FIN* |
| Audit & feedback | *AF* |

^1^ Cochrane Effective Practice and Organisation of Care (EPOC) distinct components in knowledge translation (KT) interventions for quality of life (QoL) outcome.

We were unable to assess for inconsistency in the QoL network, since the loops were informed only by multi-arm studies. Networks of complex interventions are often sparse (i.e., few studies and many interventions), which increases the challenge of evaluating transitivity and consistency assumptions. Empirical evidence has shown that inconsistency tests in sparse networks are, on average, associated with larger p-values compared to dense networks (i.e., many studies and few interventions), which is probably due to the decreased power in detecting inconsistency in sparse networks.^215^ Validity of transitivity can be assessed conceptually comparing the mean/mode of potential effect modifiers across intervention comparisons (e.g., using box plots and mean/mode of available evidence). In our case example, our team judged that there was no serious violation of transitivity and the RCTs contributing to the network geometry were similar enough to be analysed jointly in an NMA.

Evaluating the individual components in the additive CNMA model, we found that the additive CNMA led to more conservative (i.e., intervention effects closer to SMD = 0) and more precise results compared with NMA effects. **Appendix 9** presents the mean intervention effects estimated by each of the NMA and CNMA models for the 29 different combinations of the 15 components in the 35 RCTs in the QoL network. For example, the combination of components *re + s-M + px_re + px_ed* that ranked best in NMA according to the P-score ranking statistic (**Appendix** **Figure 3**) with SMD = 1.84 (95% CI 1.16 to 2.53) and P-score = 99%, had a more conservative and precise effect in CNMA: SMD = 0.42 (95% CI 0.06 to 0.78) and P-score = 91%. This is because available data contributed to estimating more parameters in NMA compared with CNMA (28 parameters for intervention effects vs 15 parameters for component effects). These differences led to variation in the ranking of combinations of components according to the P-score (**Appendix** **Figure 3**). Also, heterogeneity in the CNMA increased compared with the standard NMA model (NMA: 𝐼^2^ = 19%, τ^2^ = 0.00, Q_NMA_ = 11.11, *df* = 9, p-value = 0.268; Additive CNMA: 𝐼^2^ = 69%, τ^2^ = 0.03, Q_add_ = 77.82, *df* = 24, p-value < 0.001). The Q test for the difference between the NMA and CNMA (corresponding to the likelihood ratio test)^212^ was statistically significant, suggesting that the additivity assumption could not be justified for all combinations of components (Q_diff_ = 66.71, *df* = 15; p-value < 0.001). This suggested that combined components resulted in a larger or smaller effect than the sum of their individual effects. Since the additive CNMA could not be justified, we generated interaction CNMA models.

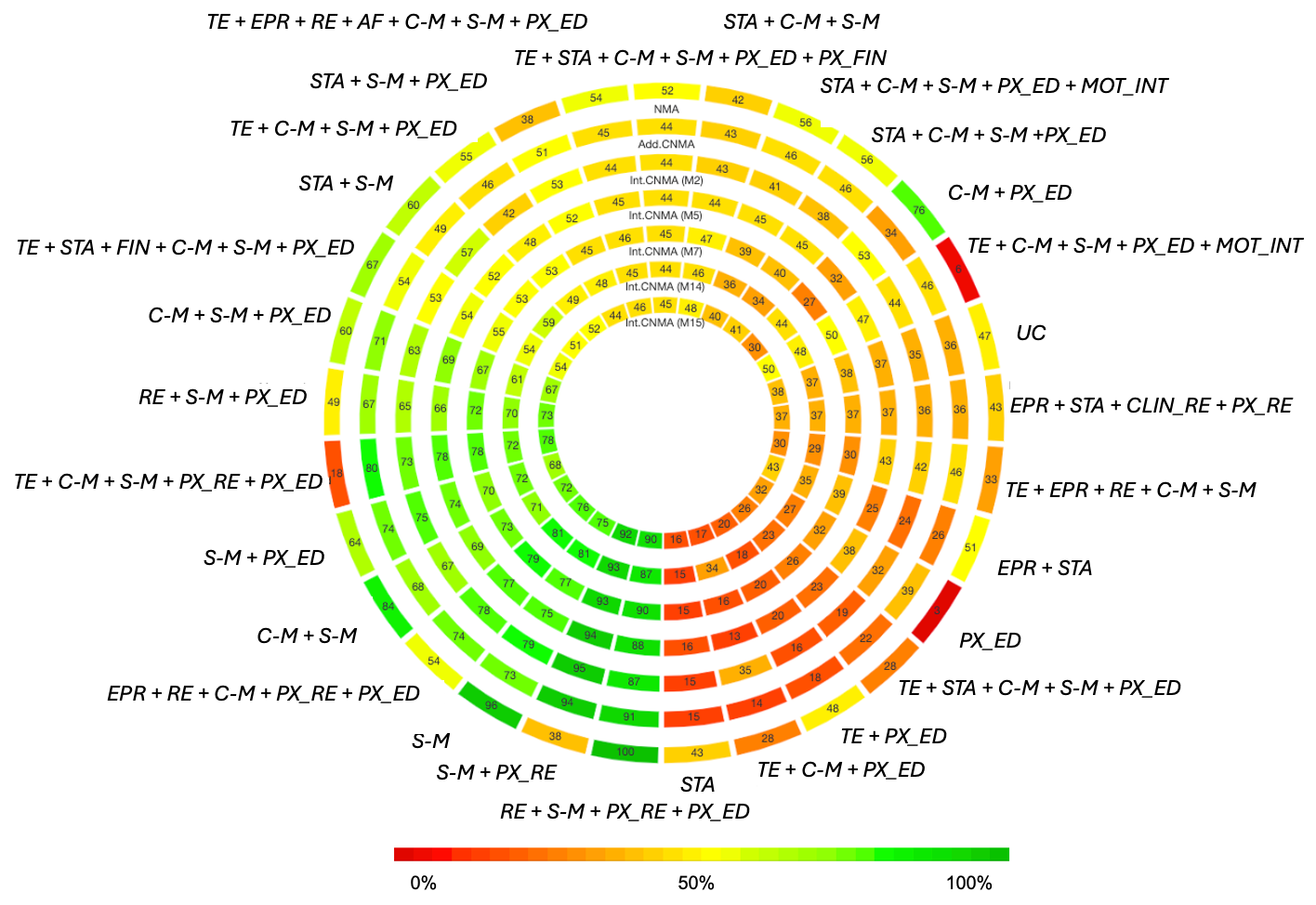

**Appendix Figure 3. Rank-heat plot for the quality of life outcome with KT interventions across different combinations of components and based on NMA and CNMA models. Numbers within each sector correspond to the P-score values as calculated within NMA. Each model corresponds to a separate ring starting with NMA, additive CNMA, and then interaction CNMA models (Model 2, Model 5, Model 7, Model 14, Model 15). Sectors are coloured according to P-score values as per the transformation of three colours red (0%; worst performing combination of components), yellow (50%), and green (100%; best performing combination of components).**

Unlike the previous example, the additivity assumption is not justifiable in the QoL network. Starting from the additive CNMA model, we used the forward selection procedure to add clinically relevant and identifiable interaction terms. The clinicians on the team suggested that all estimable interaction terms were clinically relevant. We relaxed the additivity assumption by adding systematically each estimable two-way and three-way interaction, as identified in the clinically relevant and available data (i.e., each component pair found in the available combinations with more than two components could be considered a potential two-way interaction and each set of three components found in the available combinations of more than three components could be considered a potential three-way interaction). We therefore have seven estimable two-way and five three-way interactions, all of which were considered clinically relevant (**Appendix** **Table 3**). Hence, we had data from seven combinations of two components, corresponding to seven models, each defined as the additive model plus one two-way interaction, and five combinations of three components, corresponding to an additional five models, each defined as the additive model plus one three-way interaction and the identifiable relevant lower order (two-way) interactions. Using the Q test (i.e., the difference in Cochran’s Q statistics), we compared each model to the additive CNMA, NMA, and the nested interaction model with the lowest Q to define the best fitting model. We subsequently combined the best fitting models to generate more complex multi-interaction models.

**Appendix Table 3. Identifiable and clinically relevant two-way and three-way interactions in the quality of life network.** **Models 1-7 arise due to including one 2nd order interaction, while models 8 to 12 arise from including one 3rd order interaction (with identifiable relevant 2nd order ones)**

| **Two-way interactions (*Corresponding Model*)** | **Three-way interactions**  **(*Corresponding Model*)** |
| --- | --- |
| 1. *c-M*px_ed*   *(Model 1)*   2. *c-M*s-M*   *(Model 2)*   3. *EPR*sta*   *(Model 3)*   4. *s-M*px_ed*   *(Model 4)*   5. *s-M*px_re*   *(Model 5)*   6. *sta*s-M*   *(Model 6)*   7. *te*px_ed*   *(Model 7)* | 1. *c-M*s-M*px_ed*   *(Model 8: c-M*s-M*px_ed + c-M*px_ed + c-M*s-M + s-M*px_ed)*   2. *re*s-M*px_ed*   *(Model 9: re*s-M*px_ed + s-M*px_ed)*   3. *sta*c-M*s-M*   *(Model 10: sta*c-M*s-M + sta*s-M + c-M*s-M)*   4. *sta*s-M*px_ed*   *(Model 11: sta*s-M*px_ed + sta*s-M + s-M*px_ed)*   5. *te*c-M*px_ed*   *(Model 12: te*c-M*px_ed + c-M*px_ed + te*px_ed)* |

Of the seven single-interaction models, only Models 2, 5, and 7 reduced the Q statistic significantly from that of the additive model, suggesting better fit to the data (**Appendix** **Table 3**). Three-way interaction CNMA models (Models 8 to 12) were considered by combining three-way interaction terms with their relevant identifiable lower-order two-way interaction terms. We recommend including relevant lower-order interactions, where identifiable, alongside higher-order interaction terms in the model. This approach can determine if the estimated effects are driven by only some components, rather than all, when possible. Additional two-interaction CNMA models (Models 13 to 15) were generated by combining the interactions from Models 2, 5, and 7. These interactions performed well individually, leading us to hypothesize that combining them might yield even better results. Comparing the interaction CNMA models with the NMA model, we obtained substantial differences in Q statistics (ranging between 66.46 and 61.26, with differences in *df* ranging from 12 to 14, and all p-values < 0.001) and increases in heterogeneity (from 𝐼^2^ = 19% for the standard NMA to a maximum of 𝐼^2^ = 72%; **Appendix** **Table 4**), suggesting that the NMA model should be preferred. However, if the aim of the review is to obtain inferences about individual components, given that the additivity assumption does not hold, Models 2, 5, and 7 are the best CNMA models to consider because they demonstrate significantly lower Q statistics compared to the additive CNMA, and are more parsimonious than Models 14 and 15, models with which they share similar Q statistics. Addition of further interaction terms may reduce the Q statistic further; however, our network includes a small number of studies per intervention comparison and there is a trade-off between the number of parameters to estimate and statistical power. The remaining models (Models 1, 3, 4, 6, and 8 to 13) did not show significantly lower Q statistics compared to the additive CNMA, and therefore, they are not considered further.

In this example, we explored adding second-order interactions individually and in combination, as well as incorporating third-order interactions one at a time. However, none of these approaches statistically significantly improved model fit compared to the standard NMA. While we could explore more complex models—including up to seven-way interactions due to the presence of a seven-component intervention in the data—the interpretation of such models becomes increasingly difficult, essentially approaching the standard NMA model. Also, even with clinically relevant interactions, these models might become overly data-driven. Since our aim was illustrative, we limited our exploration to CNMA models with up to three-way interactions.

Determining which interaction terms should be included in CNMA can be challenging. An entirely data-driven approach investigating all possible (and identifiable) interactions may result in spurious significant interactions and is time consuming for large networks with many components. In our QoL example, we used clinical knowledge along with a forward selection approach to identify interactions of interest that were identifiable in the data. We relaxed the additivity assumption by adding each estimable two-way and three-way interaction of interest individually, identifying improvements in model fit when the Q statistic of the interaction CNMA was significantly lower than that of the additive CNMA, using an adjusted p-value (e.g., p < 0.157).^216,217^ Alternative approaches are also possible, such as comparing heterogeneity or deviance information criteria.^218^

Entirely data-driven approaches to interaction CNMA model generation use a solely forward or backward selection approach. In backward selection, we would start with the standard NMA (full interaction) model and gradually remove interactions, one by one, until we identify the most parsimonious model with a Q statistic not significantly different from that of the NMA. This approach is only applicable to connected networks and has several limitations,^219^ one being that different models with the same Q statistic can lead to very different intervention effects, especially in sparse networks. Interaction CNMA model development solely based on a data-driven approach that compares model-fit statistics is discouraged.^212,219^ When the additivity assumption is violated, interaction terms in CNMA should ideally be specified *a priori*, based on clinical expertise and the literature to avoid identifying spuriously significantly better models.

**Appendix Table 4. Model selection for the quality of life outcome. Statistically significant differences have been highlighted in bold text. Q statistics along with their degrees of freedom (df) and p-value for identifiable two-way and three-way interactions in interaction CNMA models.**

| **Model** | **Q-value** | ***df*** | **p-value** | **τ^2^** | **I^2^** | **Comparison with standard NMA (Additive/Interaction CNMA - Standard NMA)** | | | **Comparison with additive CNMA (Additive - Interaction CNMA)** | | | **Comparison with previous nested model with lowest Q (Nested Single-Interaction - Multiple-Interaction CNMA)** | | |
| --- | --- | --- | --- | --- | --- | --- | --- | --- | --- | --- | --- | --- | --- | --- |
|  |  |  |  |  |  | **Q_diff_** | ***df*** | **p-value** | **Q_diff_** | ***df*** | **p-value** | **Q_diff_** | ***df*** | **p-value** |
| Standard NMA | 11.11 | 9 | 0.268 | 0.00 | 19% |  |  |  |  |  |  |  |  |  |
| Additive CNMA | 77.82 | 24 | **<0.001** | 0.04 | 69% | 66.71 | 15 | **<0.001 ^a^** |  |  |  |  |  |  |
| **Single-interaction (two-way interactions) CNMA models** | | | | | | | | | | | | | | |
| 1) *c-M * px_ed* | 76.73 | 23 | **<0.001** | 0.04 | 70% | 65.62 | 14 | **<0.001 ^b^** | 1.09 | 1 | 0.297 ^c^ |  |  |  |
| 2) *c-M * s-M* | 75.45 | 23 | **<0.001** | 0.04 | 70% | 64.34 | 14 | **<0.001 ^b^** | 2.37 | 1 | **0.124 ^c^** |  |  |  |
| 3) *EPR * sta* | 77.57 | 23 | **<0.001** | 0.04 | 70% | 66.46 | 14 | **<0.001 ^b^** | 0.25 | 1 | 0.614 ^c^ |  |  |  |
| 4) *s-M * px_ed* | 75.96 | 23 | **<0.001** | 0.04 | 70% | 64.85 | 14 | **<0.001 ^b^** | 1.86 | 1 | 0.173 ^c^ |  |  |  |
| 5) *s-M * px_re* | 75.39 | 23 | **<0.001** | 0.04 | 70% | 64.28 | 14 | **<0.001 ^b^** | 2.43 | 1 | **0.119 ^c^** |  |  |  |
| 6) *sta * s-M* | 77.28 | 23 | **<0.001** | 0.04 | 70% | 66.17 | 14 | **<0.001 ^b^** | 0.54 | 1 | 0.464 ^c^ |  |  |  |
| 7) *te * px_ed* | 73.90 | 23 | **<0.001** | 0.04 | 69% | 62.79 | 14 | **<0.001 ^b^** | 3.92 | 1 | **0.048 ^c^** |  |  |  |
| **Multi-interaction (three-way interaction and relevant lower-order interactions) CNMA models** | | | | | | | | | | | | | | |
| 8) *c-M * s-M * px_ed + c-M * px_ed*  *+ c-M * s-M*  *+ s-M * px_ed* | 74.21 | 21 | **<0.001** | 0.04 | 72% | 63.10 | 12 | **<0.001 ^d^** | 3.61 | 3 | 0.307 ^f^ |  |  |  |
| 9) *re * s-M * px_ed*  *+ s-M * px_ed* | 75.24 | 22 | **<0.001** | 0.04 | 71% | 64.13 | 13 | **<0.001 ^e^** | 2.58 | 2 | 0.275 ^g^ |  |  |  |
| 10) *sta * c-M * s-M*  *+ sta * s-M*  *+ c-M * s-M* | 74.29 | 21 | **<0.001** | 0.04 | 72% | 63.18 | 12 | **<0.001 ^d^** | 3.53 | 3 | 0.317 ^f^ |  |  |  |
| 11)*sta * s-M * px_ed*  *+ sta * s-M*  *+ s-M * px_ed* | 75.03 | 21 | **<0.001** | 0.04 | 72% | 63.92 | 12 | **<0.001 ^d^** | 6.40 | 4 | 0.171 ^h^ |  |  |  |
| 12) *te * c-M * px_ed*  *+ c-M * px_ed*  *+ te * px_ed* | 76.65 | 22 | **<0.001** | 0.04 | 71% | 65.54 | 13 | **<0.001 ^e^** | 1.17 | 2 | 0.558 ^g^ |  |  |  |
| **Two two-way interaction CNMA models** | | | | | | | | | | | | | | |
| 13) *c-M * s-M*  *+ s-M * px_re* | 74.34 | 22 | **<0.001** | 0.04 | 70% | 63.23 | 13 | **<0.001 ^e^** | 3.48 | 2 | 0.175 ^g^ | 1.05 ^i^ | 1 | 0.305 ^c^ |
| 14) *c-M * s-M*  *+ te * px_ed* | 72.43 | 22 | **<0.001** | 0.04 | 70% | 61.32 | 13 | **<0.001 ^e^** | 5.39 | 2 | **0.067 ^g^** | 1.48 ^j^ | 1 | 0.224 ^c^ |
| 15) *s-M * px_re*  *+ te * px_ed* | 73.49 | 22 | **<0.001** | 0.04 | 70% | 62.38 | 13 | **<0.001 ^e^** | 4.33 | 2 | **0.115 ^g^** | 0.42 ^j^ | 1 | 0.519 ^c^ |

a Compared to a p-value of 0.012 (obtained in R as p-value=1-pchisq(2*15,15))

b Compared to a p-value of 0.014 (obtained in R as p-value=1-pchisq(2*14,14))

c Compared to a p-value of 0.157 (obtained in R as p-value=1-pchisq(2*1,1))

d Compared to a p-value of 0.020 (obtained in R as p-value=1-pchisq(2*12,12))

e Compared to a p-value of 0.017 (obtained in R as p-value=1-pchisq(2*13,13))

f Compared to a p-value of 0.112 (obtained in R as p-value=1-pchisq(2*3,3))

g Compared to a p-value of 0.135 (obtained in R as p-value=1-pchisq(2*2,2))

h Compared to a p-value of 0.092 (obtained in R as p-value=1-pchisq(2*4,4))

i Compared to Model 5

j Compared to Model 7

### Appendix 4. Leave one component out scatter plots for intervention comparisons that differ by one component using NMA results in quality of life.

Each scatterplot corresponds to a separate component included in the network.

Appendix 4 depicts a ‘leave one component out’ scatter plot for each combination of components that differ by a specific component. Overall, the inclusion of the assessed components leads to higher SMDs, whereas points close to the diagonal indicate that the additivity assumption is possible in these effects. That is, under the additivity assumption, component effects (e.g., *C-M, px_ed*) are expected to be consistent across the two combinations of components groups (e.g., *te + c-M + px_ed vs c-M + px_ed*) resulting in their cancellation within the overall intervention effect. Hence, the intervention effect can be attributed to the difference between treatments *te* vs. *UC.*

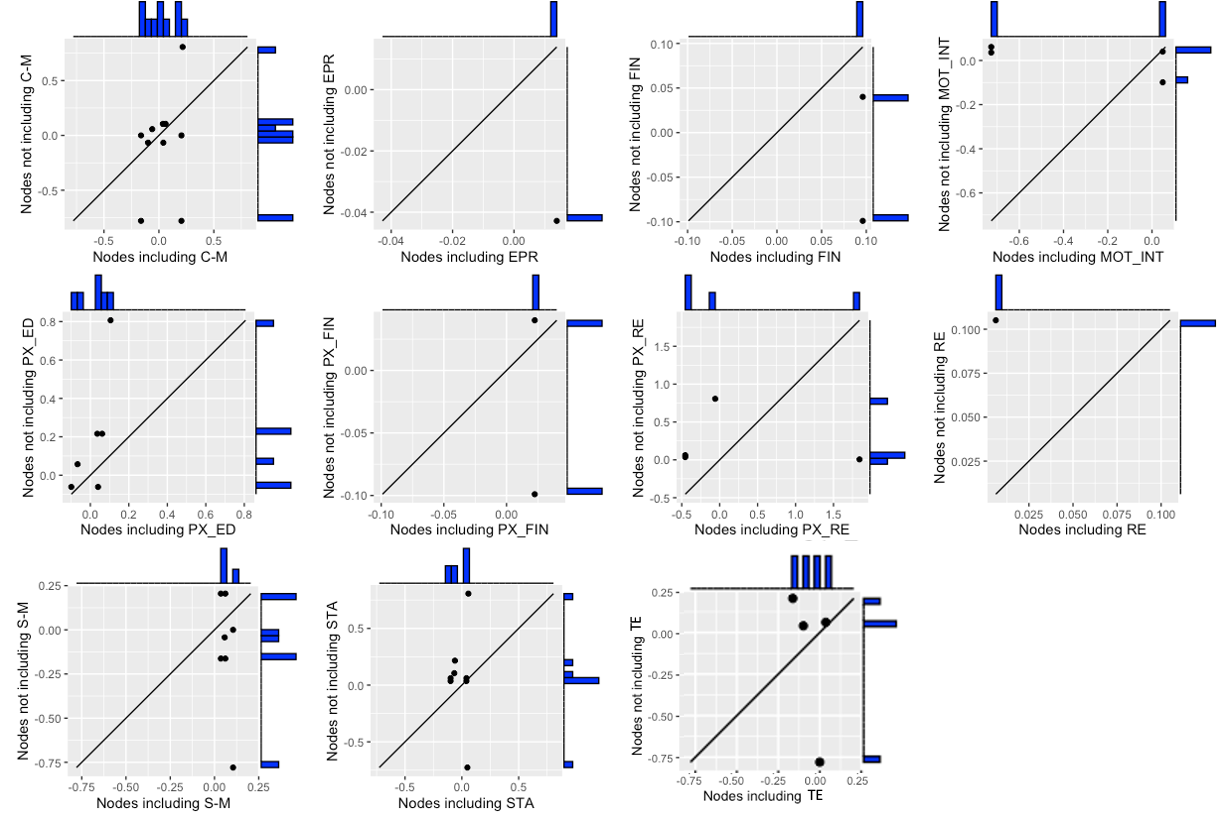

### Appendix 5. Violin plots for the efficacy of components using NMA results in quality of life.

Plot requires at least two data points per component to present a box plot.

Appendix 5 shows the box plots of the individual component effects as estimated in the combinations that they were part of in the NMA model. The component that was part of the least efficacious combination of components was *mot_int* with a median SMD = -0.34.

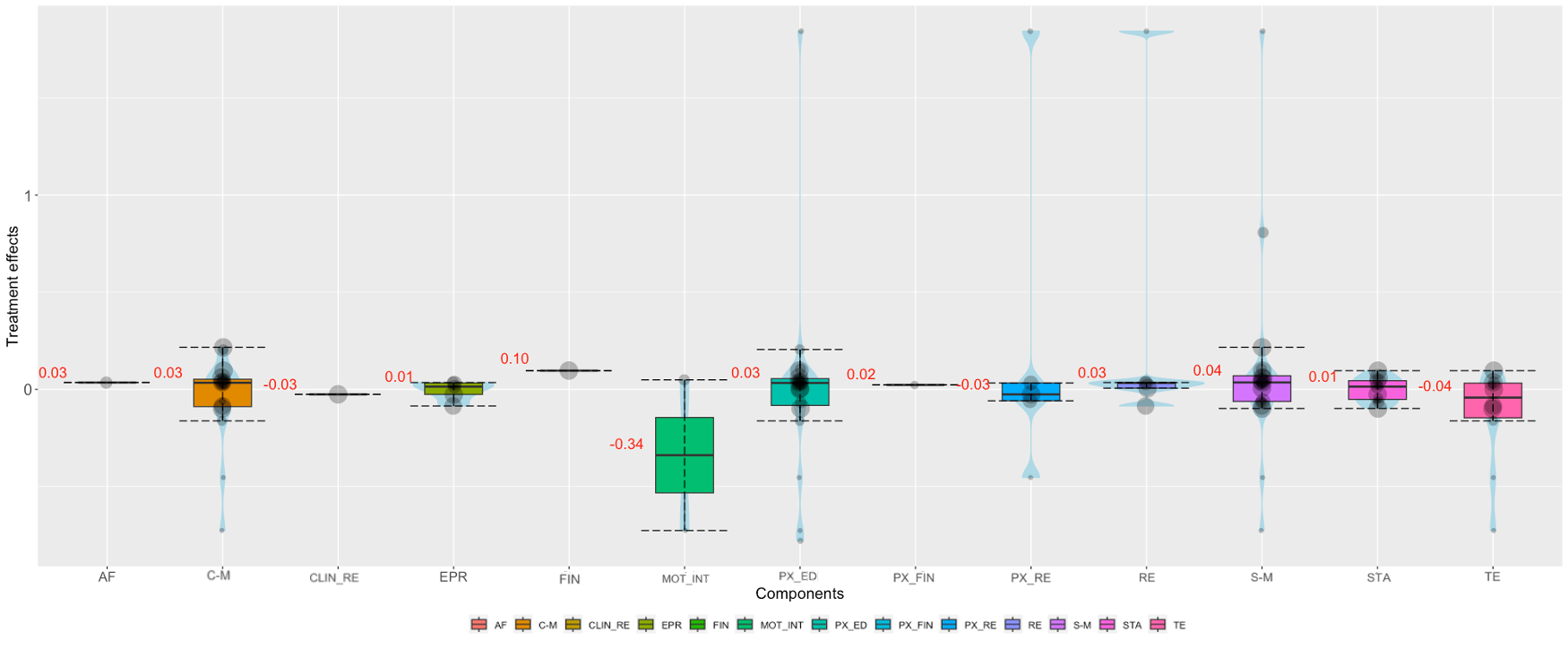

### Appendix 6. Component heat plots for the efficacy of pairs of combinations of components using NMA results in quality of life.

Each cell presents the median NMA effect of all KT interventions that include the paired combination of components. Grey boxes are proportional to the underlying precision of the intervention effect estimate. Empty cells correspond to unidentifiable combinations of components in the data.

Appendix 6 presents the effects of pairwise component combinations as a median SMD of all the combinations that they were part of the NMA model. For example, the combination of components *px_re + s-M* was included in three interventions (*re + s-M + px_re + px_ed, te + c-M + s-M + px_re + px_ed, s-M + px_re*) and the median estimated SMD across these interventions vs *UC* was -0.06. As median SMD increases in the plot, the colour intensity also increases, indicating that the underlying pairwise component combination performs better/worse than *UC* (depending on the direction).

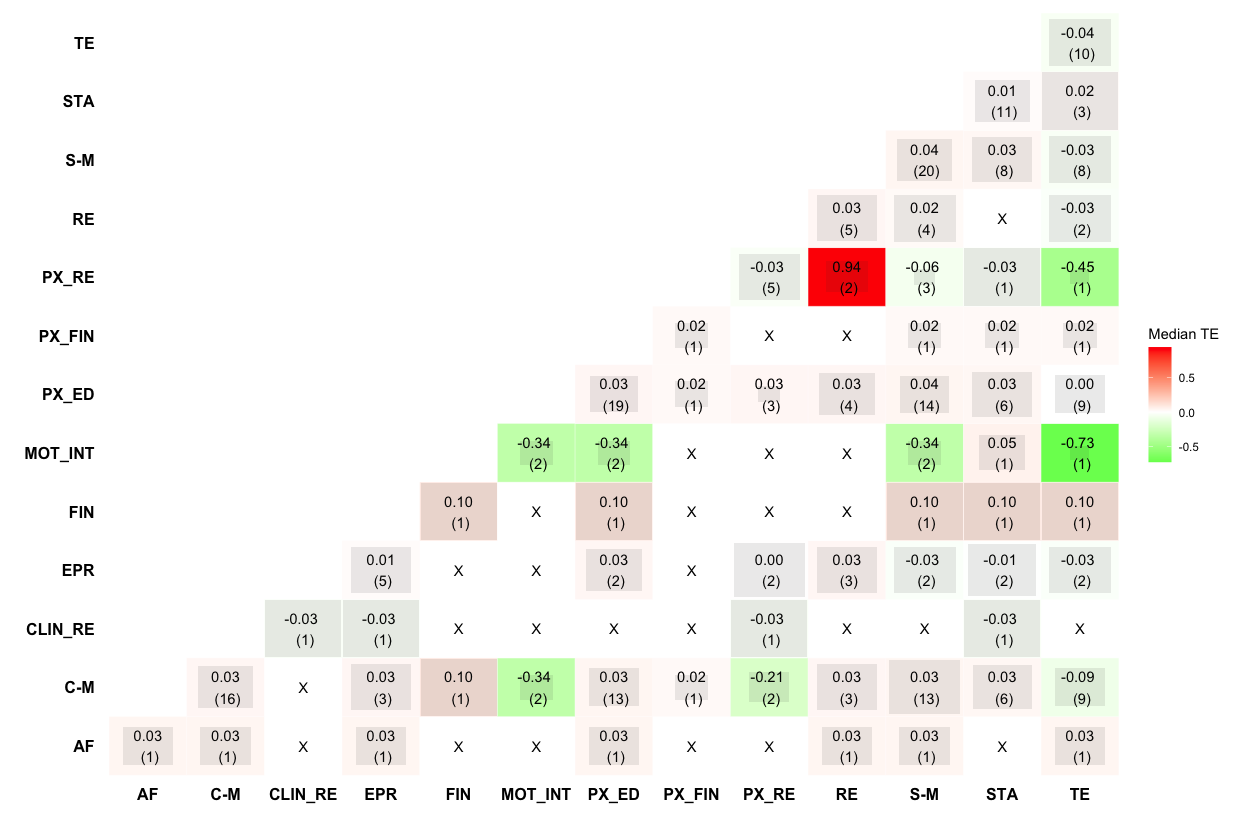

### Appendix 7. Prevention of post-operative complications NMA and CNMA results.

Treatment and component effects (odds ratios) along with their 95% confidence intervals for standard network meta-analysis (NMA), additive component NMA (CNMA), and interaction CNMA models.

| **Combination of components vs UC** | **NMA** | **Additive CNMA** | **Interaction CNMA  (Model 1)** |
| --- | --- | --- | --- |
| *cog* | 0.92 (0.43, 1.94) | 0.91 (0.43, 1.92) | 0.91 (0.43, 1.91) |
| *EXE* | 0.50 (0.39, 0.64) | 0.53 (0.42, 0.66) | 0.51 (0.40, 0.64) |
| *EXE + nut* | 0.52 (0.26, 1.05) | 0.34 (0.26, 0.46) | 0.48 (0.29, 0.80) |
| *EXE + nut + pSY* | 0.64 (0.45, 0.92) | 0.61 (0.44, 0.84) | 0.66 (0.47, 0.92) |
| *EXE + PSY* | 0.77 (0.37, 1.59) | 0.92 (0.65, 1.32) | 0.70 (0.41, 1.18) |
| *NUT* | 0.62 (0.50, 0.77) | 0.66 (0.54, 0.81) | 0.62 (0.50, 0.77) |
| *PSY* | 1.36 (0.24, 7.76) | 1.75 (1.67, 2.61) | 1.38 (0.82, 2.30) |
| **Component vs UC** | **NMA** | **Additive CNMA** | **Interaction CNMA  (Model 1)** |
| *COG* | -- | 0.91 (0.43, 1.91) | 0.91 (0.43, 1.91) |
| *EXE* | -- | 0.53 (0.42, 0.66) | 0.51 (0.40, 0.64) |
| *NUT* | -- | 0.66 (0.54, 0.81) | 0.62 (0.50, 0.77) |
| *PSY* | -- | 1.75 (1.17, 2.61) | 1.38 (0.82, 2.30) |
| *EXE * NUT* | -- | -- | 1.52 (0.86, 2.71) |
| τ^2^ | 0.15 | 0.14 | 0.14 |
| I^2^ | 31% | 30% | 30% |
| Q, df, p-value | 144.20, 100, 0.003 | 147.72, 103, 0.003 | 144.69, 102, 0.004 |
| Difference in Q statistics (CNMA and NMA model) | -- | 3.52, 3, 0.318 | 0.49, 2, 0.784 |

P-scores for treatments and components within standard network meta-analysis (NMA), additive component NMA (CNMA), and interaction CNMA models.

| **Treatment** | **NMA** | **Additive CNMA** | **Interaction CNMA  (Model 1)** |
| --- | --- | --- | --- |
| *cog* | 0.31 | 0.36 | 0.32 |
| *EXE* | 0.85 | 0.80 | 0.86 |
| *EXE + nut* | 0.75 | 1.00 | 0.85 |
| *EXE + nut + pSY* | 0.59 | 0.68 | 0.57 |
| *EXE + PSY* | 0.44 | 0.32 | 0.53 |
| *NUT* | 0.62 | 0.59 | 0.63 |
| *PSY* | 0.24 | 0.01 | 0.05 |
| *UC* | 0.19 | 0.25 | 0.20 |

### Appendix 8. Prevention of falls-related fractures NMA and CNMA results.

Treatment and component effects (log odds ratios) along with their 95% confidence intervals for network meta-analysis (NMA) and additive component NMA (CNMA) models.

| **Combinations of components vs UC** | **NMA** | **Additive CNMA** | **Component vs UC** | **Additive CNMA** |
| --- | --- | --- | --- | --- |
| *bf* | -0.14 (-0.57, 0.28) | -0.14 (-0.45, 0.17) | *bf* | -0.14 (-0.45, 0.17) |
| *bf + cl-qi* | 0.10 (-1.20, 1.39) | 0.03 (-0.42, 0.47) | *ca* | -0.18 (-0.37, 0.00) |
| *bf + cl-qi + pa-qi* | -0.65 (-3.41, 2.11) | -0.03 (-0.50, 0.44) | *cbt* | -1.00 (-2.46, 0.46) |
| *bf + cp + ex + pa-qi* | -2.14 (-4.32, 0.04) | -2.13 (-4.29, 0.02) | *cl-qi* | 0.17 (-0.19, 0.52) |
| *bf + su-pm* | -0.41 (-2.03, 1.22) | -0.4 (-1.99, 1.19) | *cp* | -1.78 (-3.94, 0.39) |
| *ca* | -0.12 (-0.43, 0.18) | -0.18 (-0.37, 0.00) | *de-al* | -0.31 (-1.61, 0.99) |
| *ca + cl-qi + di + pa-qi + vi-d* | -1.11 (-4.34, 2.12) | -0.33 (-1.19, 0.52) | *de-hp* | -0.16 (-0.48, 0.17) |
| *ca + cl-qi + mf + vi-d* | -0.55 (-1.61, 0.51) | -0.21 (-0.51, 0.08) | *de-ft* | 2.12 (-0.18, 4.42) |
| *ca + ea + pa-qi + vi-d* | -0.21 (-0.65, 0.22) | -0.28 (-0.59, 0.03) | *di* | -0.26 (-1.01, 0.49) |
| *ca + op-tx + vi-d* | -1.54 (-2.46, -0.62) | -0.51 (-1.07, 0.04) | *ea* | -0.04 (-0.32, 0.24) |
| *ca + pa-qi + vi-d* | -0.22 (-0.64, 0.20) | -0.24 (-0.50, 0.02) | *ex* | -0.16 (-0.45, 0.13) |
| *ca + vi-d* | -0.23 (-0.46, 0.00) | -0.19 (-0.35, -0.02) | *fl* | -1.63 (-4.69, 1.43) |
| *cbt + cl-qi + pa-qi* | -0.89 (-2.31, 0.52) | -0.89 (-2.30, 0.51) | *mf* | -0.19 (-0.56, 0.17) |
| *cl-qi* | 0.09 (-1.07, 1.24) | 0.17 (-0.19, 0.52) | *op-tx* | -0.33 (-0.86, 0.20) |
| *cl-qi + de-al + ex + mf + pa-qi* | -0.45 (-2.28, 1.37) | -0.56 (-1.86, 0.74) | *pa-qi* | -0.06 (-0.30, 0.18) |
| *cl-qi + de-hp* | 0.07 (-0.40, 0.54) | 0.01 (-0.34, 0.36) | *so* | 0.13 (-0.43, 0.69) |
| *cl-qi + ex + mf* | 0.15 (-0.67, 0.96) | -0.19 (-0.52, 0.14) | *su-ey* | -0.55 (-1.48, 0.39) |
| *cl-qi + ex + mf + pa-qi* | -0.01 (-2.49, 2.47) | -0.25 (-0.60, 0.11) | *su-pm* | -0.26 (-1.82, 1.30) |
| *cl-qi + ex + pa-qi* | 1.08 (-2.18, 4.35) | -0.05 (-0.59, 0.49) | *va* | 0.62 (-0.11, 1.36) |
| *cl-qi + mf* | 0.14 (-0.35, 0.63) | -0.03 (-0.26, 0.20) | *vi-d* | 0.00 (-0.15, 0.14) |
| *cl-qi + mf + pa-qi* | -0.16 (-0.45, 0.14) | -0.09 (-0.33, 0.15) | -- | -- |
| *cl-qi + mf + pa-qi + so* | -0.05 (-0.85, 0.75) | 0.04 (-0.54, 0.62) | -- | -- |
| *cp + de-ft + pa-qi* | 0.32 (-0.59, 1.23) | 0.29 (-0.52, 1.09) | -- | -- |
| *de-al* | -0.41 (-2.24, 1.42) | -0.31 (-1.61, 0.99) | -- | -- |
| *de-hp + ea + ex + pa-qi* | -0.15 (-0.77, 0.48) | -0.41 (-0.82, -0.01) | -- | -- |
| *de-hp + pa-qi* | -0.37 (-1.01, 0.27) | -0.21 (-0.61, 0.18) | -- | -- |
| *di* | -0.28 (-1.26, 0.69) | -0.26 (-1.01, 0.49) | -- | -- |
| *di + ex* | -0.66 (-1.72, 0.39) | -0.42 (-1.24, 0.41) | -- | -- |
| *ea + ex + mf* | -0.69 (-1.71, 0.32) | -0.4 (-0.83, 0.04) | -- | -- |
| *ea + pa-qi* | -0.19 (-0.61, 0.23) | -0.1 (-0.37, 0.17) | -- | -- |
| *ea + va* | 0.58 (-0.12, 1.29) | 0.58 (-0.10, 1.27) | -- | -- |
| *ex* | -0.42 (-0.89, 0.05) | -0.16 (-0.45, 0.13) | -- | -- |
| *ex + mf* | -1.27 (-3.86, 1.31) | -0.36 (-0.74, 0.03) | -- | -- |
| *ex + pa-qi* | -0.25 (-1.13, 0.63) | -0.22 (-0.61, 0.17) | -- | -- |
| *fl* | -1.63 (-4.69, 1.44) | -1.63 (-4.69, 1.43) | -- | -- |
| *mf* | -0.08 (-0.68, 0.53) | -0.19 (-0.56, 0.17) | -- | -- |
| *op-tx* | 0.56 (-0.22, 1.33) | -0.33 (-0.86, 0.20) |  |  |
| *op-tx + vi-d* | -0.21 (-1.12, 0.70) | -0.33 (-0.88, 0.22) |  |  |
| *pa-qi* | -0.03 (-0.47, 0.42) | -0.06 (-0.30, 0.18) |  |  |
| *pa-qi + vi-d* | -0.09 (-1.15, 0.97) | -0.06 (-0.33, 0.22) |  |  |
| *so* | 0.18 (-0.96, 1.33) | 0.13 (-0.43, 0.69) |  |  |
| *su-ey* | -0.54 (-1.49, 0.40) | -0.55 (-1.48, 0.39) |  |  |
| *vi-d* | 0.06 (-0.13, 0.26) | 0.00 (-0.15, 0.14) | -- | -- |
| **Quantifying heterogeneity / inconsistency:** | | | | |
| τ^2^ | 0.03 | 0.03 | | |
| I^2^ | 38% | 30% | | |
| Q, df, p-value | 54.62, 34, 0.014 | 81.59, 57, 0.018 | | |
| Difference in Q statistics (Additive and NMA model) | 18.75, 15, 0.225 | 26.98, 23, 0.257 | | |

P-scores for treatments and components within standard network meta-analysis (NMA) and additive component NMA (CNMA) models.

| **Treatment** | **NMA** | **Additive CNMA** |
| --- | --- | --- |
| *BF* | 0.4712 | 0.4573 |
| *BF+CL-QI* | 0.3613 | 0.286 |
| *BF+CL-QI+PA-QI* | 0.5995 | 0.3474 |
| *BF+CP+EX+PA-QI* | 0.9141 | 0.9436 |
| *BF+SU-PM* | 0.5635 | 0.5823 |
| *CA* | 0.4536 | 0.5147 |
| *CA+CL-QI+DI+PA-QI+VI-D* | 0.6793 | 0.6033 |
| *CA+CL-QI+MF+VI-D* | 0.6543 | 0.5517 |
| *CA+EA+PA-QI+VI-D* | 0.5198 | 0.6298 |
| *CA+OP-TX+VI-D* | 0.9128 | 0.7784 |
| *CA+PA-QI+VI-D* | 0.5269 | 0.595 |
| *CA+VI-D* | 0.5443 | 0.5176 |
| *CBT+CL-QI+PA-QI* | 0.7425 | 0.7926 |
| *CL-QI* | 0.3587 | 0.1484 |
| *CL-QI+DE-AL+EX+MF+PA-QI* | 0.5723 | 0.6828 |
| *CL-QI+DE-HP* | 0.3222 | 0.2942 |
| *CL-QI+EX+MF* | 0.3067 | 0.5153 |
| *CL-QI+EX+MF+PA-QI* | 0.4322 | 0.5929 |
| *CL-QI+EX+PA-QI* | 0.2255 | 0.3699 |
| *CL-QI+MF* | 0.2798 | 0.3117 |
| *CL-QI+MF+PA-QI* | 0.4835 | 0.3862 |
| *CL-QI+MF+PA-QI+SO* | 0.4114 | 0.2986 |
| *CP+DE-FT+PA-QI* | 0.2333 | 0.1778 |
| *DE-AL* | 0.5588 | 0.5456 |
| *DE-HP+EA+EX+PA-QI* | 0.4698 | 0.7371 |
| *DE-HP+PA-QI* | 0.6098 | 0.5387 |
| *DI* | 0.5347 | 0.5428 |
| *DI+EX* | 0.7019 | 0.6672 |
| *EA+EX+MF* | 0.7146 | 0.7072 |
| *EA+PA-QI* | 0.5033 | 0.3993 |
| *EA+VA* | 0.1228 | 0.0521 |
| *EX* | 0.6498 | 0.4757 |
| *EX+MF* | 0.7494 | 0.6842 |
| *EX+PA-QI* | 0.5231 | 0.5499 |
| *FL* | 0.7838 | 0.8058 |
| *MF* | 0.4247 | 0.5104 |
| *OP-TX* | 0.1355 | 0.6243 |
| *OP-TX+VI-D* | 0.5014 | 0.6227 |
| *PA-QI* | 0.3825 | 0.3432 |
| *PA-QI+VI-D* | 0.4376 | 0.3489 |
| *SO* | 0.3141 | 0.2245 |
| *SU-EY* | 0.6611 | 0.7159 |
| *UC* | 0.3499 | 0.259 |
| *VI-D* | 0.3031 | 0.268 |

### Appendix 9. Quality of life NMA and CNMA results.

Treatment and component effects (standardized mean differences) along with their 95% confidence intervals for network meta-analysis (NMA), additive component NMA (CNMA), and interaction CNMA models.

| **Combination of components vs UC** | **NMA** | **Additive CNMA** | **Interaction CNMA  (Model 2)** | **Interaction CNMA  (Model 5)** | **Interaction CNMA  (Model 7)** | **Interaction CNMA  (Model 14)** | **Interaction CNMA  (Model 15)** |
| --- | --- | --- | --- | --- | --- | --- | --- |
| *c-M + px_ed* | 0.21 (-0.15, 0.56) | -0.02 (-0.21, 0.18) | 0.09 (-0.22, 0.40) | -0.04 (-0.29, 0.21) | -0.07 (-0.29, 0.15) | 0.03 (-0.31, 0.37) | -0.06 (-0.31, 0.19) |
| *c-M + s-M* | 0.22 (0.09, 0.34) | 0.16 (0.00, 0.33) | 0.17 (0.00, 0.34) | 0.17 (0.00, 0.34) | 0.19 (0.02, 0.37) | 0.19 (0.01, 0.37) | 0.20 (0.01, 0.38) |
| *c-M + s-M + px_ed* | 0.06 (-0.13, 0.25) | 0.18 (-0.01, 0.37) | 0.15 (-0.06, 0.35) | 0.17 (-0.02, 0.37) | 0.15 (-0.04, 0.35) | 0.13 (-0.08, 0.34) | 0.16 (-0.04, 0.36) |
| *EPR + re + c-M + px_re + px_ed* | 0.03 (-0.22, 0.28) | 0.25 (-0.11, 0.61) | 0.28 (-0.09, 0.66) | 0.28 (-0.13, 0.70) | 0.29 (-0.08, 0.66) | 0.32 (-0.06, 0.70) | 0.28 (-0.14, 0.70) |
| *EPR + sta* | 0.01 (-0.26, 0.29) | -0.09 (-0.43, 0.26) | -0.10 (-0.45, 0.24) | -0.11 (-0.49, 0.26) | -0.01 (-0.38, 0.36) | -0.03 (-0.42, 0.35) | 0.01 (-0.45, 0.48) |
| *EPR + sta + clin_re + px_re* | -0.03 (-0.21, 0.16) | -0.03 (-0.43, 0.38) | -0.03 (-0.43, 0.38) | -0.03 (-0.43, 0.38) | -0.03 (-0.43, 0.38) | -0.03 (-0.43, 0.38) | -0.03 (-0.44, 0.39) |
| *px_ed* | -0.78 (-1.42, -0.14) | 0.01 (-0.17, 0.19) | -0.02 (-0.22, 0.17) | 0.00 (-0.19, 0.19) | -0.04 (-0.25, 0.17) | -0.07 (-0.28, 0.15) | -0.04 (-0.25, 0.17) |
| *re + s-M + px_ed* | 0.01 (-0.15, 0.17) | 0.17 (-0.09, 0.43) | 0.17 (-0.09, 0.43) | 0.16 (-0.10, 0.43) | 0.21 (-0.07, 0.48) | 0.2 (-0.07, 0.48) | 0.22 (-0.07, 0.51) |
| *re + s-M + px_re + px_ed* | 1.84 (1.16, 2.53) | 0.42 (0.06, 0.78) | 0.38 (0.01, 0.76) | 0.39 (-0.01, 0.80) | 0.40 (0.04, 0.76) | 0.38 (0.00, 0.75) | 0.43 (0.02, 0.84) |
| *s-M* | 0.81 (0.48, 1.14) | 0.19 (0.03, 0.36) | 0.25 (0.05, 0.45) | 0.21 (0.02, 0.41) | 0.22 (0.05, 0.40) | 0.27 (0.06, 0.48) | 0.22 (0.02, 0.42) |
| *s-M + px_ed* | 0.11 (-0.23, 0.44) | 0.21 (-0.05, 0.46) | 0.23 (-0.03, 0.49) | 0.22 (-0.04, 0.48) | 0.18 (-0.07, 0.44) | 0.2 (-0.06, 0.47) | 0.18 (-0.10, 0.45) |
| *s-M + px_re* | -0.06 (-0.31, 0.19) | 0.45 (0.12, 0.77) | 0.46 (0.14, 0.79) | 0.44 (0.12, 0.77) | 0.42 (0.09, 0.75) | 0.44 (0.11, 0.77) | 0.43 (0.09, 0.76) |
| *sta* | -0.04 (-0.41, 0.33) | -0.14 (-0.33, 0.06) | -0.14 (-0.33, 0.06) | -0.14 (-0.33, 0.06) | -0.15 (-0.35, 0.05) | -0.15 (-0.35, 0.05) | -0.15 (-0.35, 0.05) |
| *sta + c-M + s-M* | -0.06 (-0.57, 0.45) | 0.03 (-0.20, 0.26) | 0.03 (-0.20, 0.26) | 0.03 (-0.20, 0.26) | 0.04 (-0.18, 0.27) | 0.04 (-0.19, 0.28) | 0.05 (-0.19, 0.28) |
| *sta + c-M + s-M + px_ed* | 0.04 (-0.17, 0.25) | 0.04 (-0.14, 0.23) | 0.01 (-0.19, 0.21) | 0.03 (-0.16, 0.23) | 0.00 (-0.19, 0.20) | -0.02 (-0.23, 0.19) | 0.01 (-0.20, 0.21) |
| *sta + c-M + s-M + px_ed + mot_int* | 0.05 (-0.28, 0.38) | 0.05 (-0.33, 0.42) | 0.01 (-0.36, 0.39) | 0.03 (-0.35, 0.42) | -0.01 (-0.39, 0.38) | -0.03 (-0.42, 0.36) | 0.00 (-0.40, 0.39) |
| *sta + s-M* | 0.06 (-0.09, 0.20) | 0.06 (-0.12, 0.24) | 0.11 (-0.10, 0.33) | 0.07 (-0.13, 0.28) | 0.07 (-0.11, 0.25) | 0.12 (-0.10, 0.33) | 0.07 (-0.14, 0.27) |
| *sta + s-M + px_ed* | -0.07 (-0.38, 0.25) | 0.07 (-0.14, 0.28) | 0.09 (-0.12, 0.30) | 0.08 (-0.14, 0.29) | 0.03 (-0.19, 0.25) | 0.05 (-0.18, 0.28) | 0.03 (-0.22, 0.27) |
| *te + c-M + px_ed* | -0.16 (-0.55, 0.22) | -0.15 (-0.36, 0.06) | -0.02 (-0.37, 0.33) | -0.16 (-0.38, 0.06) | -0.15 (-0.36, 0.06) | -0.04 (-0.39, 0.32) | -0.14 (-0.37, 0.09) |
| *te + c-M + s-M + px_ed* | 0.04 (-0.15, 0.23) | 0.04 (-0.14, 0.23) | 0.03 (-0.16, 0.22) | 0.05 (-0.14, 0.24) | 0.08 (-0.12, 0.27) | 0.06 (-0.14, 0.26) | 0.07 (-0.13, 0.27) |
| *te + c-M + s-M + px_ed + mot_int* | -0.73 (-1.5, 0.05) | 0.05 (-0.35, 0.44) | 0.04 (-0.36, 0.43) | 0.05 (-0.35, 0.45) | 0.06 (-0.33, 0.46) | 0.05 (-0.35, 0.45) | 0.07 (-0.34, 0.47) |
| *te + c-M + s-M + px_re + px_ed* | -0.45 (-1.23, 0.32) | 0.30 (-0.04, 0.63) | 0.25 (-0.11, 0.6) | 0.28 (-0.07, 0.63) | 0.27 (-0.07, 0.61) | 0.23 (-0.13, 0.59) | 0.29 (-0.07, 0.64) |
| *te + EPR + re + AF + c-M + s-M + px_ed* | 0.03 (-0.26, 0.33) | 0.03 (-0.43, 0.50) | 0.03 (-0.43, 0.50) | 0.03 (-0.43, 0.50) | 0.03 (-0.43, 0.05) | 0.04 (-0.44, 0.50) | 0.04 (-0.44, 0.51) |
| *te + EPR + re + c-M + s-M* | -0.09 (-0.29, 0.12) | 0.04 (-0.29, 0.37) | 0.03 (-0.31, 0.36) | 0.02 (-0.34, 0.38) | -0.09 (-0.50, 0.33) | -0.09 (-0.50, 0.33) | -0.09 (-0.51, 0.34) |
| *te + px_ed* | 0.00 (-0.16, 0.16) | -0.12 (-0.35, 0.1) | -0.14 (-0.37, 0.09) | -0.12 (-0.35, 0.11) | -0.12 (-0.34, 0.11) | -0.13 (-0.36, 0.10) | -0.12 (-0.35, 0.11) |
| *te + sta + c-M + s-M + px_ed* | -0.10 (-0.24, 0.04) | -0.09 (-0.29, 0.11) | -0.11 (-0.31, 0.10) | -0.09 (-0.29, 0.12) | -0.07 (-0.28, 0.13) | -0.09 (-0.30, 0.12) | -0.08 (-0.29, 0.13) |
| *te + sta + c-M + s-M + px_ed + px_fin* | 0.02 (-0.42, 0.47) | 0.02 (-0.55, 0.59) | 0.02 (-0.55, 0.60) | 0.02 (-0.55, 0.60) | 0.02 (-0.55, 0.59) | 0.02 (-0.55, 0.60) | 0.02 (-0.56, 0.60) |
| *te + sta + fin + c-M + s-M + px_ed* | 0.10 (-0.05, 0.24) | 0.10 (-0.29, 0.48) | 0.10 (-0.29, 0.48) | 0.10 (-0.29, 0.49) | 0.10 (-0.29, 0.48) | 0.10 (-0.30, 0.49) | 0.10 (-0.30, 0.49) |
| **Component vs UC** | **NMA** | **Additive CNMA** | **Interaction CNMA  (Model 2)** | **Interaction CNMA  (Model 5)** | **Interaction CNMA  (Model 7)** | **Interaction CNMA  (Model 14)** | **Interaction CNMA  (Model 15)** |
| *AF* | -- | -0.02 (-0.58, 0.54) | 0.03 (-0.54, 0.60) | 0.01 (-0.58, 0.60) | -0.20 (-0.86, 0.45) | -0.14 (-0.82, 0.54) | -0.25 (-1.04, 0.55) |
| *c-M* | -- | -0.03 (-0.22, 0.16) | 0.11 (-0.25, 0.48) | -0.04 (-0.26, 0.17) | -0.03 (-0.22, 0.16) | 0.10 (-0.28, 0.47) | -0.02 (-0.24, 0.20) |
| *clin_re* | -- | -0.19 (-0.79, 0.40) | -0.14 (-0.75, 0.47) | -0.27 (-1.00, 0.47) | -0.21 (-0.81, 0.39) | -0.16 (-0.78, 0.45) | -0.17 (-0.94, 0.60) |
| *EPR* | -- | 0.05 (-0.34, 0.43) | 0.03 (-0.36, 0.42) | 0.03 (-0.39, 0.44) | 0.14 (-0.28, 0.56) | 0.12 (-0.32, 0.55) | 0.16 (-0.34, 0.67) |
| *fin* | -- | 0.19 (-0.25, 0.62) | 0.20 (-0.24, 0.64) | 0.18 (-0.26, 0.62) | 0.17 (-0.27, 0.61) | 0.19 (-0.26, 0.63) | 0.17 (-0.28, 0.62) |
| *mot_int* | -- | 0.00 (-0.37, 0.37) | 0.01 (-0.36, 0.38) | 0.00 (-0.38, 0.37) | -0.01 (-0.38, 0.36) | -0.01 (-0.38, 0.37) | -0.01 (-0.39, 0.37) |
| *px_ed* | -- | 0.01 (-0.17, 0.19) | -0.02 (-0.22, 0.17) | 0.00 (-0.19, 0.19) | -0.04 (-0.25, 0.17) | -0.07 (-0.28, 0.15) | -0.04 (-0.25, 0.17) |
| *px_fin* | -- | 0.11 (-0.49, 0.72) | 0.13 (-0.48, 0.74) | 0.11 (-0.50, 0.72) | 0.10 (-0.51, 0.7) | 0.11 (-0.50, 0.72) | 0.10 (-0.52, 0.71) |
| *px_re* | -- | 0.25 (-0.05, 0.56) | 0.22 (-0.10, 0.53) | 0.35 (-0.29, 1.00) | 0.20 (-0.13, 0.52) | 0.17 (-0.16, 0.51) | 0.13 (-0.67, 0.92) |
| *re* | -- | -0.04 (-0.37, 0.29) | -0.06 (-0.39, 0.27) | -0.05 (-0.40, 0.29) | 0.02 (-0.33, 0.37) | 0.00 (-0.36, 0.36) | 0.04 (-0.35, 0.44) |
| *s-M* | -- | 0.19 (0.03, 0.36) | 0.25 (0.05, 0.45) | 0.21 (0.02, 0.41) | 0.22 (0.05, 0.40) | 0.27 (0.06, 0.48) | 0.22 (0.02, 0.42) |
| *sta* | -- | -0.14 (-0.33, 0.06) | -0.14 (-0.33, 0.06) | -0.14 (-0.33, 0.06) | -0.15 (-0.35, 0.05) | -0.15 (-0.35, 0.05) | -0.15 (-0.35, 0.05) |
| *te* | -- | -0.13 (-0.33, 0.06) | -0.11 (-0.32, 0.09) | -0.12 (-0.33, 0.09) | -0.44 (-1.07, 0.18) | -0.39 (-1.04, 0.25) | -0.49 (-1.26, 0.29) |
| *cM.sM* | -- | -- | -0.20 (-0.63, 0.23) | -- | -- | -0.17 (-0.61, 0.27) | -- |
| *sM.pxre* | -- | -- | -- | -0.12 (-0.82, 0.58) | -- | -- | 0.08 (-0.74, 0.90) |
| *te.pxed* | -- | -- | -- | -- | 0.36 (-0.33, 1.06) | 0.33 (-0.39, 1.04) | 0.41 (-0.42, 1.23) |
| τ^2^ | 0.00 | 0.04 | 0.04 | 0.04 | 0.04 | 0.04 | 0.04 |
| I^2^ | 19% | 69% | 70% | 70% | 69% | 70% | 70% |
| Q, df, p-value | 11.11, 9, 0.268 | 77.82, 24 <0.001 | 75.45, 23 <0.001 | 75.39, 23 <0.001 | 73.90, 23, <0.001 | 72.43, 22, <0.001 | 73.49, 22, <0.001 |
| Difference in Q statistics (CNMA and NMA model) | -- | 66.71, 15, <0.001 | 64.34, 14, <0.001 | 64.28, 14, <0.001 | 62.79, 14, <0.001 | 61.32, 13, <0.001 | 62.38, 13, <0.001 |

### Supplementary File References
