## Supplementary material for "Analysing complex interventions using component network meta-analysis": R code example

### CNMA Examples

2025-05-30

##### Load necessary libraries and auxiliary function

The R package **netmeta** will be used to conduct standard and component NMA. The other R packages are used in import the data sets or to generate figures.

```
library("readxl")
library("viscomp")
library("ggplot2")
library("gridExtra")
library("netmeta")
```

```
## Loading required package: meta

## Loading required package: metadat

## Loading 'meta' package (version 8.1-0).
## Type 'help(meta)' for a brief overview.

## Loading 'netmeta' package (version 3.2-0).
## Type 'help("netmeta-package")' for a brief overview.
```

```
suppressMessages(library("dplyr"))
library("stringr")
library("tibble")
#
# Print network meta-analysis results with three digits and various settings
# for heterogeneity statistics
#
settings.meta(digits = 3,
  digits.Q = 2, digits.tau2 = 3, digits.I2 = 0, digits.pval.Q = 3)
#
# Extent width from 80 to 100 characters
#
options(width = 100)
```

***Note, the version of netmeta must be 3.0-0 or greater.***

In addition, we load some auxiliary R functions.

```
source("funcs.R")
```

### 1. Binary outcome example: Prehabilitation

Before running the CNMA analyses, we import the data set and fit a standard NMA. The workflow is very similar for all three data sets.

We start by defining specific settings for the prehabilitation outcome.

```
Sheet <- Name <- "Prehabilitation"
#
Title <- paste0("Component and Standard NMA\n(", Name, ")")
TitleNMA <- paste0("Standard NMA\n(", Name, ")")
TitleCNMA <- paste0("CNMA: Treatment-level effects\n(", Name, ")")
TitleCompCNMA <- paste0("CNMA: Component-level effects\n(", Name, ")")
#
EffectSize <- "OR"
SmallVal <- "desirable"
#
Reference <- "UC"
#
left.int <- "Intervention"
left.comp <- "Component"
right.int <- right.comp <- "Usual Care"
#
LeftLabelInt <- paste("Favours", left.int)
LeftLabelComp <- paste("Favours", left.comp)
RightLabelInt <- paste("Favours", right.int)
RightLabelComp <- paste("Favours", right.comp)
```

#### 1.1 Transform data set to comparison-based format

We import the dataset, i.e., a sheet in the provided Excel file 'CNMA_data.xlsx' and transform the dataset into a format required by *netmeta()* to run the standard NMA.

```
dat_preh <- read_xlsx("CNMA_data.xlsx", sheet = Sheet)
#
pw_preh <- pairwise(list(t1, t2, t3),
  event = list(r1, r2, r3), n = list(n1, n2, n3),
  data = dat_preh, studlab = refid,
  sm = EffectSize,
  reference.group = Reference,
  append = FALSE)
```

Then, we check the connectivity of the network.

```
netconnection(pw_preh)
```

```
## Number of studies: k = 106
## Number of pairwise comparisons: m = 108
## Number of treatments: n = 8
## Number of designs: d = 8
## Number of networks: 1
```

We cannot conduct a standard NMA if the number of subnetworks is different from 1 indicating a disconnected network.

#### 1.2 Fit standard NMA

Finally, we conduct the standard NMA as the network is connected.

```
nma_preh <- netmeta(pw_preh, common = FALSE, small.values = SmallVal)
```

##### Network diagram

From the network diagram, we see that the network is basically a star-shaped network. The only loop exists due to a single three-arm study (indicated by the blue area) comparing UC, EXE, and EXE+PSY.

```
par(mar = c(5.1, 4.1, 4.1, 10.1))
netgraph(nma_preh, points = TRUE,
  multiarm = TRUE, col.multiarm = "blue", thickness = "equal", seq = "optimal")
```

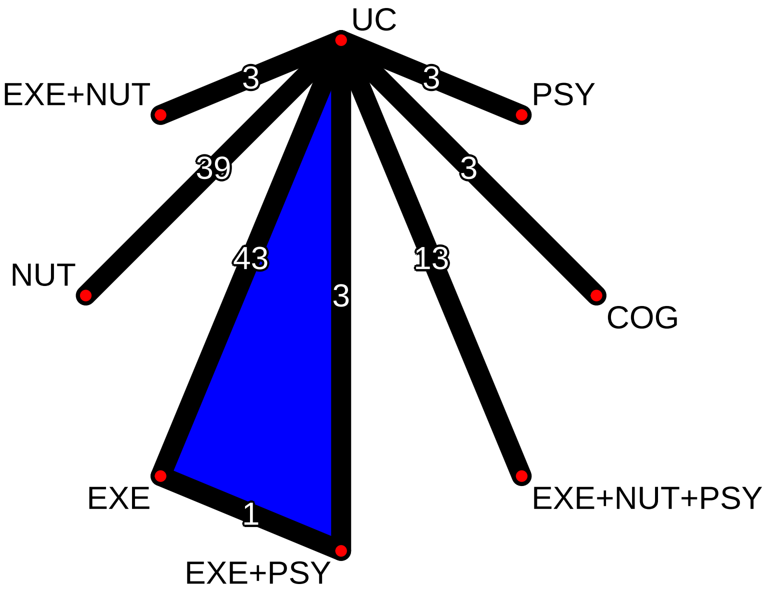


##### Rank treatments

Rank treatments by P-scores:

```
netrank(nma_preh)
```

```
##             P-score
## EXE          0.8526
## EXE+NUT      0.7537
## NUT          0.6219
## EXE+NUT+PSY  0.5895
## EXE+PSY      0.4412
## COG          0.3133
## PSY          0.2387
## UC           0.1890
```

##### Standard NMA forest plot

Generate a forest plot of the standard NMA effect estimates sorted by P-scores:

```
xlim_vals <-
  range(nma_preh$lower.nma.random, nma_preh$upper.nma.random, na.rm = TRUE)
#
```

```
# Confidence limits are stored on the log scale for the odds ratio as effect
# measure; limits for the x-axis in the forest plot must be back-transformed
#
xlim_vals <- round(exp(xlim_vals), 2)
```

```
forest(nma_preh, sortvar = -Pscore,
  smlab = TitleNMA,
  label.left = LeftLabelInt, label.right = RightLabelInt,
  leftcols = c("studlab", "k"),
  leftlabs =
    c(paste("Contrast to", nma_preh$reference.group), "Direct\nComparisons"),
  xlim = xlim_vals)
```

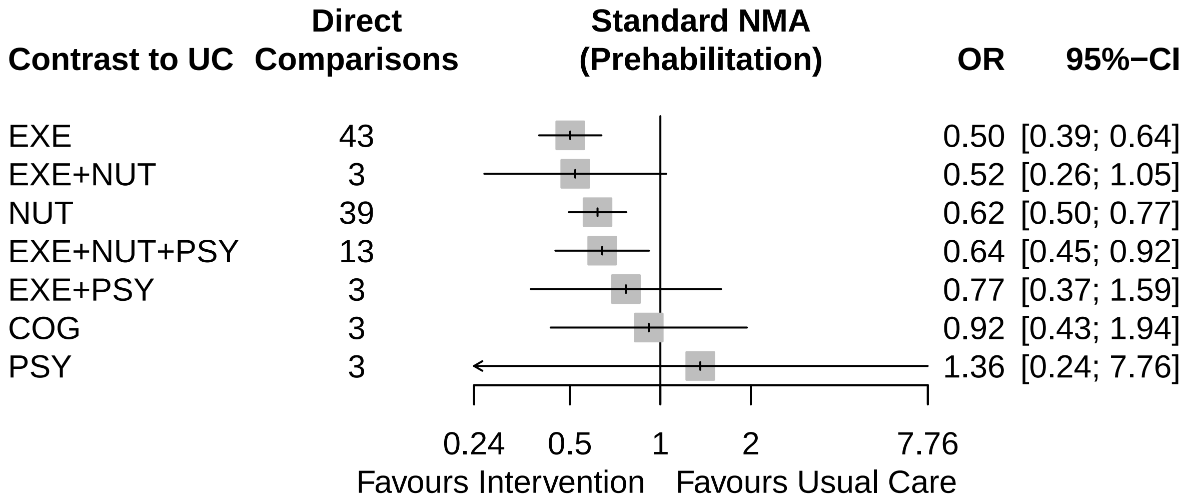


##### 1.3 Fit the additive CNMA model

The reference treatment from the standard NMA is defined as an inactive component in the additive CNMA model.

```
cnma_preh <- netcomb(nma_preh, inactive = nma_preh$reference.group)
cnma_preh
```

```
## Number of studies: k = 106
## Number of pairwise comparisons: m = 108
## Number of treatments: n = 8
## Number of active components: c = 4
## Number of designs: d = 8
##
## Random effects model (inactive component: 'UC')
##
## Treatment estimate (sm = 'OR', comparison: other treatments vs 'UC'):
##                  OR          95%-CI     z  p-value
## COG           0.912 [0.432; 1.925] -0.24   0.8093
## EXE           0.530 [0.423; 0.663] -5.55 < 0.0001
## EXE+NUT       0.348 [0.262; 0.463] -7.26 < 0.0001
## EXE+NUT+PSY 0.608 [0.442; 0.835] -3.07   0.0021
## EXE+PSY       0.925 [0.650; 1.316] -0.43   0.6642
```

```
## NUT         0.657 [0.536; 0.805] -4.05 < 0.0001
## PSY         1.746 [1.169; 2.606]  2.72   0.0064
## UC              .              .     .        .
##
## Incremental effect for existing combinations:
##               iOR         95%-CI     z  p-value
## EXE+NUT     0.348 [0.262; 0.463] -7.26 < 0.0001
## EXE+NUT+PSY 0.608 [0.442; 0.835] -3.07   0.0021
## EXE+PSY     0.925 [0.650; 1.316] -0.43   0.6642
##
## Incremental effect for components:
##       iOR         95%-CI     z  p-value
## COG 0.912 [0.432; 1.925] -0.24   0.8093
## EXE 0.530 [0.423; 0.663] -5.55 < 0.0001
## NUT 0.657 [0.536; 0.805] -4.05 < 0.0001
## PSY 1.746 [1.169; 2.606]  2.72   0.0064
##
## Quantifying heterogeneity / inconsistency:
## tau^2 = 0.141; tau = 0.3752; I^2 = 30% [11%; 45%]
##
## Heterogeneity statistics:
##                     Q  df p-value
## Additive model 147.72 103   0.003
## Standard model 144.20 100   0.003
## Difference       3.52   3   0.318
##
## Details of network meta-analysis methods:
## - Frequentist graph-theoretical approach
## - Component network meta-analysis
## - DerSimonian-Laird estimator for tau^2
## - Calculation of I^2 based on Q
```

##### Check the additivity assumption

The additivity assumption can be assessed by comparing the Q statistics of the additive CNMA and the standard NMA model. A p-value greater than the appropriate cut-off (based on the difference in degrees of freedom between the two models) suggests that the additivity assumption is met.

The difference between the Q statistics for the additive CNMA and standard NMA is 3.52 with 3 degrees of freedom resulting in a p-value well above the cut-off value, suggesting that the additivity assumption is met.

##### Rank treatments

Rank treatments by P-scores:

```r
netrank(cnma_preh)
```

```
##             P-score
## EXE+NUT      0.9982
## EXE          0.7965
## EXE+NUT+PSY  0.6784
## NUT          0.5915
## COG          0.3576
## EXE+PSY      0.3200
## UC           0.2478
## PSY          0.0100
```

##### Additive CNMA forest plots

First, generate a forest plot of all treatment-level effects:

```
xlim_vals <-
  range(cnma_preh$lower.cnma.random, cnma_preh$upper.cnma.random, na.rm = TRUE)
#
xlim_vals <- round(exp(xlim_vals), 2)
```

```
forest(cnma_preh, sortvar = -TE,
  smlab = TitleCNMA,
  label.left = LeftLabelInt, label.right = RightLabelInt,
  leftcols = c("studlab", "k"),
  leftlabs =
    c(paste("Contrast to", cnma_preh$reference.group), "Direct\nComparisons"),
  xlim = xlim_vals)
```

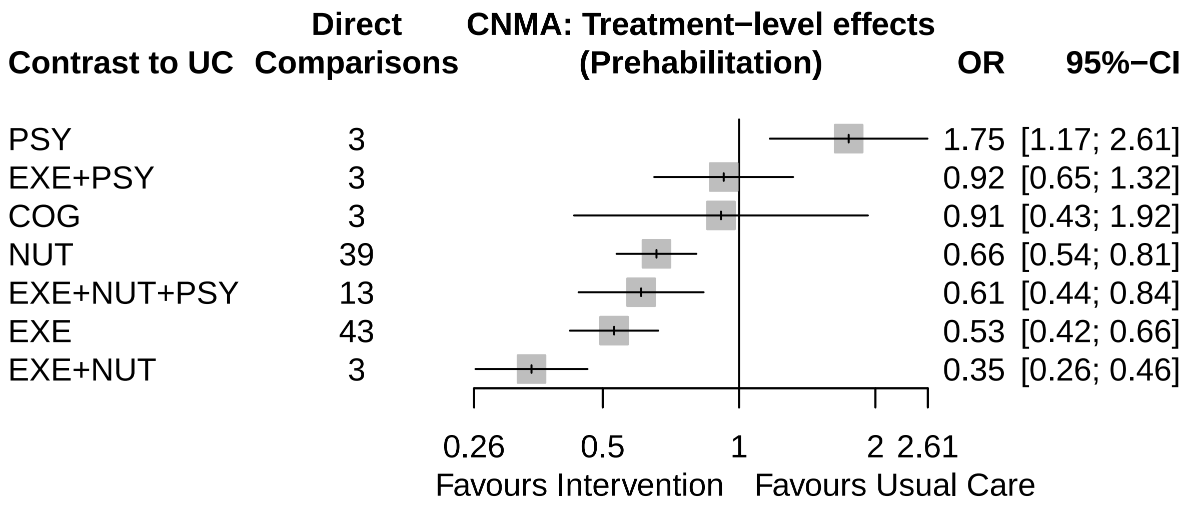


Next, generate a forest plot of all component-level effects:

```
# Component additive CNMA forest plot
forest(netcomplex(cnma_preh, cnma_preh$comps),
  smlab = TitleCompCNMA,
  label.left = LeftLabelComp, label.right = RightLabelComp,
  xlim = xlim_vals)
```

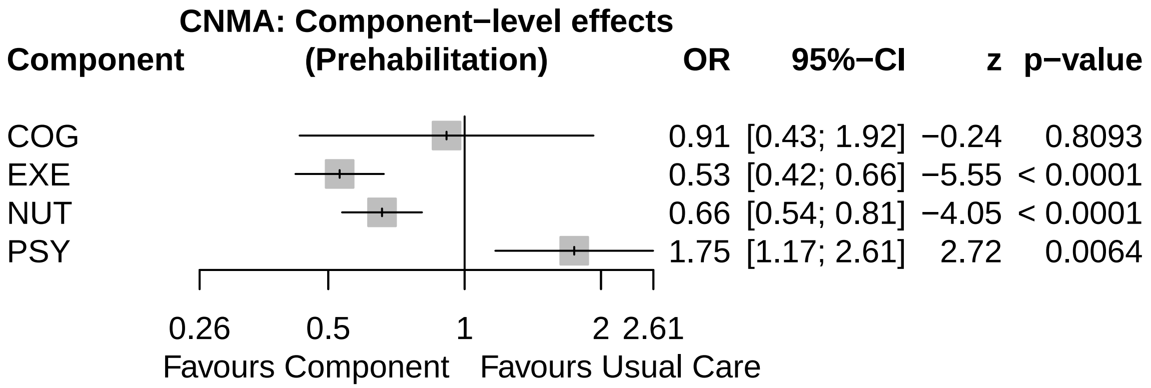


Finally, generate a forest plot comparing the results of the standard NMA and CNMA models. The font size, spacing, and square size can be adjusted to fit the figure to the page.

```
nb_preh <- netbind(cnma_preh, nma_preh,
  name = c("Additive CNMA", "Standard NMA"),
  col.study = c("red", "black"), col.square = c("red", "black"))
#
forest(nb_preh,
  smlab = Title,
  label.left = LeftLabelInt, label.right = RightLabelInt,
  addrow.subgroups = FALSE)
```

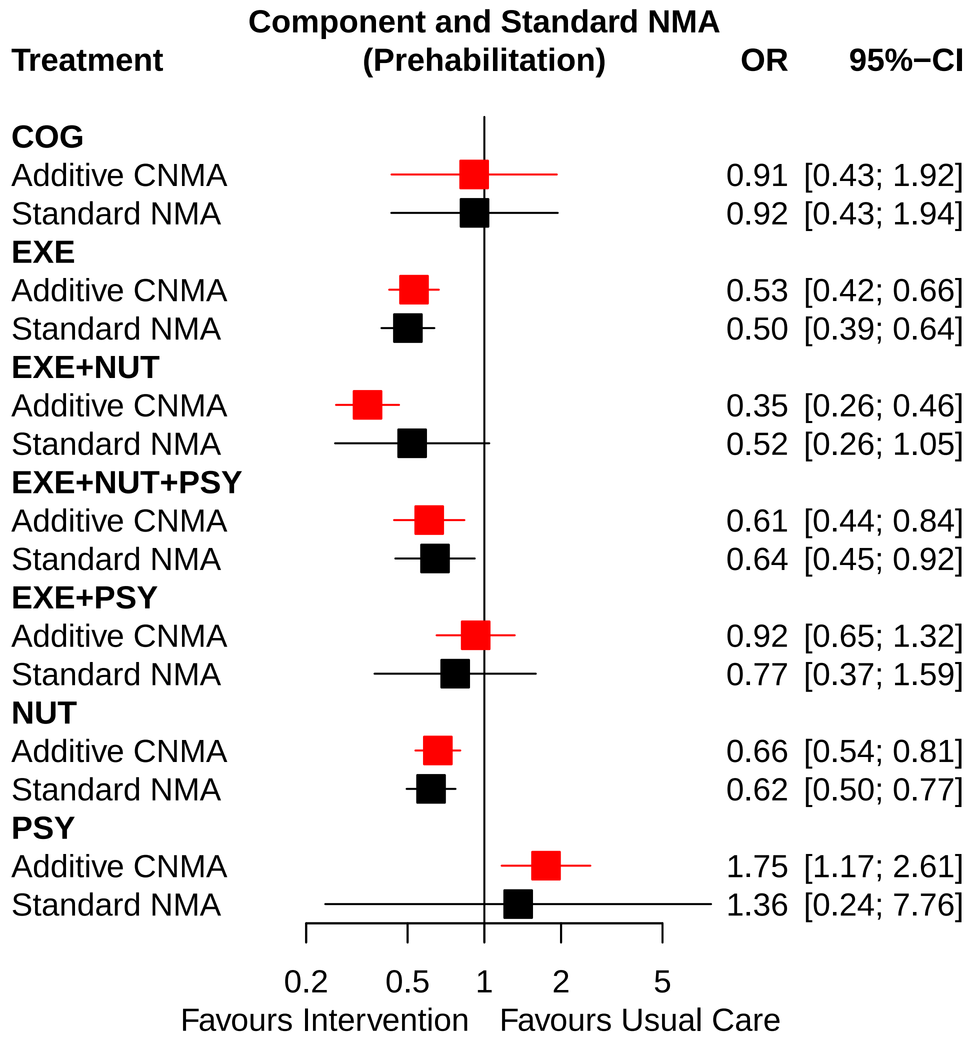


#### 1.4 Model selection of interaction CNMA models

When the additivity assumption is not met, interactions can be introduced to the additive model and the new models assessed for fit. When the additive model was run originally, a C matrix of all treatments and components was generated, where each row is a treatment and columns represent each component. Ones and zeros indicate whether a treatment row contains each of the components in the columns.

The C matrix of the prehabilitation CNMA is given by

```
cnma_preh$C.matrix
```

```
##              COG EXE NUT PSY
## COG            1   0   0   0
## EXE            0   1   0   0
## EXE+NUT        0   1   1   0
## EXE+NUT+PSY    0   1   1   1
## EXE+PSY        0   1   0   1
## NUT            0   0   1   0
## PSY            0   0   0   1
```

```
## UC              0   0   0   0
```

The CNMA contains eight interventions (rows of the C matrix) and four active components (columns).

To fit an interaction model, *netcomb()* is used, but the C matrix of the additive model is replaced with a new C matrix that contains an extra column representing the interaction term.

The auxiliary function *cnma_model_fit()* can be used to fit all interaction CNMA models with a single 2-way (or higher) interaction term.

For the prehabilitation dataset only two 2-way interaction terms exist.

```
cnma_model_fit(cnma_preh, n = 2)
```

```
##  interaction      Q  df  pval  tau2  I2 Q.nma df.nma pval.nma Q.add df.add pval.add
##    EXE * NUT 144.69 102 0.004 0.137 30%  0.49      2    0.784  3.03      1    0.082
##    EXE * PSY 147.64 102 0.002 0.144 31%  3.44      2    0.179  0.08      1    0.771
```

And only a single three-way interaction.

```
cnma_model_fit(cnma_preh, n = 3)
```

```
##      interaction      Q  df  pval  tau2  I2 Q.nma df.nma pval.nma Q.add df.add pval.add
##  EXE * NUT * PSY 147.43 102 0.002 0.145 31%  3.23      2    0.199  0.29      1    0.588
```

#### 1.5 Build the interaction CNMA models

Based on the model fit statistics generated for each interaction model above, we have selected the 'best-fitting' models (i.e., those with Q statistics significantly lower than that of the additive model). In a forward selection process, we will build upon these models, generating models with multiple interaction terms. We will then test each model's fit compared to that of its nested lower-order interaction model.

##### Create C matrices

First, we create a vector with all available combinations of two components using *combinations()* which was already used internally in the auxiliary function *cnma_model_fit()*. Interaction terms can only be added to an additive CNMA if the combination is available.

```
combs2_preh <- combinations(cnma_preh, n = 2)
combs2_preh
```

```
## [1] "EXE+NUT" "EXE+PSY"
```

Next, we create the C matrices C1_preh and C2_preh for the single 2-way interactions.

```
# Interaction model 1: EXE * NUT
C1_preh <- createC(cnma_preh, combs2_preh[1])
# Interaction model 2: EXE * PSY
C2_preh <- createC(cnma_preh, combs2_preh[2])
```

and C matrix C3_preh with the two 2-way interactions.

```
# Interaction model 3: EXE * NUT and EXE * PSY
C3_preh <- createC(cnma_preh, combs2_preh)
```

We see that the C matrix C3_preh has two additional columns.

```
prmatrix(C3_preh)
```

```
##              COG EXE NUT PSY EXE * NUT EXE * PSY
## COG            1   0   0   0         0         0
## EXE            0   1   0   0         0         0
```

```
## EXE+NUT        0   1   1   0          1          0
## EXE+NUT+PSY    0   1   1   1          1          1
## EXE+PSY        0   1   0   1          0          1
## NUT            0   0   1   0          0          0
## PSY            0   0   0   1          0          0
## UC             0   0   0   0          0          0
```

Next, we will manually create C matrix C4_preh with the two 2-way interactions and the only three-way interaction.

```
# Interaction model 4: EXE * NUT + EXE * PSY + EXE * NUT * PSY
C4_preh <- createC(C3_preh, combinations(cnma_preh, n = 3))
prmatrix(C4_preh)
```

```
##               COG EXE NUT PSY EXE * NUT EXE * PSY EXE * NUT * PSY
## COG             1   0   0   0         0         0               0
## EXE             0   1   0   0         0         0               0
## EXE+NUT         0   1   1   0         1         0               0
## EXE+NUT+PSY     0   1   1   1         1         1               1
## EXE+PSY         0   1   0   1         0         1               0
## NUT             0   0   1   0         0         0               0
## PSY             0   0   0   1         0         0               0
## UC              0   0   0   0         0         0               0
```

```
cnma_model_fit(cnma_preh, C1_preh, C2_preh, C3_preh, C4_preh, selection = TRUE)
```

```
##                model Q.diff df.diff pval.diff
##  Interaction model 1   3.03       1     0.082
##  Interaction model 2   0.08       1     0.771
##  Interaction model 3   3.03       2     0.219
##  Interaction model 4   3.52       3     0.318
```

##### Compare model fit with previous nested interaction model

We also must compare model fit with each multi-term interaction model's previous nested (single-interaction) model with the lowest Q statistic. First, we will identify the previous nested model with the lowest Q statistic, then generate model fit statistics:

```
cnma_model_selection(cnma_preh, C1_preh, C2_preh, C3_preh, C4_preh)
```

```
##                model        nested model Q.diff df  pval
##  Interaction model 3 Interaction model 1   0.00  1 0.975
##  Interaction model 4 Interaction model 1   0.49  2 0.784
```

#### 1.6 Present standard NMA and interaction cNMA model (with usual care as reference)

```
nb_preh_ia <- netbind(cnma_preh,
  netcomb(nma_preh, C.matrix = C1_preh, inactive = nma_preh$reference.group),
  nma_preh,
  name = c("Additive CNMA", "Interaction CNMA",
           "Standard NMA"),
  col.study = c("red", "blue", "black"),
  col.square = c("red", "blue", "black"))
#
forest(nb_preh_ia,
```

```
xlim = c(0.15, 10), at = c(0.25, 0.5, 1, 2, 5, 10),
reference.group = "UC",
label.left = "Favours other",
label.right = "Favours Usual Care",
addrow.subgroups = FALSE)
```

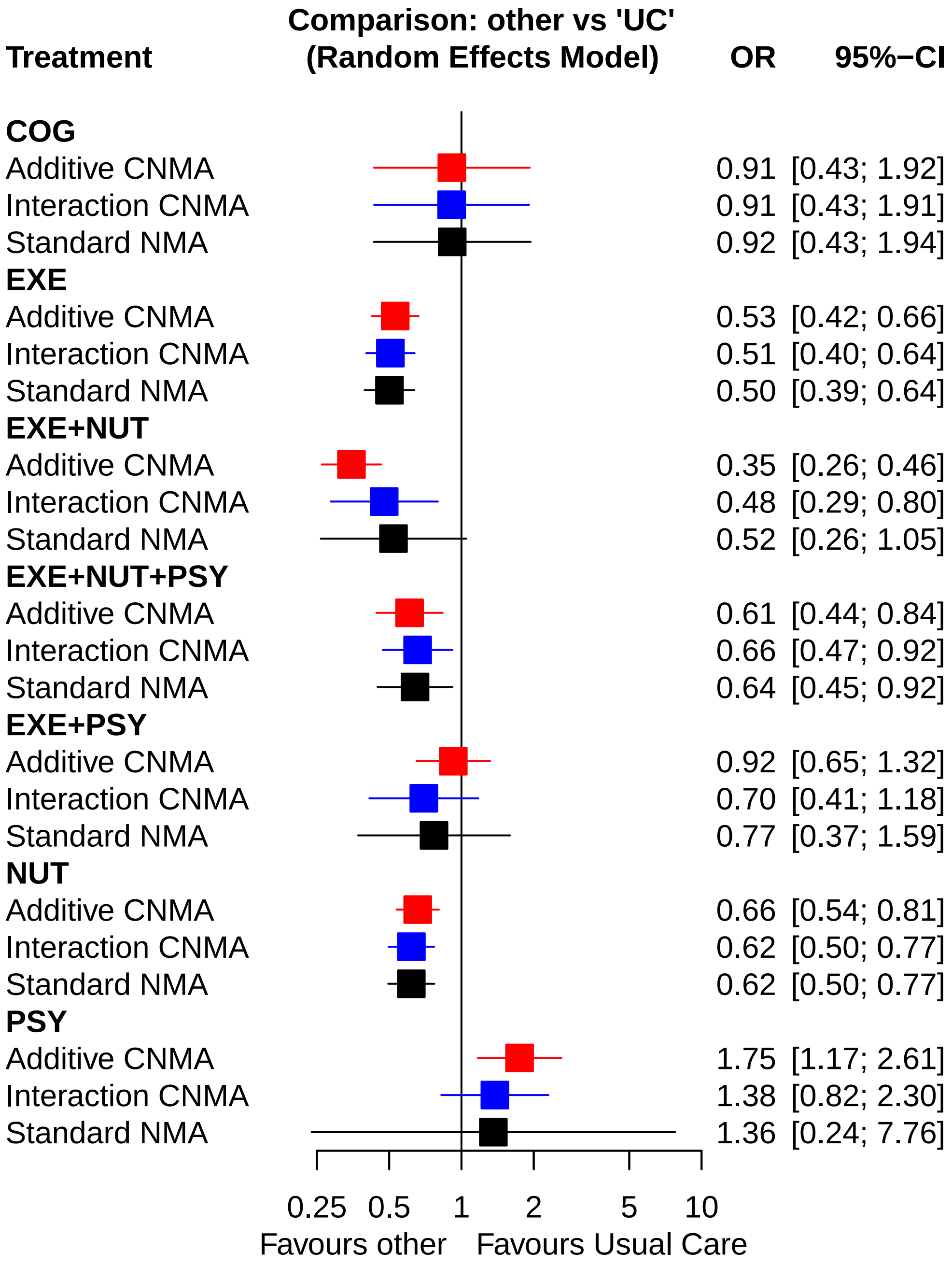

### 2. Binary outcome example: Fractures

We have to change the sheet and outcome name.

```
Sheet <- Name <- "Fractures"
#
Title <- paste0("Component and Standard NMA\n(", Name, ")")
TitleNMA <- paste0("Standard NMA\n(", Name, ")")
TitleCNMA <- paste0("CNMA: Treatment-level effects\n(", Name, ")")
TitleCompCNMA <- paste0("CNMA: Component-level effects\n(", Name, ")")
```

#### 2.1 Transform data set to comparison-based format

```
dat_frac <- read_xlsx("CNMA_data.xlsx", sheet = Sheet)
#
pw_frac <- pairwise(list(t1, t2, t3, t4),
  event = list(r1, r2, r3, r4), n = list(n1, n2, n3, n4),
  data = dat_frac, studlab = refid,
  sm = EffectSize,
  reference.group = Reference,
  append = FALSE)
```

Check the connectivity of the network.

```
netconnection(pw_frac)
```

```
## Number of studies: k = 68
## Number of pairwise comparisons: m = 89
## Number of treatments: n = 44
## Number of designs: d = 49
## Number of networks: 1
```

#### 2.2 Fit standard NMA

Finally, we conduct the standard NMA as the network is connected.

```
nma_frac <- netmeta(pw_frac, common = FALSE, small.values = SmallVal)
```

##### Network diagram

```
par(mar = c(5.1, 4.1, 4.1, 10.1))
netgraph(nma_frac, points = TRUE,
  multiarm = TRUE, col.multiarm = "blue", thickness = "equal", seq = "optimal")
```

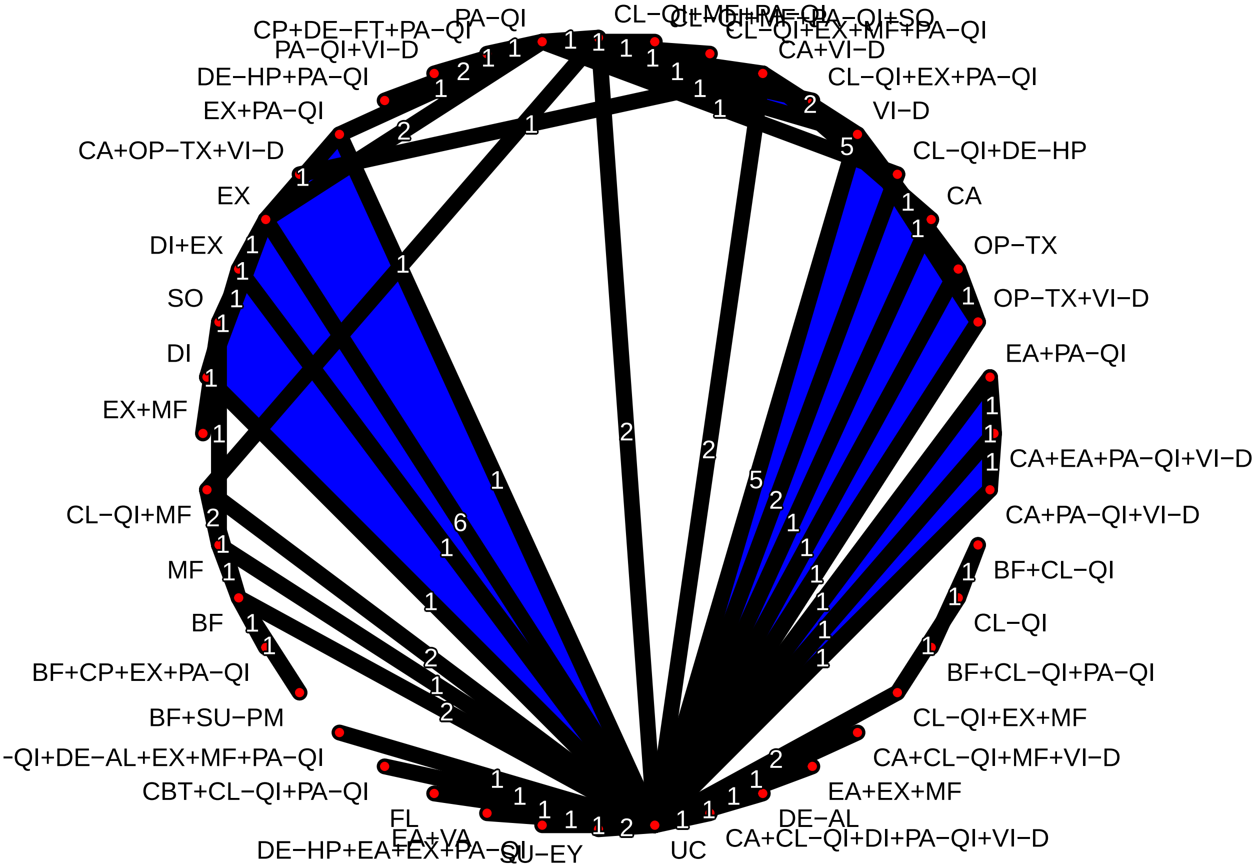


##### Rank treatments

Rank treatments by P-scores:

```
netrank(nma_frac)
```

```
##                            P-score
## BF+CP+EX+PA-QI              0.9141
## CA+OP-TX+VI-D               0.9128
## FL                          0.7838
## EX+MF                       0.7494
## CBT+CL-QI+PA-QI             0.7425
## EA+EX+MF                    0.7146
## DI+EX                       0.7019
## CA+CL-QI+DI+PA-QI+VI-D      0.6793
## SU-EY                       0.6611
## CA+CL-QI+MF+VI-D            0.6543
## EX                          0.6498
## DE-HP+PA-QI                 0.6098
## BF+CL-QI+PA-QI              0.5995
## CL-QI+DE-AL+EX+MF+PA-QI     0.5723
## BF+SU-PM                    0.5635
## DE-AL                       0.5588
## CA+VI-D                     0.5443
## DI                          0.5347
## CA+PA-QI+VI-D               0.5269
## EX+PA-QI                    0.5231
```

```
## CA+EA+PA-QI+VI-D         0.5198
## EA+PA-QI                 0.5033
## OP-TX+VI-D               0.5014
## CL-QI+MF+PA-QI           0.4835
## BF                       0.4712
## DE-HP+EA+EX+PA-QI        0.4698
## CA                       0.4536
## PA-QI+VI-D               0.4376
## CL-QI+EX+MF+PA-QI        0.4322
## MF                       0.4247
## CL-QI+MF+PA-QI+SO        0.4114
## PA-QI                    0.3825
## BF+CL-QI                 0.3613
## CL-QI                    0.3587
## UC                       0.3499
## CL-QI+DE-HP              0.3222
## SO                       0.3141
## CL-QI+EX+MF              0.3067
## VI-D                     0.3031
## CL-QI+MF                 0.2798
## CP+DE-FT+PA-QI           0.2333
## CL-QI+EX+PA-QI           0.2255
## OP-TX                    0.1355
## EA+VA                    0.1228
```

##### Standard NMA forest plot

Generate a forest plot of the standard NMA effect estimates sorted by P-scores:

```r
xlim_vals <-
  range(nma_frac$lower.nma.random, nma_frac$upper.nma.random, na.rm = TRUE)
#
# Confidence limits are stored on the log scale for the odds ratio as effect
# measure; limits for the x-axis in the forest plot must be back-transformed
#
xlim_vals <- round(exp(xlim_vals), 2)
```

```r
forest(nma_frac, sortvar = -Pscore,
  smlab = TitleNMA,
  label.left = LeftLabelInt, label.right = RightLabelInt,
  leftcols = c("studlab", "k"),
  leftlabs =
    c(paste("Contrast to", nma_frac$reference.group), "Direct\nComparisons"),
  xlim = xlim_vals)
```

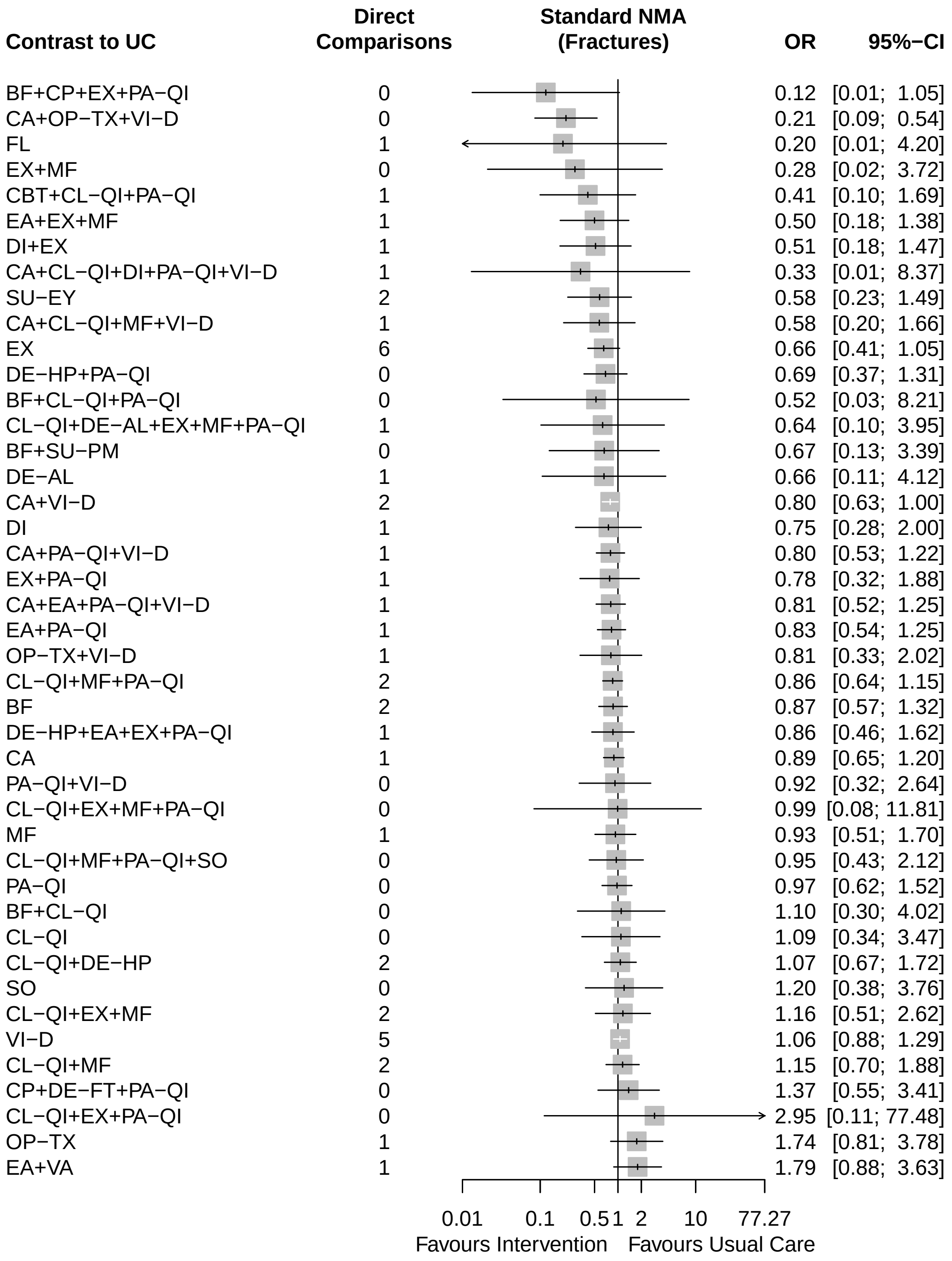


#### 2.3 Fit the additive CNMA model

The reference treatment from the standard NMA is defined as an inactive component in the additive CNMA model.

```r
cnma_frac <- netcomb(nma_frac, inactive = nma_frac$reference.group)
cnma_frac
```

```
## Number of studies: k = 68
## Number of pairwise comparisons: m = 89
## Number of treatments: n = 44
## Number of active components: c = 20
## Number of designs: d = 49
##
## Random effects model (inactive component: 'UC')
##
## Treatment estimate (sm = 'OR', comparison: other treatments vs 'UC'):
##                              OR           95%-CI     z p-value
## BF                        0.871 [0.637; 1.191] -0.87  0.3869
## BF+CL-QI                  1.028 [0.659; 1.601]  0.12  0.9045
## BF+CL-QI+PA-QI            0.970 [0.608; 1.549] -0.13  0.8999
## BF+CP+EX+PA-QI            0.119 [0.014; 1.024] -1.94  0.0525
## BF+SU-PM                  0.669 [0.136; 3.289] -0.49  0.6212
## CA                        0.832 [0.692; 1.000] -1.96  0.0499
## CA+CL-QI+DI+PA-QI+VI-D    0.716 [0.305; 1.679] -0.77  0.4419
## CA+CL-QI+MF+VI-D          0.807 [0.600; 1.085] -1.42  0.1556
## CA+EA+PA-QI+VI-D          0.754 [0.552; 1.029] -1.78  0.0755
## CA+OP-TX+VI-D             0.599 [0.344; 1.044] -1.81  0.0706
## CA+PA-QI+VI-D             0.785 [0.605; 1.019] -1.82  0.0688
## CA+VI-D                   0.831 [0.704; 0.981] -2.19  0.0287
## CBT+CL-QI+PA-QI           0.410 [0.101; 1.668] -1.25  0.2129
## CL-QI                     1.180 [0.831; 1.675]  0.92  0.3559
## CL-QI+DE-AL+EX+MF+PA-QI 0.573 [0.156; 2.099] -0.84  0.4004
## CL-QI+DE-HP               1.009 [0.709; 1.435]  0.05  0.9617
## CL-QI+EX+MF               0.827 [0.592; 1.155] -1.12  0.2647
## CL-QI+EX+MF+PA-QI         0.781 [0.548; 1.113] -1.37  0.1719
## CL-QI+EX+PA-QI            0.949 [0.553; 1.629] -0.19  0.8493
## CL-QI+MF                  0.971 [0.769; 1.226] -0.25  0.8039
## CL-QI+MF+PA-QI            0.917 [0.722; 1.165] -0.71  0.4782
## CL-QI+MF+PA-QI+SO         1.041 [0.584; 1.856]  0.14  0.8907
## CP+DE-FT+PA-QI            1.333 [0.595; 2.988]  0.70  0.4850
## DE-AL                     0.734 [0.200; 2.688] -0.47  0.6400
## DE-HP+EA+EX+PA-QI         0.661 [0.440; 0.993] -1.99  0.0464
## DE-HP+PA-QI               0.808 [0.542; 1.203] -1.05  0.2929
## DI                        0.773 [0.364; 1.640] -0.67  0.5019
## DI+EX                     0.658 [0.289; 1.502] -0.99  0.3203
## EA+EX+MF                  0.673 [0.434; 1.044] -1.77  0.0773
## EA+PA-QI                  0.907 [0.692; 1.189] -0.70  0.4813
## EA+VA                     1.790 [0.904; 3.544]  1.67  0.0946
## EX                        0.852 [0.638; 1.136] -1.09  0.2753
## EX+MF                     0.701 [0.479; 1.026] -1.83  0.0676
## EX+PA-QI                  0.804 [0.545; 1.187] -1.10  0.2729
## FL                        0.196 [0.009; 4.181] -1.04  0.2967
## MF                        0.823 [0.574; 1.180] -1.06  0.2897
## OP-TX                     0.721 [0.425; 1.223] -1.21  0.2251
## OP-TX+VI-D                0.720 [0.415; 1.250] -1.17  0.2434
## PA-QI                     0.944 [0.744; 1.199] -0.47  0.6390
## PA-QI+VI-D                0.944 [0.716; 1.244] -0.41  0.6804
## SO                        1.136 [0.650; 1.985]  0.45  0.6551
```

```
## SU-EY                   0.579 [0.227; 1.473] -1.15  0.2513
## UC                          .             .     .       .
## VI-D                    0.999 [0.864; 1.155] -0.01  0.9903
##
## Incremental effect for existing combinations:
##                           iOR         95%-CI     z p-value
## BF+CL-QI                1.028 [0.659; 1.601]  0.12  0.9045
## BF+CL-QI+PA-QI          0.970 [0.608; 1.549] -0.13  0.8999
## BF+CP+EX+PA-QI          0.119 [0.014; 1.024] -1.94  0.0525
## BF+SU-PM                0.669 [0.136; 3.289] -0.49  0.6212
## CA+CL-QI+DI+PA-QI+VI-D  0.716 [0.305; 1.679] -0.77  0.4419
## CA+CL-QI+MF+VI-D        0.807 [0.600; 1.085] -1.42  0.1556
## CA+EA+PA-QI+VI-D        0.754 [0.552; 1.029] -1.78  0.0755
## CA+OP-TX+VI-D           0.599 [0.344; 1.044] -1.81  0.0706
## CA+PA-QI+VI-D           0.785 [0.605; 1.019] -1.82  0.0688
## CA+VI-D                 0.831 [0.704; 0.981] -2.19  0.0287
## CBT+CL-QI+PA-QI         0.410 [0.101; 1.668] -1.25  0.2129
## CL-QI+DE-AL+EX+MF+PA-QI 0.573 [0.156; 2.099] -0.84  0.4004
## CL-QI+DE-HP             1.009 [0.709; 1.435]  0.05  0.9617
## CL-QI+EX+MF             0.827 [0.592; 1.155] -1.12  0.2647
## CL-QI+EX+MF+PA-QI       0.781 [0.548; 1.113] -1.37  0.1719
## CL-QI+EX+PA-QI          0.949 [0.553; 1.629] -0.19  0.8493
## CL-QI+MF                0.971 [0.769; 1.226] -0.25  0.8039
## CL-QI+MF+PA-QI          0.917 [0.722; 1.165] -0.71  0.4782
## CL-QI+MF+PA-QI+SO       1.041 [0.584; 1.856]  0.14  0.8907
## CP+DE-FT+PA-QI          1.333 [0.595; 2.988]  0.70  0.4850
## DE-HP+EA+EX+PA-QI       0.661 [0.440; 0.993] -1.99  0.0464
## DE-HP+PA-QI             0.808 [0.542; 1.203] -1.05  0.2929
## DI+EX                   0.658 [0.289; 1.502] -0.99  0.3203
## EA+EX+MF                0.673 [0.434; 1.044] -1.77  0.0773
## EA+PA-QI                0.907 [0.692; 1.189] -0.70  0.4813
## EA+VA                   1.790 [0.904; 3.544]  1.67  0.0946
## EX+MF                   0.701 [0.479; 1.026] -1.83  0.0676
## EX+PA-QI                0.804 [0.545; 1.187] -1.10  0.2729
## OP-TX+VI-D              0.720 [0.415; 1.250] -1.17  0.2434
## PA-QI+VI-D              0.944 [0.716; 1.244] -0.41  0.6804
##
## Incremental effect for components:
##         iOR          95%-CI     z p-value
## BF    0.871 [0.637;  1.191] -0.87  0.3869
## CA    0.832 [0.692;  1.000] -1.96  0.0499
## CBT   0.368 [0.086;  1.581] -1.34  0.1789
## CL-QI 1.180 [0.831;  1.675]  0.92  0.3559
## CP    0.169 [0.019;  1.479] -1.61  0.1083
## DE-AL 0.734 [0.200;  2.688] -0.47  0.6400
## DE-FT 8.332 [0.835; 83.094]  1.81  0.0708
## DE-HP 0.855 [0.618;  1.183] -0.95  0.3445
## DI    0.773 [0.364;  1.640] -0.67  0.5019
## EA    0.961 [0.729;  1.266] -0.28  0.7761
## EX    0.852 [0.638;  1.136] -1.09  0.2753
## FL    0.196 [0.009;  4.181] -1.04  0.2967
## MF    0.823 [0.574;  1.180] -1.06  0.2897
## OP-TX 0.721 [0.425;  1.223] -1.21  0.2251
## PA-QI 0.944 [0.744;  1.199] -0.47  0.6390
```

```
## SO    1.136 [0.650;  1.985]  0.45  0.6551
## SU-EY 0.579 [0.227;  1.473] -1.15  0.2513
## SU-PM 0.769 [0.161;  3.660] -0.33  0.7409
## VA    1.863 [0.892;  3.892]  1.66  0.0976
## VI-D  0.999 [0.864;  1.155] -0.01  0.9903
##
## Quantifying heterogeneity / inconsistency:
## tau^2 = 0.026; tau = 0.1628; I^2 = 30% [3%; 50%]
##
## Heterogeneity statistics:
##                    Q df p-value
## Additive model 81.59 57   0.018
## Standard model 54.62 34   0.014
## Difference     26.98 23   0.257
##
## Details of network meta-analysis methods:
## - Frequentist graph-theoretical approach
## - Component network meta-analysis
## - DerSimonian-Laird estimator for tau^2
## - Calculation of I^2 based on Q
```

##### Check the additivity assumption

The difference between the Q statistics for the additive CNMA and standard NMA is 26.98 with 23 degrees of freedom resulting in a p-value well above the cut-off value, suggesting that the additivity assumption is met.

```
cnma_model_fit(cnma_frac, n = 2)
```

```
## Warning in createC.matrix(C.matrix, i, sep.comps = x$sep.comps, sep.ia = sep.ia, : Information on
## interaction 'BF * SU-PM' is already available in column 'SU-PM'.

## Warning in createC.matrix(C.matrix, i, sep.comps = x$sep.comps, sep.ia = sep.ia, : Information on
## interaction 'EA * VA' is already available in column 'VA'.

## Warning in max(str_count(cn.i.ia, fixed(cnma$sep.ia))): no non-missing arguments to max; returning
## -Inf
## Warning in max(str_count(cn.i.ia, fixed(cnma$sep.ia))): no non-missing arguments to max; returning
## -Inf

##    interaction     Q df  pval  tau2  I2 Q.nma df.nma pval.nma Q.add df.add pval.add
##     BF * CL-QI 80.84 56 0.017 0.027 31% 26.22     22    0.242  0.76      1    0.385
##      CA * VI-D 80.76 56 0.017 0.029 31% 26.15     22    0.245  0.83      1    0.362
##  CL-QI * DE-HP 81.27 56 0.015 0.028 31% 26.65     22    0.225  0.32      1    0.569
##     CL-QI * MF 81.50 56 0.015 0.028 31% 26.89     22    0.216  0.09      1    0.763
##  DE-HP * PA-QI 81.27 56 0.015 0.028 31% 26.65     22    0.225  0.32      1    0.569
##        DI * EX 81.42 56 0.015 0.027 31% 26.81     22    0.219  0.17      1    0.680
##     EA * PA-QI 81.23 56 0.015 0.027 31% 26.62     22    0.226  0.36      1    0.549
##        EX * MF 80.74 56 0.017 0.027 31% 26.13     22    0.246  0.85      1    0.356
##     EX * PA-QI 80.10 56 0.019 0.026 30% 25.49     22    0.274  1.49      1    0.222
##   OP-TX * VI-D 72.53 56 0.068 0.018 23% 17.91     22    0.711  9.06      1    0.003
##   PA-QI * VI-D 79.74 56 0.020 0.028 30% 25.12     22    0.291  1.86      1    0.173
```

##### Rank treatments

Rank treatments by P-scores:

```r
netrank(cnma_frac)
```

```
##                           P-score
## BF+CP+EX+PA-QI             0.9436
## FL                         0.8058
## CBT+CL-QI+PA-QI            0.7926
## CA+OP-TX+VI-D              0.7784
## DE-HP+EA+EX+PA-QI          0.7371
## SU-EY                      0.7159
## EA+EX+MF                   0.7072
## EX+MF                      0.6842
## CL-QI+DE-AL+EX+MF+PA-QI    0.6828
## DI+EX                      0.6672
## CA+EA+PA-QI+VI-D           0.6298
## OP-TX                      0.6243
## OP-TX+VI-D                 0.6227
## CA+CL-QI+DI+PA-QI+VI-D     0.6033
## CA+PA-QI+VI-D              0.5950
## CL-QI+EX+MF+PA-QI          0.5929
## BF+SU-PM                   0.5823
## CA+CL-QI+MF+VI-D           0.5517
## EX+PA-QI                   0.5499
## DE-AL                      0.5456
## DI                         0.5428
## DE-HP+PA-QI                0.5387
## CA+VI-D                    0.5176
## CL-QI+EX+MF                0.5153
## CA                         0.5147
## MF                         0.5104
## EX                         0.4757
## BF                         0.4573
## EA+PA-QI                   0.3993
## CL-QI+MF+PA-QI             0.3862
## CL-QI+EX+PA-QI             0.3699
## PA-QI+VI-D                 0.3489
## BF+CL-QI+PA-QI             0.3474
## PA-QI                      0.3432
## CL-QI+MF                   0.3117
## CL-QI+MF+PA-QI+SO          0.2986
## CL-QI+DE-HP                0.2942
## BF+CL-QI                   0.2860
## VI-D                       0.2680
## UC                         0.2590
## SO                         0.2245
## CP+DE-FT+PA-QI             0.1778
## CL-QI                      0.1484
## EA+VA                      0.0521
```

##### Additive CNMA forest plots

First, generate a forest plot of all treatment-level effects:

```r
xlim_vals <-
  range(cnma_frac$lower.cnma.random, cnma_frac$upper.cnma.random, na.rm = TRUE)
#
```

```r
xlim_vals <- round(exp(xlim_vals), 2)
```

```r
forest(cnma_frac, sortvar = -TE,
  smlab = TitleCNMA,
  label.left = LeftLabelInt, label.right = RightLabelInt,
  leftcols = c("studlab", "k"),
  leftlabs =
    c(paste("Contrast to", cnma_frac$reference.group), "Direct\nComparisons"),
  xlim = xlim_vals)
```

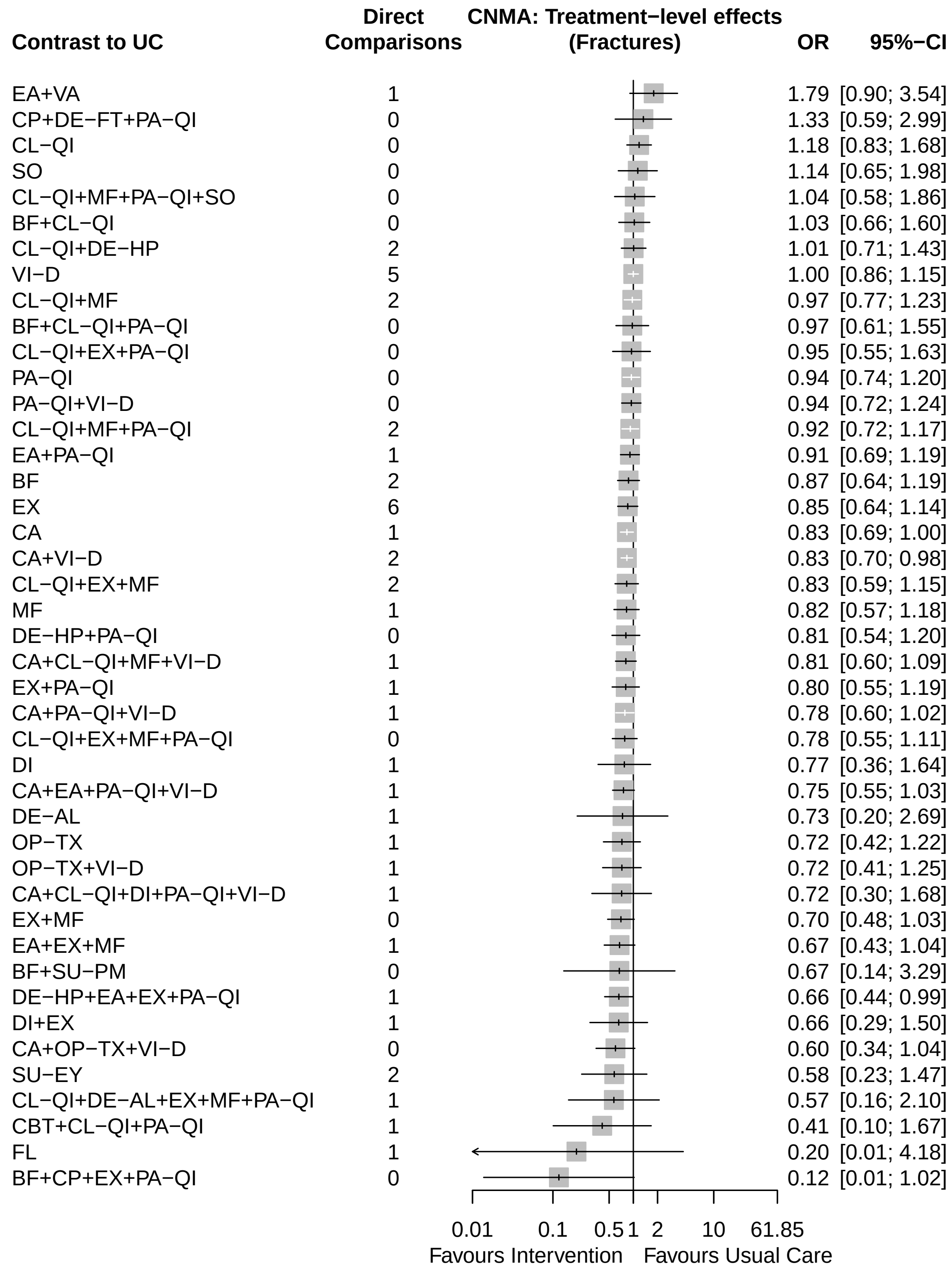


Next, generate a forest plot of all component-level effects:

```
# Component additive CNMA forest plot
forest(netcomplex(cnma_frac, cnma_frac$comps),
  smlab = TitleCompCNMA,
  label.left = LeftLabelComp, label.right = RightLabelComp,
  xlim = xlim_vals)
```

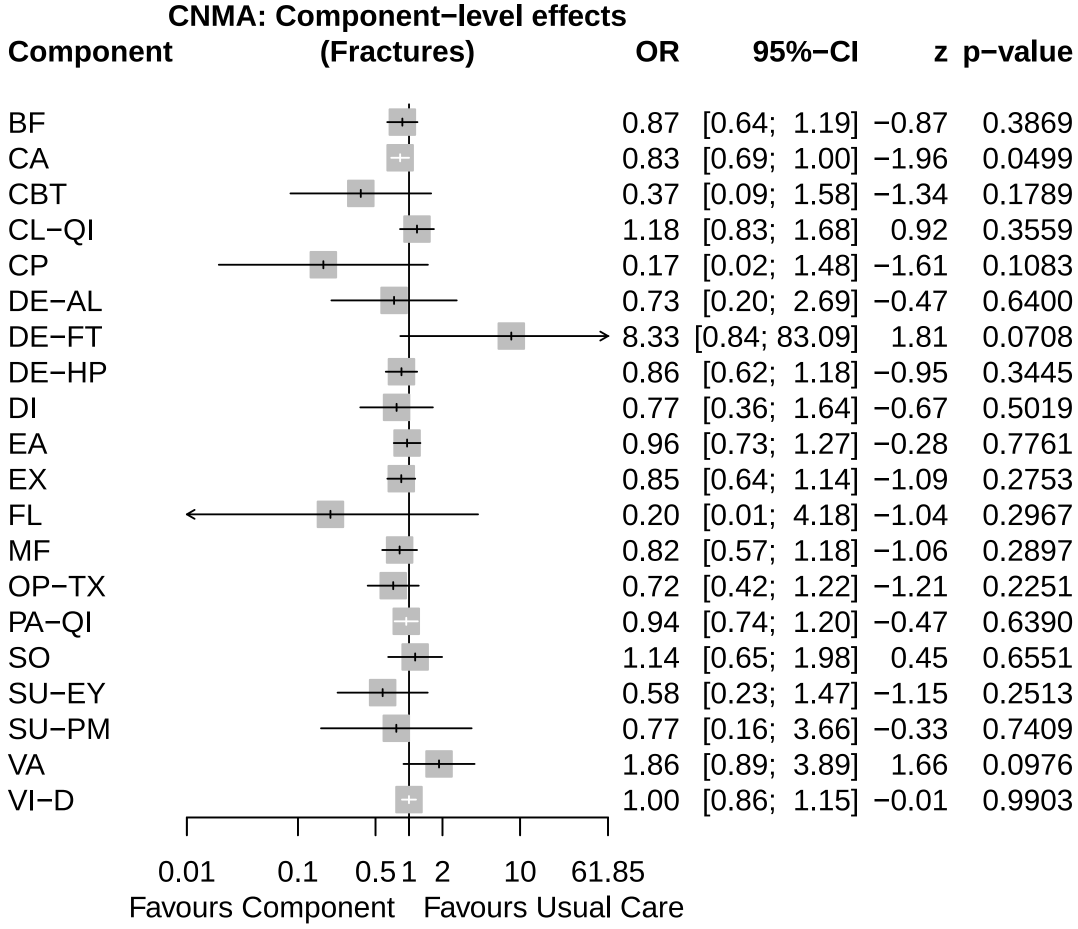


Finally, generate a forest plot comparing the results of the standard NMA and CNMA models. The font size, spacing, and square size can be adjusted to fit the figure to the page.

```
nb_frac <- netbind(cnma_frac, nma_frac,
  name = c("Additive CNMA", "Standard NMA"),
  col.study = c("red", "black"), col.square = c("red", "black"))
#
forest(nb_frac,
  smlab = Title,
  label.left = LeftLabelInt, label.right = RightLabelInt,
  fontsize = 6, spacing = 0.5, square.size = 0.4,
  addrow.subgroups = FALSE)
```

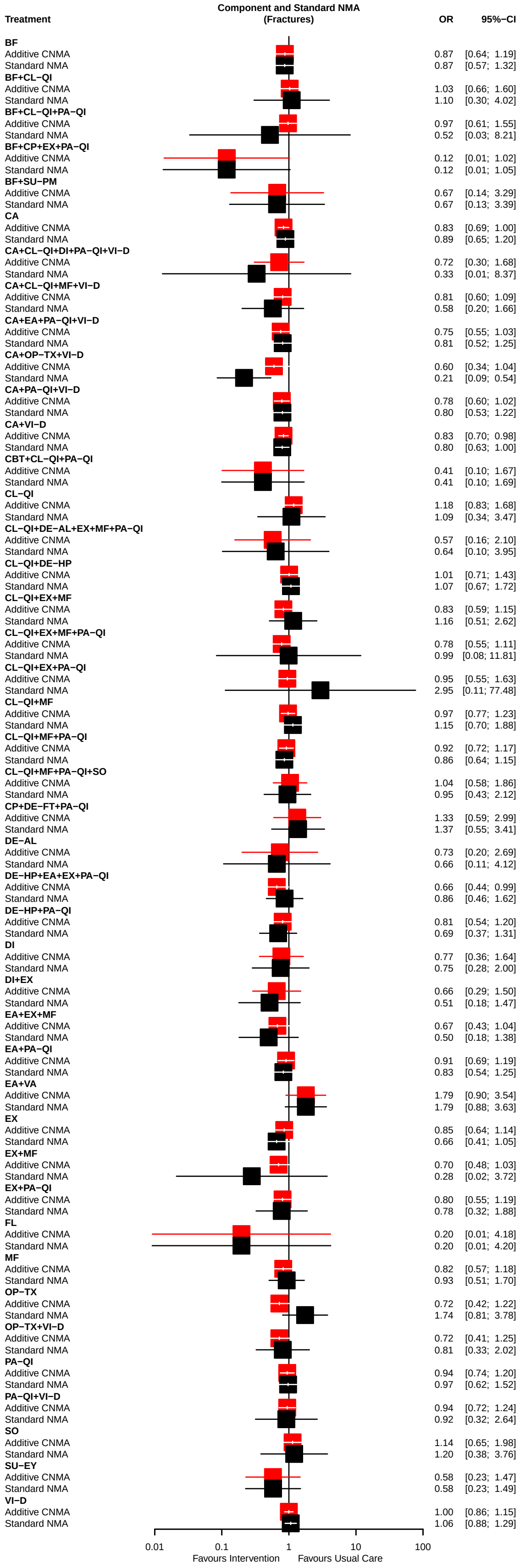
Component and Standard NMA
(Fractures)
Treatment
OR
95%−CI
BF
Additive CNMA 0.87 [0.64; 1.19]
Standard NMA 0.87 [0.57; 1.32]
BF+CL−QI
Additive CNMA 1.03 [0.66; 1.60]
Standard NMA 1.10 [0.30; 4.02]
BF+CL−QI+PA−QI
Additive CNMA 0.97 [0.61; 1.55]
Standard NMA 0.52 [0.03; 8.21]
BF+CP+EX+PA−QI
Additive CNMA 0.12 [0.01; 1.02]
Standard NMA 0.12 [0.01; 1.05]
BF+SU−PM
Additive CNMA 0.67 [0.14; 3.29]
Standard NMA 0.67 [0.13; 3.39]
CA
Additive CNMA 0.83 [0.69; 1.00]
Standard NMA 0.89 [0.65; 1.20]
CA+CL−QI+DI+PA−QI+VI−D
Additive CNMA 0.72 [0.30; 1.68]
Standard NMA 0.33 [0.01; 8.37]
CA+CL−QI+MF+VI−D
Additive CNMA 0.81 [0.60; 1.09]
Standard NMA 0.58 [0.20; 1.66]
CA+EA+PA−QI+VI−D
Additive CNMA 0.75 [0.55; 1.03]
Standard NMA 0.81 [0.52; 1.25]
CA+OP−TX+VI−D
Additive CNMA 0.60 [0.34; 1.04]
Standard NMA 0.21 [0.09; 0.54]
CA+PA−QI+VI−D
Additive CNMA 0.78 [0.60; 1.02]
Standard NMA 0.80 [0.53; 1.22]
CA+VI−D
Additive CNMA 0.83 [0.70; 0.98]
Standard NMA 0.80 [0.63; 1.00]
CBT+CL−QI+PA−QI
Additive CNMA 0.41 [0.10; 1.67]
Standard NMA 0.41 [0.10; 1.69]
CL−QI
Additive CNMA 1.18 [0.83; 1.68]
Standard NMA 1.09 [0.34; 3.47]
CL−QI+DE−AL+EX+MF+PA−QI
Additive CNMA 0.57 [0.16; 2.10]
Standard NMA 0.64 [0.10; 3.95]
CL−QI+DE−HP
Additive CNMA 1.01 [0.71; 1.43]
Standard NMA 1.07 [0.67; 1.72]
CL−QI+EX+MF
Additive CNMA 0.83 [0.59; 1.15]
Standard NMA 1.16 [0.51; 2.62]
CL−QI+EX+MF+PA−QI
Additive CNMA 0.78 [0.55; 1.11]
Standard NMA 0.99 [0.08; 11.81]
CL−QI+EX+PA−QI
Additive CNMA 0.95 [0.55; 1.63]
Standard NMA 2.95 [0.11; 77.48]
CL−QI+MF
Additive CNMA 0.97 [0.77; 1.23]
Standard NMA 1.15 [0.70; 1.88]
CL−QI+MF+PA−QI
Additive CNMA 0.92 [0.72; 1.17]
Standard NMA 0.86 [0.64; 1.15]
CL−QI+MF+PA−QI+SO
Additive CNMA 1.04 [0.58; 1.86]
Standard NMA 0.95 [0.43; 2.12]
CP+DE−FT+PA−QI
Additive CNMA 1.33 [0.59; 2.99]
Standard NMA 1.37 [0.55; 3.41]
DE−AL
Additive CNMA 0.73 [0.20; 2.69]
Standard NMA 0.66 [0.11; 4.12]
DE−HP+EA+EX+PA−QI
Additive CNMA 0.66 [0.44; 0.99]
Standard NMA 0.86 [0.46; 1.62]
DE−HP+PA−QI
Additive CNMA 0.81 [0.54; 1.20]
Standard NMA 0.69 [0.37; 1.31]
DI
Additive CNMA 0.77 [0.36; 1.64]
Standard NMA 0.75 [0.28; 2.00]
DI+EX
Additive CNMA 0.66 [0.29; 1.50]
Standard NMA 0.51 [0.18; 1.47]
EA+EX+MF
Additive CNMA 0.67 [0.43; 1.04]
Standard NMA 0.50 [0.18; 1.38]
EA+PA−QI
Additive CNMA 0.91 [0.69; 1.19]
Standard NMA 0.83 [0.54; 1.25]
EA+VA
Additive CNMA 1.79 [0.90; 3.54]
Standard NMA 1.79 [0.88; 3.63]
EX
Additive CNMA 0.85 [0.64; 1.14]
Standard NMA 0.66 [0.41; 1.05]
EX+MF
Additive CNMA 0.70 [0.48; 1.03]
Standard NMA 0.28 [0.02; 3.72]
EX+PA−QI
Additive CNMA 0.80 [0.55; 1.19]
Standard NMA 0.78 [0.32; 1.88]
FL
Additive CNMA 0.20 [0.01; 4.18]
Standard NMA 0.20 [0.01; 4.20]
MF
Additive CNMA 0.82 [0.57; 1.18]
Standard NMA 0.93 [0.51; 1.70]
OP−TX
Additive CNMA 0.72 [0.42; 1.22]
Standard NMA 1.74 [0.81; 3.78]
OP−TX+VI−D
Additive CNMA 0.72 [0.41; 1.25]
Standard NMA 0.81 [0.33; 2.02]
PA−QI
Additive CNMA 0.94 [0.74; 1.20]
Standard NMA 0.97 [0.62; 1.52]
PA−QI+VI−D
Additive CNMA 0.94 [0.72; 1.24]
Standard NMA 0.92 [0.32; 2.64]
SO
Additive CNMA 1.14 [0.65; 1.98]
Standard NMA 1.20 [0.38; 3.76]
SU−EY
Additive CNMA 0.58 [0.23; 1.47]
Standard NMA 0.58 [0.23; 1.49]
VI−D
Additive CNMA 1.00 [0.86; 1.15]
Standard NMA 1.06 [0.88; 1.29]
0.01
0.1
1
10
100
Favours Intervention
Favours Usual Care

### 3. Continuous outcome example: Quality of Life

In the following, we will look in detail at the results of the QoL data set with the continuous outcome.

The settings for the continuous outcome are quite different.

```
Sheet <- "QoL"
Name <- "Quality of Life"
#
Title <- paste0("Component and Standard NMA\n(", Name, ")")
TitleNMA <- paste0("Standard NMA\n(", Name, ")")
TitleCNMA <- paste0("CNMA: Treatment-level effects\n(", Name, ")")
TitleCompCNMA <- paste0("CNMA: Component-level effects\n(", Name, ")")
#
EffectSize <- "SMD"
SmallVal <- "undesirable"
#
Reference <- "UC"
#
left.int <- "Usual Care"
left.comp <- "Usual Care"
right.int <- "Intervention"
right.comp <- "Component"
#
LeftLabelInt <- paste("Favours", left.int)
LeftLabelComp <- paste("Favours", left.comp)
RightLabelInt <- paste("Favours", right.int)
RightLabelComp <- paste("Favours", right.comp)
```

Note, in this example, the sheet and outcome name should be different.

#### 3.1 Transform data set to comparison-based format

```
dat_qol <- read_xlsx("CNMA_data.xlsx", sheet = Sheet)
#
pw_qol <- pairwise(list(t1, t2, t3),
  n = list(n1, n2, n3), mean = list(m1, m2, m3), sd = list(sd1, sd2, sd3),
  data = dat_qol, studlab = study,
  sm = EffectSize,
  reference.group = Reference,
  append = FALSE)
```

Check the connectivity of the network.

```
netconnection(pw_qol)
```

```
## Number of studies: k = 35
## Number of pairwise comparisons: m = 39
## Number of treatments: n = 29
## Number of designs: d = 26
## Number of networks: 1
```

#### 3.2 Fit standard NMA

Finally, we conduct the standard NMA as the network is connected.

```r
nma_qol <- netmeta(pw_qol, common = FALSE, small.values = SmallVal)
nma_qol
```

```
## Number of studies: k = 35
## Number of pairwise comparisons: m = 39
## Number of observations: o = 11789
## Number of treatments: n = 29
## Number of designs: d = 26
##
## Random effects model
##
## Treatment estimate (sm = 'SMD', comparison: other treatments vs 'UC'):
##                                  SMD            95%-CI     z  p-value
## C-M+PX_ED                      0.205 [-0.151;  0.561]  1.13   0.2595
## C-M+S-M                        0.216 [ 0.089;  0.343]  3.33   0.0009
## C-M+S-M+PX_ED                  0.061 [-0.131;  0.253]  0.62   0.5323
## EPR+RE+C-M+PX_RE+PX_ED         0.033 [-0.217;  0.283]  0.26   0.7973
## EPR+STA                        0.014 [-0.263;  0.291]  0.10   0.9214
## EPR+STA+CLIN_RE+PX_RE         -0.026 [-0.211;  0.160] -0.27   0.7844
## PX_ED                         -0.780 [-1.416; -0.144] -2.40   0.0163
## RE+S-M+PX_ED                   0.006 [-0.153;  0.165]  0.08   0.9393
## RE+S-M+PX_RE+PX_ED             1.843 [ 1.159;  2.527]  5.28 < 0.0001
## S-M                            0.807 [ 0.477;  1.136]  4.80 < 0.0001
## S-M+PX_ED                      0.105 [-0.230;  0.440]  0.62   0.5383
## S-M+PX_RE                     -0.059 [-0.309;  0.191] -0.46   0.6439
## STA                           -0.043 [-0.415;  0.329] -0.23   0.8215
## STA+C-M+S-M                   -0.061 [-0.574;  0.452] -0.23   0.8161
## STA+C-M+S-M+PX_ED              0.040 [-0.172;  0.253]  0.37   0.7114
## STA+C-M+S-M+PX_ED+MOT_INT      0.049 [-0.280;  0.378]  0.29   0.7723
## STA+S-M                        0.057 [-0.090;  0.205]  0.76   0.4453
## STA+S-M+PX_ED                 -0.067 [-0.383;  0.250] -0.41   0.6804
## TE+C-M+PX_ED                  -0.163 [-0.546;  0.221] -0.83   0.4057
## TE+C-M+S-M+PX_ED               0.036 [-0.155;  0.227]  0.37   0.7119
## TE+C-M+S-M+PX_ED+MOT_INT      -0.727 [-1.504;  0.050] -1.83   0.0667
## TE+C-M+S-M+PX_RE+PX_ED        -0.454 [-1.229;  0.321] -1.15   0.2511
## TE+EPR+RE+AF+C-M+S-M+PX_ED     0.035 [-0.264;  0.334]  0.23   0.8192
## TE+EPR+RE+C-M+S-M             -0.086 [-0.293;  0.122] -0.81   0.4192
## TE+PX_ED                       0.000 [-0.159;  0.159]  0.00   1.0000
## TE+STA+C-M+S-M+PX_ED          -0.099 [-0.237;  0.039] -1.40   0.1611
## TE+STA+C-M+S-M+PX_ED+PX_FIN    0.023 [-0.424;  0.469]  0.10   0.9212
## TE+STA+FIN+C-M+S-M+PX_ED       0.096 [-0.052;  0.244]  1.28   0.2020
## UC                                 .                 .     .        .
##
## Quantifying heterogeneity / inconsistency:
## tau^2 = 0.003; tau = 0.0545; I^2 = 19% [0%; 60%]
##
## Tests of heterogeneity (within designs) and inconsistency (between designs):
##                     Q d.f. p-value
## Total           11.11    9   0.268
## Within designs  11.11    9   0.268
## Between designs  0.00    0      --
##
## Details of network meta-analysis methods:
## - Frequentist graph-theoretical approach
```

```
## - DerSimonian-Laird estimator for tau^2
## - Calculation of I^2 based on Q
```

##### Network diagram

```r
par(mar = c(5.1, 4.1, 4.1, 10.1))
netgraph(nma_qol, points = TRUE,
  multiarm = TRUE, col.multiarm = "blue", thickness = "equal", seq = "optimal")
```

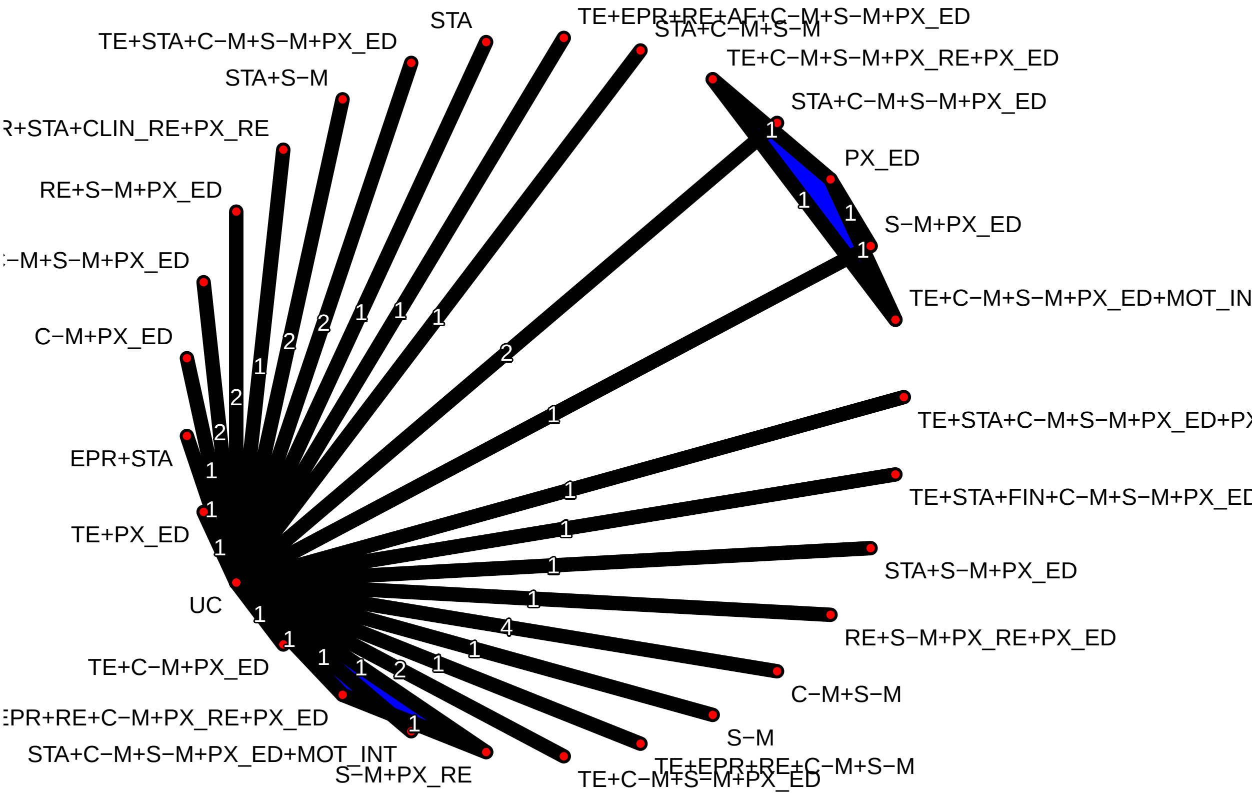


##### Rank treatments

Rank treatments by P-scores:

```r
netrank(nma_qol)
```

```
##                              P-score
## RE+S-M+PX_RE+PX_ED            0.9999
## S-M                           0.9637
## C-M+S-M                       0.8442
## C-M+PX_ED                     0.7593
## TE+STA+FIN+C-M+S-M+PX_ED      0.6737
## S-M+PX_ED                     0.6443
## STA+S-M                       0.5990
## C-M+S-M+PX_ED                 0.5990
## STA+C-M+S-M+PX_ED+MOT_INT     0.5601
## STA+C-M+S-M+PX_ED             0.5566
## TE+C-M+S-M+PX_ED              0.5506
## EPR+RE+C-M+PX_RE+PX_ED        0.5429
## TE+EPR+RE+AF+C-M+S-M+PX_ED    0.5410
```

```
## TE+STA+C-M+S-M+PX_ED+PX_FIN  0.5178
## EPR+STA                      0.5071
## RE+S-M+PX_ED                 0.4912
## TE+PX_ED                     0.4782
## UC                           0.4733
## EPR+STA+CLIN_RE+PX_RE        0.4285
## STA                          0.4266
## STA+C-M+S-M                  0.4209
## STA+S-M+PX_ED                0.3836
## S-M+PX_RE                    0.3788
## TE+EPR+RE+C-M+S-M            0.3269
## TE+STA+C-M+S-M+PX_ED         0.2842
## TE+C-M+PX_ED                 0.2782
## TE+C-M+S-M+PX_RE+PX_ED       0.1800
## TE+C-M+S-M+PX_ED+MOT_INT     0.0605
## PX_ED                        0.0298
```

##### Standard NMA forest plot

Generate a forest plot of the standard NMA effect estimates sorted by P-scores:

```
xlim_vals <-
  range(nma_qol$lower.nma.random, nma_qol$upper.nma.random, na.rm = TRUE)
```

```
forest(nma_qol, sortvar = -Pscore,
  smlab = TitleNMA,
  label.left = LeftLabelInt, label.right = RightLabelInt,
  leftcols = c("studlab", "k"),
  leftlabs =
    c(paste("Contrast to", nma_qol$reference.group), "Direct\nComparisons"),
  xlim = xlim_vals)
```

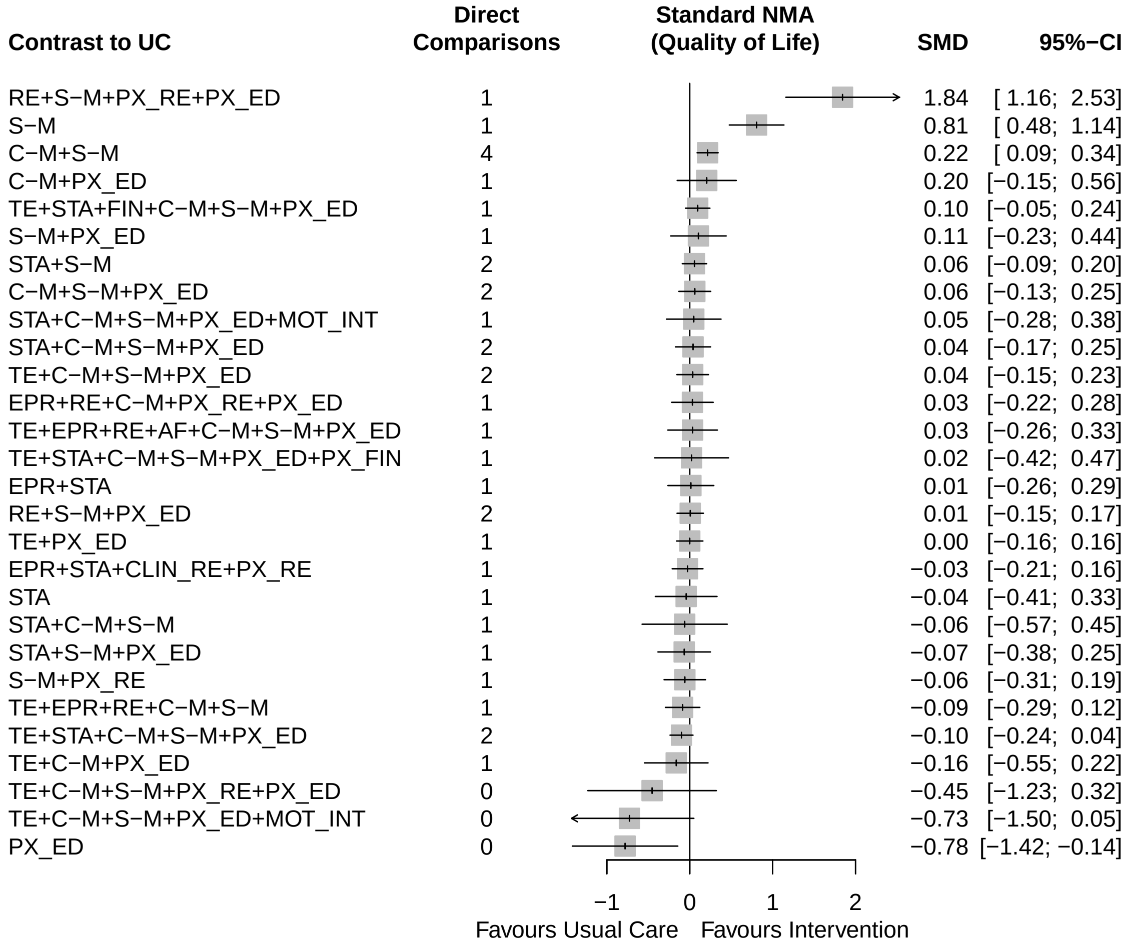


##### 3.3 Fit the additive CNMA model

The reference treatment from the standard NMA is defined as an inactive component in the additive CNMA model.

```r
cnma_qol <- netcomb(nma_qol, inactive = nma_qol$reference.group)
cnma_qol
```

```
## Number of studies: k = 35
## Number of pairwise comparisons: m = 39
## Number of treatments: n = 29
## Number of active components: c = 13
## Number of designs: d = 26
##
## Random effects model (inactive component: 'UC')
##
## Treatment estimate (sm = 'SMD', comparison: other treatments vs 'UC'):
##                                SMD           95%-CI     z  p-value
## C-M+PX_ED                   -0.015 [-0.213; 0.183] -0.15  0.8793
## C-M+S-M                      0.164 [-0.004; 0.332]  1.91  0.0556
## C-M+S-M+PX_ED                0.178 [-0.013; 0.368]  1.83  0.0677
## EPR+RE+C-M+PX_RE+PX_ED       0.249 [-0.115; 0.612]  1.34  0.1798
```

```
## EPR+STA                      -0.086 [-0.430; 0.258] -0.49  0.6248
## EPR+STA+CLIN_RE+PX_RE        -0.026 [-0.428; 0.376] -0.13  0.8995
## PX_ED                         0.014 [-0.166; 0.193]  0.15  0.8820
## RE+S-M+PX_ED                  0.169 [-0.093; 0.431]  1.27  0.2053
## RE+S-M+PX_RE+PX_ED            0.421 [ 0.060; 0.783]  2.29  0.0222
## S-M                           0.193 [ 0.031; 0.355]  2.33  0.0196
## S-M+PX_ED                     0.207 [-0.047; 0.460]  1.60  0.1099
## S-M+PX_RE                     0.446 [ 0.122; 0.769]  2.70  0.0069
## STA                          -0.135 [-0.330; 0.060] -1.36  0.1747
## STA+C-M+S-M                   0.029 [-0.198; 0.257]  0.25  0.8014
## STA+C-M+S-M+PX_ED             0.043 [-0.141; 0.227]  0.46  0.6491
## STA+C-M+S-M+PX_ED+MOT_INT     0.046 [-0.325; 0.418]  0.24  0.8073
## STA+S-M                       0.058 [-0.119; 0.236]  0.64  0.5212
## STA+S-M+PX_ED                 0.072 [-0.137; 0.280]  0.67  0.5006
## TE+C-M+PX_ED                 -0.150 [-0.360; 0.061] -1.39  0.1630
## TE+C-M+S-M+PX_ED              0.044 [-0.141; 0.228]  0.46  0.6443
## TE+C-M+S-M+PX_ED+MOT_INT      0.047 [-0.348; 0.442]  0.23  0.8155
## TE+C-M+S-M+PX_RE+PX_ED        0.296 [-0.041; 0.633]  1.72  0.0854
## TE+EPR+RE+AF+C-M+S-M+PX_ED    0.035 [-0.431; 0.500]  0.15  0.8832
## TE+EPR+RE+C-M+S-M             0.041 [-0.292; 0.374]  0.24  0.8074
## TE+PX_ED                     -0.121 [-0.346; 0.105] -1.05  0.2945
## TE+STA+C-M+S-M+PX_ED         -0.091 [-0.293; 0.110] -0.89  0.3732
## TE+STA+C-M+S-M+PX_ED+PX_FIN   0.023 [-0.549; 0.594]  0.08  0.9384
## TE+STA+FIN+C-M+S-M+PX_ED      0.096 [-0.290; 0.482]  0.49  0.6257
## UC                                .                .     .       .
##
## Incremental effect for existing combinations:
##                                 iSMD           95%-CI      z p-value
## C-M+PX_ED                    -0.015 [-0.213; 0.183] -0.15  0.8793
## C-M+S-M                       0.164 [-0.004; 0.332]  1.91  0.0556
## C-M+S-M+PX_ED                 0.178 [-0.013; 0.368]  1.83  0.0677
## EPR+RE+C-M+PX_RE+PX_ED        0.249 [-0.115; 0.612]  1.34  0.1798
## EPR+STA                      -0.086 [-0.430; 0.258] -0.49  0.6248
## EPR+STA+CLIN_RE+PX_RE        -0.026 [-0.428; 0.376] -0.13  0.8995
## RE+S-M+PX_ED                  0.169 [-0.093; 0.431]  1.27  0.2053
## RE+S-M+PX_RE+PX_ED            0.421 [ 0.060; 0.783]  2.29  0.0222
## S-M+PX_ED                     0.207 [-0.047; 0.460]  1.60  0.1099
## S-M+PX_RE                     0.446 [ 0.122; 0.769]  2.70  0.0069
## STA+C-M+S-M                   0.029 [-0.198; 0.257]  0.25  0.8014
## STA+C-M+S-M+PX_ED             0.043 [-0.141; 0.227]  0.46  0.6491
## STA+C-M+S-M+PX_ED+MOT_INT     0.046 [-0.325; 0.418]  0.24  0.8073
## STA+S-M                       0.058 [-0.119; 0.236]  0.64  0.5212
## STA+S-M+PX_ED                 0.072 [-0.137; 0.280]  0.67  0.5006
## TE+C-M+PX_ED                 -0.150 [-0.360; 0.061] -1.39  0.1630
## TE+C-M+S-M+PX_ED              0.044 [-0.141; 0.228]  0.46  0.6443
## TE+C-M+S-M+PX_ED+MOT_INT      0.047 [-0.348; 0.442]  0.23  0.8155
## TE+C-M+S-M+PX_RE+PX_ED        0.296 [-0.041; 0.633]  1.72  0.0854
## TE+EPR+RE+AF+C-M+S-M+PX_ED    0.035 [-0.431; 0.500]  0.15  0.8832
## TE+EPR+RE+C-M+S-M             0.041 [-0.292; 0.374]  0.24  0.8074
## TE+PX_ED                     -0.121 [-0.346; 0.105] -1.05  0.2945
## TE+STA+C-M+S-M+PX_ED         -0.091 [-0.293; 0.110] -0.89  0.3732
## TE+STA+C-M+S-M+PX_ED+PX_FIN   0.023 [-0.549; 0.594]  0.08  0.9384
## TE+STA+FIN+C-M+S-M+PX_ED      0.096 [-0.290; 0.482]  0.49  0.6257
##
```

```
## Incremental effect for components:
##           iSMD          95%-CI     z p-value
## AF      -0.020 [-0.576; 0.536] -0.07  0.9435
## C-M     -0.029 [-0.221; 0.163] -0.30  0.7677
## CLIN_RE -0.193 [-0.789; 0.404] -0.63  0.5272
## EPR      0.049 [-0.336; 0.435]  0.25  0.8027
## FIN      0.188 [-0.248; 0.623]  0.84  0.3985
## MOT_INT  0.003 [-0.366; 0.373]  0.02  0.9855
## PX_ED    0.014 [-0.166; 0.193]  0.15  0.8820
## PX_FIN   0.114 [-0.492; 0.720]  0.37  0.7123
## PX_RE    0.252 [-0.053; 0.558]  1.62  0.1050
## RE      -0.038 [-0.367; 0.291] -0.22  0.8223
## S-M      0.193 [ 0.031; 0.355]  2.33  0.0196
## STA     -0.135 [-0.330; 0.060] -1.36  0.1747
## TE      -0.134 [-0.331; 0.063] -1.34  0.1812
##
## Quantifying heterogeneity / inconsistency:
## tau^2 = 0.036; tau = 0.1900; I^2 = 69% [54%; 80%]
##
## Heterogeneity statistics:
##                    Q df p-value
## Additive model 77.82 24 < 0.001
## Standard model 11.11  9   0.268
## Difference     66.71 15 < 0.001
##
## Details of network meta-analysis methods:
## - Frequentist graph-theoretical approach
## - Component network meta-analysis
## - DerSimonian-Laird estimator for tau^2
## - Calculation of I^2 based on Q
```

**Check the additivity assumption**

The difference between the Q statistics for the additive CNMA and standard NMA is 66.71 with 15 degrees of freedom resulting in a p-value well below the cut-off value, suggesting that the additivity assumption does not hold.

#### 3.4 Model selection of interaction CNMA models

We fit interaction CNMA models with a single 2-way interaction and calculate the difference in Q statistics between each of the interaction models and the additive model.

```r
cnma_model_fit(cnma_qol, n = 2)
```

```
##  interaction     Q df    pval  tau2  I2 Q.nma df.nma pval.nma Q.add df.add pval.add
##  C-M * PX_ED 76.73 23 < 0.001 0.039 70% 65.62     14  < 0.001  1.09      1    0.297
##    C-M * S-M 75.45 23 < 0.001 0.037 70% 64.34     14  < 0.001  2.37      1    0.124
##    EPR * STA 77.57 23 < 0.001 0.038 70% 66.46     14  < 0.001  0.25      1    0.614
##  S-M * PX_ED 75.96 23 < 0.001 0.039 70% 64.85     14  < 0.001  1.86      1    0.173
##  S-M * PX_RE 75.39 23 < 0.001 0.037 70% 64.28     14  < 0.001  2.43      1    0.119
##    STA * S-M 77.28 23 < 0.001 0.038 70% 66.17     14  < 0.001  0.54      1    0.464
##   TE * PX_ED 73.90 23 < 0.001 0.036 69% 62.79     14  < 0.001  3.92      1    0.048
```

Repeat the above for all available 3-way interactions in the data set.

```r
cnma_model_fit(cnma_qol, n = 3)
```

```
##        interaction     Q df    pval  tau2  I2 Q.nma df.nma pval.nma Q.add df.add pval.add
##  C-M * S-M * PX_ED 74.72 23 < 0.001 0.037 69% 63.61     14  < 0.001  3.10      1    0.078
##   RE * S-M * PX_ED 77.57 23 < 0.001 0.038 70% 66.46     14  < 0.001  0.25      1    0.614
##    STA * C-M * S-M 77.11 23 < 0.001 0.039 70% 66.00     14  < 0.001  0.71      1    0.399
##  STA * S-M * PX_ED 76.43 23 < 0.001 0.038 70% 65.32     14  < 0.001  1.39      1    0.239
##   TE * C-M * PX_ED 76.98 23 < 0.001 0.040 70% 65.87     14  < 0.001  0.84      1    0.360
```

Theoretically, you could repeat this process for all interactions up to 7-way interactions in the QOL data set (because there are treatments containing up to 7 components), but for illustration purposes, we will stop with the 3-way interaction models.

#### 3.5 Build the interaction CNMA models

First, we will build the single-interaction models for available 2-way and 3-way interactions:

```r
combs2_qol <- combinations(cnma_qol, n = 2)
combs2_qol
```

```
## [1] "C-M+PX_ED" "C-M+S-M"   "EPR+STA"   "S-M+PX_ED" "S-M+PX_RE" "STA+S-M"   "TE+PX_ED"
```

```r
#
combs3_qol <- combinations(cnma_qol, n = 3)
combs3_qol
```

```
## [1] "C-M+S-M+PX_ED" "RE+S-M+PX_ED"  "STA+C-M+S-M"   "STA+S-M+PX_ED" "TE+C-M+PX_ED"
```

```r
#
# Interaction model 1: C-M * PX_ED
C1_qol <- createC(cnma_qol, combs2_qol[1])
colnames_ia(C1_qol)
```

```
## [1] "C-M * PX_ED"
```

```r
# Interaction model 2: C-M * S-M
C2_qol <- createC(cnma_qol, combs2_qol[2])
colnames_ia(C2_qol)
```

```
## [1] "C-M * S-M"
```

```r
# Interaction model 3: EPR * STA
C3_qol <- createC(cnma_qol, combs2_qol[3])
colnames_ia(C3_qol)
```

```
## [1] "EPR * STA"
```

```r
# Interaction model 4: S-M * PX_ED
C4_qol <- createC(cnma_qol, combs2_qol[4])
colnames_ia(C4_qol)
```

```
## [1] "S-M * PX_ED"
```

```r
# Interaction model 5: S-M * PX_RE
C5_qol <- createC(cnma_qol, combs2_qol[5])
colnames_ia(C5_qol)
```

```
## [1] "S-M * PX_RE"
```

```r
# Interaction model 6: STA * S-M
C6_qol <- createC(cnma_qol, combs2_qol[6])
colnames_ia(C6_qol)
```

```
## [1] "STA * S-M"
```

```r
# Interaction model 7: TE * PX_ED
C7_qol <- createC(cnma_qol, combs2_qol[7])
colnames_ia(C7_qol)
```

```
## [1] "TE * PX_ED"
```

```r
# Interaction model 8:  C-M * S-M * PX_ED + C-M * PX_ED + C-M * S-M + S-M * PX_ED
C8_qol <- createC(cnma_qol, c(combs3_qol[1], combs2_qol[c(1, 2, 4)]))
colnames_ia(C8_qol)
```

```
## [1] "C-M * S-M * PX_ED" "C-M * PX_ED"       "C-M * S-M"         "S-M * PX_ED"
```

```r
# Interaction model 9: RE * S-M * PX_ED + S-M * PX_ED
C9_qol <- createC(cnma_qol, c(combs3_qol[2], combs2_qol[4]))
colnames_ia(C9_qol)
```

```
## [1] "RE * S-M * PX_ED" "S-M * PX_ED"
```

```r
# Interaction model 10: STA * C-M * S-M + C-M * S-M + STA * S-M
C10_qol <- createC(cnma_qol, c(combs3_qol[3], combs2_qol[c(2, 6)]))
colnames_ia(C10_qol)
```

```
## [1] "STA * C-M * S-M" "C-M * S-M"       "STA * S-M"
```

```r
# Interaction model 11: STA * S-M * PX_ED + S-M * PX_ED + STA * S-M
C11_qol <- createC(cnma_qol, c(combs3_qol[4], combs2_qol[c(4, 6)]))
colnames_ia(C11_qol)
```

```
## [1] "STA * S-M * PX_ED" "S-M * PX_ED"       "STA * S-M"
```

```r
# Interaction model 12: TE * C-M * PX_ED + C-M * PX_ED
C12_qol <- createC(cnma_qol, c(combs3_qol[5], combs2_qol[1]))
colnames_ia(C12_qol)
```

```
## [1] "TE * C-M * PX_ED" "C-M * PX_ED"
```

Next, we will manually create C matrices for two-interaction models built upon the C matrices of the 2-way interaction models above:

```r
# Interaction model 13: c-M * s-M (C2_qol) + s-M * px_re
C13_qol <- createC(C2_qol, "s-M + px_re")

# Interaction model 14: c-M * s-M (C2_qol) + te * px_ed
C14_qol <- createC(C2_qol, "te + px_ed")

# Interaction model 15: s-M * px_re (C5_qol) + te * px_ed
C15_qol <- createC(C5_qol, "te + px_ed")

# The following code builds a final model based upon the model fit that will be
# generated for the models above. It can be run later for completeness of the
# example:

# Interaction model 16: c-M * s-M (C13_qol) + s-M * px_re (C13_qol) + te * px_ed
```

```r
C16_qol <- createC(C13_qol, "te + px_ed")
```

```r
cnma_model_fit(cnma_qol, C1_qol, C2_qol, C3_qol, C4_qol, C5_qol, C6_qol,
  C7_qol, C8_qol, C9_qol, C10_qol, C11_qol, C12_qol, C13_qol, C14_qol,
  C15_qol, C16_qol, selection = TRUE)
```

```
## Warning: The following components are not uniquely identifiable: 'C-M', 'C-M * PX_ED', 'C-M * S-M',
## 'C-M * S-M * PX_ED'
```

```
##                  model Q.diff df.diff pval.diff
##   Interaction model 1   1.09       1     0.297
##   Interaction model 2   2.37       1     0.124
##   Interaction model 3   0.25       1     0.614
##   Interaction model 4   1.86       1     0.173
##   Interaction model 5   2.43       1     0.119
##   Interaction model 6   0.54       1     0.464
##   Interaction model 7   3.92       1     0.048
##   Interaction model 8   3.61       3     0.307
##   Interaction model 9   2.58       2     0.275
##  Interaction model 10   3.53       3     0.317
##  Interaction model 11   2.79       3     0.426
##  Interaction model 12   1.17       2     0.558
##  Interaction model 13   3.48       2     0.175
##  Interaction model 14   5.39       2     0.067
##  Interaction model 15   4.33       2     0.115
##  Interaction model 16   5.45       3     0.142
```

##### Compare model fit with previous nested interaction model

We also must compare model fit with each multi-term interaction model's previous nested (single-interaction) model with the lowest Q statistic. First, we will identify the previous nested model with the lowest Q statistic, then generate model fit statistics for the models 13 to 16:

```r
multi.models <- 13:16
#
cnma_model_selection(cnma_qol, C1_qol, C2_qol, C3_qol, C4_qol, C5_qol, C6_qol,
  C7_qol, C8_qol, C9_qol, C10_qol, C11_qol, C12_qol, C13_qol, C14_qol,
  C15_qol, C16_qol, subset = multi.models)
```

```
## Warning: The following components are not uniquely identifiable: 'C-M', 'C-M * PX_ED', 'C-M * S-M',
## 'C-M * S-M * PX_ED'
```

```
##                  model        nested model Q.diff df  pval
##  Interaction model 13 Interaction model 5   1.05  1 0.305
##  Interaction model 14 Interaction model 7   1.48  1 0.224
##  Interaction model 15 Interaction model 7   0.42  1 0.519
##  Interaction model 16 Interaction model 7   1.54  2 0.464
```

### 4. Further CNMA visualizations

Assessment of the additivity assumption can also be aided by visualization of the leave-one-component-out plots using *loccos()* from R package **viscomp**. These plots depict the set of interventions that differ by one component, comparing the NMA relative effect of the intervention that includes the underlying component vs the intervention effect that does not include the component. Points close to the diagonal indicate that the additivity assumption is possible in these effects.

```
DropComps <- "COG"
#
loccos.comps <- cnma_preh$comps[!(cnma_preh$comps %in% DropComps)]
plt <- vector("list", length(loccos.comps))
for (i in seq_along(loccos.comps))
  plt[[i]] <- loccos(nma_preh, combination = loccos.comps[i])
#
grid.arrange(grobs = plt, ncol = 4)
```

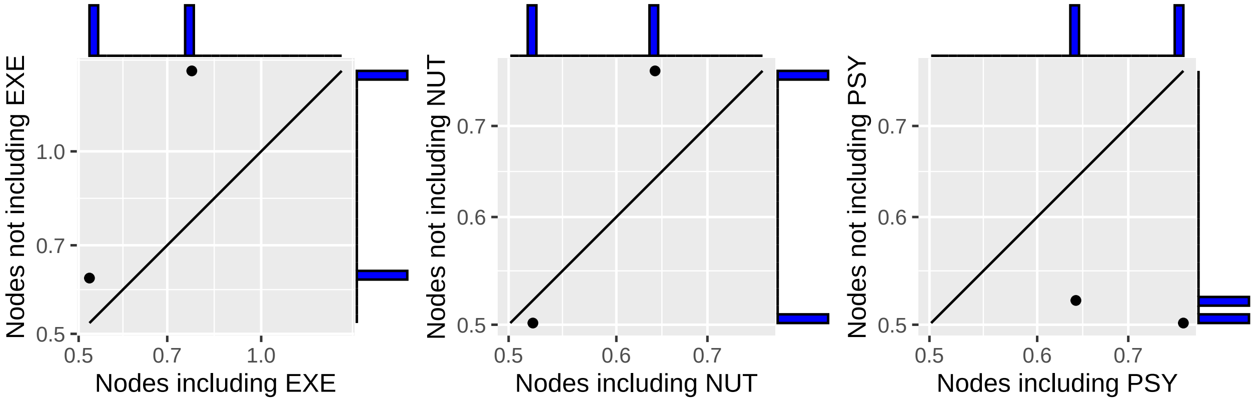


A component heat map for the network can be generated with *heatcomp()* from R package **viscomp**. This plot presents the effects of pairwise component combinations as a median treatment effect (e.g., SMD) of all the combinations that they were part of the NMA model.

```
heatcomp(nma_preh)
```

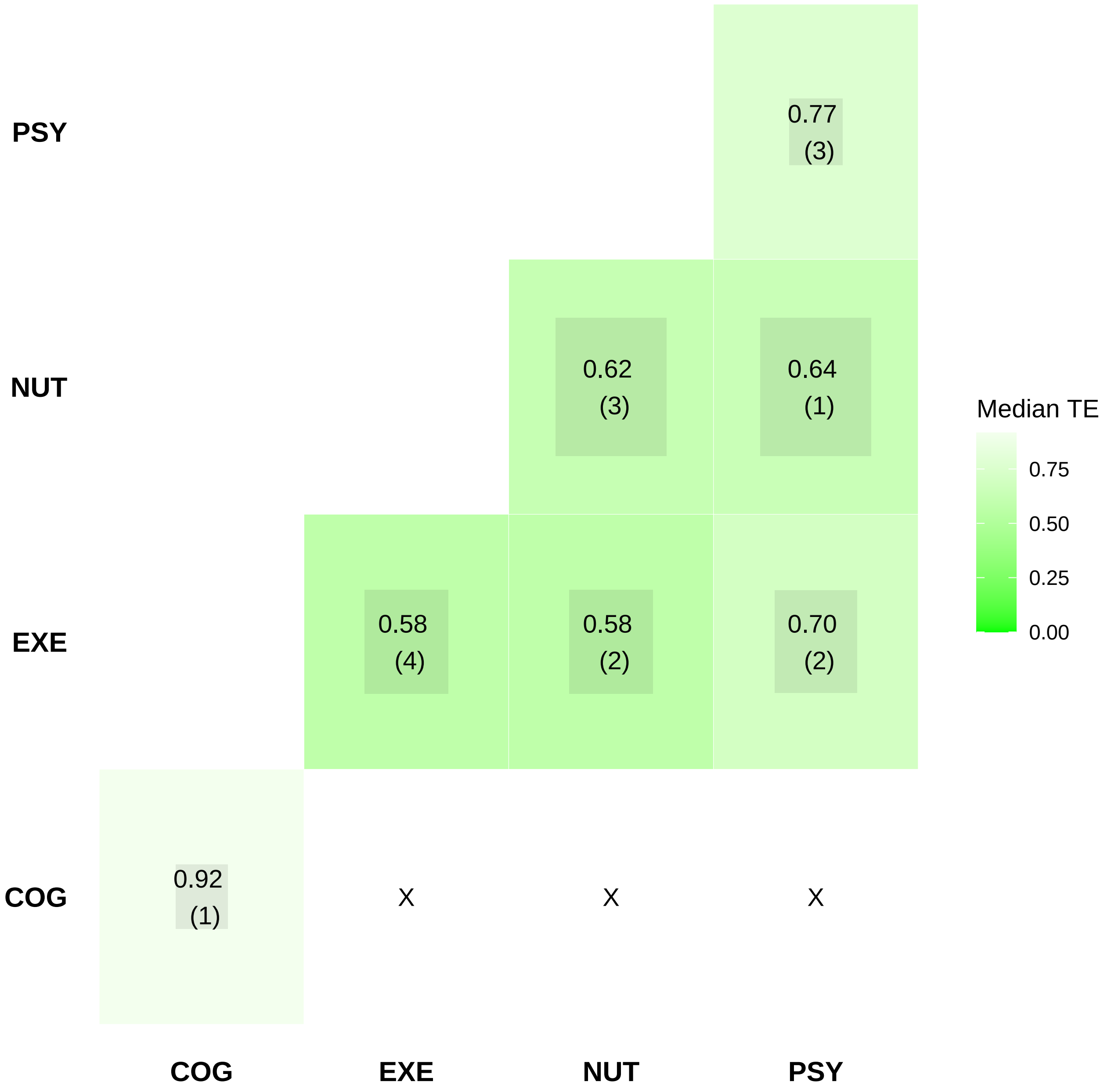


Furthermore, violin plots of the median component-level effects across all treatments containing the component can be generated with *specc()* from R package **viscomp**. Components found in only one treatment will not be plotted:

```
specc(nma_preh) +
  theme(axis.text.x = element_text(size = 13),
        axis.text =  element_text(size = 15),
        axis.title =  element_text(size = 15))
```

```
## Warning: Violin plot requires at least 2 data point
```

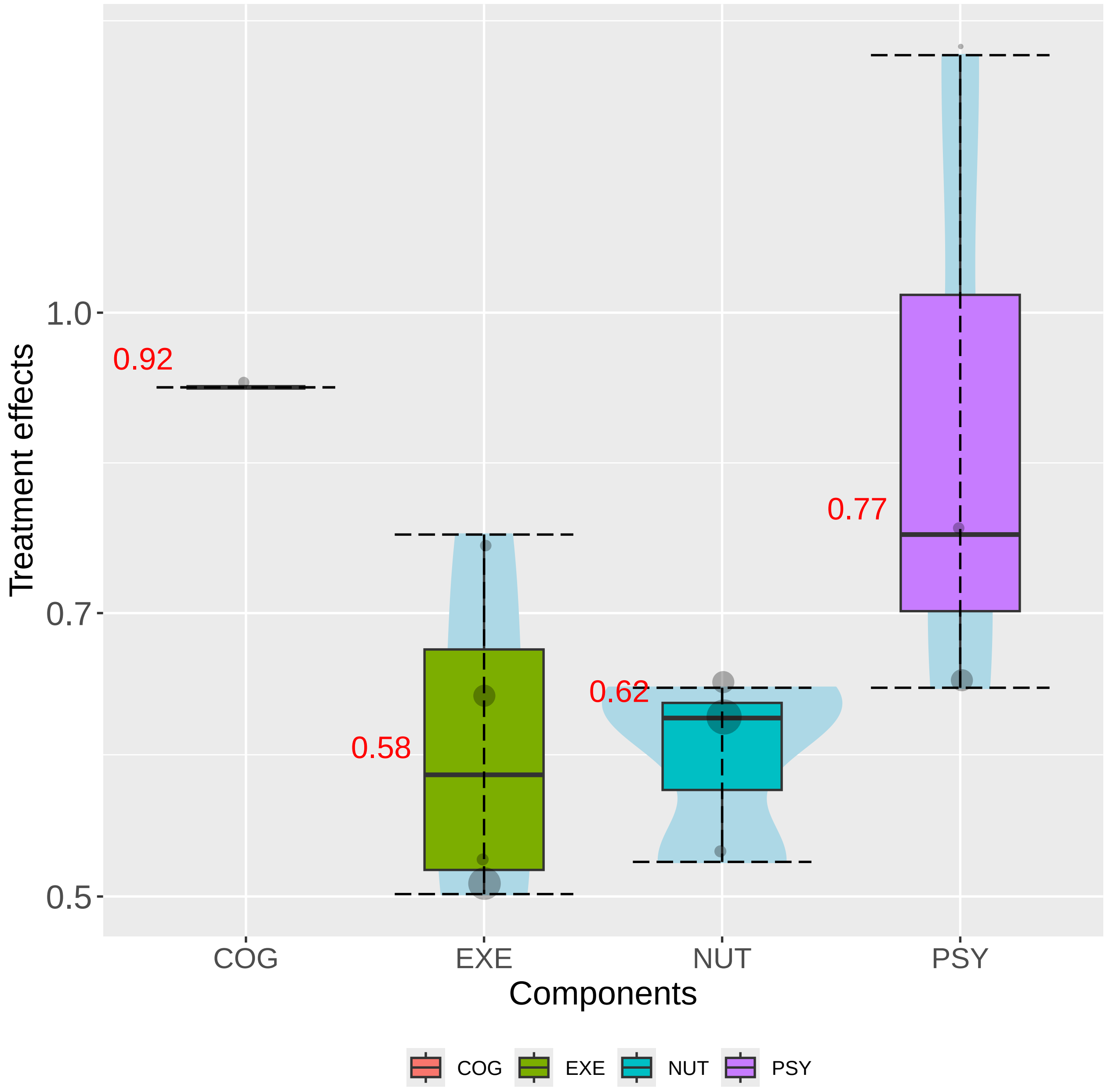

Treatment effects
1.0
0.7
0.5
0.92
0.58
0.62
0.77
COG
EXE
NUT
PSY
Components
COG
EXE
NUT
PSY

### 5. Detailed results for interaction CNMA models

Here, we provide commands to get details on all interactions models for the QoL outcome. We do not show the actual results which span over about 20 pages.

First, we fit all interaction CNMA models.

```
C.interactions <- list(C1_qol, C2_qol, C3_qol, C4_qol, C5_qol, C6_qol,
  C7_qol, C8_qol, C9_qol, C10_qol, C11_qol, C12_qol, C13_qol, C14_qol,
  C15_qol, C16_qol)

# Create an object to house the CNMA model results
net.interactions <- vector("list", length(C.interactions))

# Loop through each multi-term interaction and for each one run the
# interaction CNMA and calculate model fit statistics

for (i in seq_along(C.interactions)) {
  net.interactions[[i]] <-
    netcomb(nma_qol, C.matrix = C.interactions[[i]],
            inactive = nma_qol$reference.group,
            sep.comps = cnma_qol$sep.comps)
  }
#
names(net.interactions) <- names(C.interactions)
```

The following command applies the *netcomplex()* to all interaction models.

```
lapply(net.interactions, netcomplex)
```

The P-scores for each multi-term interaction model can also be calculated.

```
lapply(net.interactions, netrank, small.values = SmallVal)
```
