## Supplementary material for "Analysing complex interventions using component network meta-analysis": R code: funcs.R

```
set <- function(sheet, sm, small.values,
                reference.group, drop.comps,
                left.int, left.comp, right.int, right.comp,
                binary,
                name = sheet) {
  res <- vector("list")
  #
  res$sheet <- sheet
  res$Title <- paste0("Component and Standard NMA\n(", name, ")")
  res$TitleNNA <- paste0("Standard NMA\n(", name, ")")
  res$TitleCNMA <- paste0("CNMA: Treatment-level effects\n(", name, ")")
  res$TitleCompCNMA <- paste0("CNMA: Component-level effects\n(", name, ")")
  #
  res$EffectSize <- sm
  res$SmallVal <- small.values
  res$Reference <- reference.group
  res$DropComps <- drop.comps
  #
  LeftLabelInt <- paste("Favours", left.int)
  LeftLabelComp <- paste("Favours", left.comp)
  RightLabelInt <- paste("Favours", right.int)
  RightLabelComp <- paste("Favours", right.comp)
  res$binary <- binary
  #
  res
}


cnma_model_fit <- function(cnma, ..., n, selection = FALSE) {
  
  C.list <- list(...)
  missing.n <- missing(n)
  #
  if (!missing.n) {
    #
    # Warning if an additional C matrix is provided
    #
    if (length(C.list) > 0)
      warning("Input for argument '...' ignored as argument 'n' is provided.",
              call. = FALSE)
    #
    # Loop through available combinations to create a C matrix with
    # interaction term
    #
    combs <- combinations(cnma, n)
    n.combs <- length(combs)
    C.matrix <- vector("list", length = n.combs)
    #
    for (i in seq_len(n.combs)) {
      C.matrix[[i]] <- createC(cnma, combs[i])
    }
  }
  else {
    C.matrix <- C.list
  }
  #
  if (length(C.matrix) == 0) {
    if (!missing.n)
      warning("No combination available to construct C matrix.",
              call. = FALSE)
    else
      warning("No C matrix provided.", call. = FALSE)
    #
    return(NULL)
  }
  else
    n.mat <- length(C.matrix)
  
  
  # Create a data frame to house model fit statistics
  #
  res <-
    data.frame(model = rep("", n.mat),
               interaction = "",
               n.ia = NA, max.way = NA,
               Q = NA, df = NA, pval = NA,
               tau2 = NA, I2 = NA,
               Q.nma = NA, df.nma = NA, pval.nma = NA,
               Q.add = NA, df.add = NA, pval.add = NA)
  
  for (i in seq_len(n.mat)) {
    #
    # Fit the interaction CNMA model with the new C matrix
    #
    cnet.i <- netcomb(cnma$x, C.matrix = C.matrix[[i]],
                      inactive = cnma$inactive,
                      sep.comps = cnma$sep.comps)
    #
    res$model[i] <- paste("Interaction model", i)
    #
    # Put the interaction name in the data frame
    #
    cn.i <- colnames(C.matrix[[i]])
    cn.i.ia <- cn.i[!(cn.i %in% cnma$comps)]
    #
    res$interaction[i] <- paste(cn.i.ia, collapse = " + ")
    #
    res$n.ia[i] <- length(cn.i.ia)
    res$max.way[i] <- 1 + max(str_count(cn.i.ia, fixed(cnma$sep.ia)))
    #
    # Calculate the difference in Q stats with the additive model
    # (i.e., additive - interaction)
    #
    Q.i <- cnma$Q.additive - cnet.i$Q.additive
    # Calculate the difference in degrees of freedom with the additive model
    df.Q.i <- cnma$df.Q.additive - cnet.i$df.Q.additive
    # Calculate the p-value for the difference in Q stats with the additive model
    pval.Q.i <- pchisq(Q.i, df.Q.i, lower.tail = FALSE)
    #
    # Calculate the difference in Q stats with the standard NMA model
    # (i.e., interaction - nma)
    #
    Q.nma.i <- cnet.i$Q.additive - cnma$Q.standard
    #
    # Calculate the difference in degrees of freedom with the standard NMA model
    #
    df.Q.nma.i <- cnet.i$df.Q.additive - cnma$df.Q.standard
    #
    # Calcuate the p-value for the difference in Q stats with the
    # standard NMA model
    #
    pval.Q.nma.i <- pchisq(Q.nma.i, df.Q.nma.i, lower.tail = FALSE)
    #
    # If the degrees of freedom of the interaction model is less than that of the
    # additive model, add the model fit data to the data frame
    #
    if (cnet.i$df.Q.additive < cnma$df.Q.additive) {
      res[i, c("Q", "df", "pval")] <-
        c(cnet.i$Q.additive, cnet.i$df.Q.additive, cnet.i$pval.Q.additive)
      res[i, c("Q.add", "df.add", "pval.add")] <-
        c(Q.i, df.Q.i, pval.Q.i)
      res[i, c("Q.nma", "df.nma", "pval.nma")] <-
        c(Q.nma.i, df.Q.nma.i, pval.Q.nma.i)
      #
      res$tau2[i] <- round(cnet.i$tau^2, 3)
      res$I2[i] <- round(cnet.i$I2, 3)
    }
  }
  #
  res <- subset(res, !(is.na(Q) & is.na(df) & is.na(pval)))
  #
  attr(res, "selection") <- selection
  #
  class(res) <- c("cnma_model_fit", class(res))
  #
  res
}

print.cnma_model_fit <- function(x,
                                 digits.Q = gs("digits.Q"),
                                 digits.tau2 =  gs("digits.tau2"),
                                 digits.I2 =  gs("digits.I2"),
                                 digits.pval = gs("digits.pval.Q"),
                                 selection = attr(x, "selection"),
                                 ...) {
  formatN <- meta:::formatN
  formatPT <- meta:::formatPT
  #
  x$n.ia <- x$max.way <- NULL
  #
  x$Q <- formatPT(x$Q, digits = digits.Q)
  x$pval <- formatPT(x$pval, digits = digits.pval)
  #
  x$tau2 <- formatPT(x$tau2, digits = digits.tau2)
  x$I2 <- paste0(formatN(100 * x$I2, digits = digits.I2), "%")
  #
  x$Q.nma <- formatPT(x$Q.nma, digits = digits.Q)
  x$pval.nma <- formatPT(x$pval.nma, digits = digits.pval)
  #
  x$Q.add <- formatPT(x$Q.add, digits = digits.Q)
  x$pval.add <- formatPT(x$pval.add, digits = digits.pval)
  #
  if (selection) {
    x$interaction <- x$Q <- x$df <- x$pval <- x$tau2 <- x$I2 <-
      x$Q.nma <- x$df.nma <- x$pval.nma <- NULL
    #
    nam <- names(x)
    nam <- ifelse(nam == "Q.add", "Q.diff", nam)
    nam <- ifelse(nam == "df.add", "df.diff", nam)
    nam <- ifelse(nam == "pval.add", "pval.diff", nam)
    names(x) <- nam
  }
  else {
    x$model <- NULL
  }
  #
  mat <- as.matrix(x)
  #
  rownames(mat) <- rep("", nrow(mat))
  #
  prmatrix(mat, quote = FALSE, right = TRUE)
  #
  invisible(NULL)
}

colnames_ia <- function(x) {
  cn <- colnames(x)
  #
  cn[grepl("*", cn, fixed = TRUE)]
}

cnma_model_selection <- function(cnma, ..., subset = NULL, details = FALSE) {
  
  C.matrix <- list(...)
  #
  if (length(C.matrix) == 0)
      stop("No C matrix provided.", call. = FALSE)
  
  mf <- cnma_model_fit(cnma, ..., selection = TRUE)
  class(mf) <- "data.frame"
  #
  sel.multi <- which(mf$n.ia > 1)
  #
  if (!is.null(subset))
    sel.multi <- sel.multi[sel.multi %in% subset]
  #
  mf.multi <- mf[sel.multi, ]
  mf.single <- mf[-sel.multi, ]
  #
  res <- NULL
  #
  r <- 0
  for (i in sel.multi) {
    r <- r + 1
    mf.i <- mf[i, ]
    mf.oth.i <- mf[-i &
                     ((mf$n.ia < mf.i$n.ia) |
                        (mf$n.ia == mf.i$n.ia & mf$max.way < mf.i$max.way)), ]
    #
    if (details) 
      cat(paste0("*** ", mf.i$model, " ***\n\n",
                 "Interactions: ", mf.i$interaction, "\n\n"))
    #
    ia.i <- unlist(netmeta:::compsplit(mf.i$interaction, cnma$sep.comps))
    mf.min.i <- subset(mf.oth.i, interaction %in% ia.i) %>%
      slice_min(Q, n = 1, with_ties = FALSE)
    #
    Q.diff <- mf.min.i$Q - mf.i$Q
    df <- mf.min.i$df - mf.i$df
    pval <- pchisq(Q.diff, df, lower.tail = FALSE)
    #
    res <- rbind(res,
                 data.frame(model = mf.i$model,
                            comparator = mf.min.i$model,
                            Q.diff, df, pval))
    #
    if (details) {
      cat("Nested model with lowest Q:\n")
      #
      print(mf.min.i %>% select(model, interaction, Q:pval) %>%
              column_to_rownames("model"))
      #
      cat("\nComparison:\n")
      #
      Q.diff <- meta:::formatN(Q.diff, digits = gs("digits.Q"))
      pval <- meta:::formatPT(pval, digits = gs("digits.pval.Q"))
      #
      res.i <- data.frame(Q.diff, df, pval) %>% rename("p-value" = pval)
      rownames(res.i) <- ""
      print(res.i)
      #
      if (r < length(sel.multi))
        cat("\n")
    }
  }
  #
  class(res) <- c("cnma_model_selection", class(res))
  #
  res
}

print.cnma_model_selection <- function(x,
                                       digits.Q = gs("digits.Q"),
                                       digits.pval = gs("digits.pval.Q"),
                                       ...) {
  
  x$Q.diff <- meta:::formatN(x$Q.diff, digits = digits.Q)
  x$pval <- meta:::formatPT(x$pval, digits = digits.pval)
  #
  mat <- as.matrix(x)
  #
  colnames(mat)[colnames(mat) == "comparator"] <- "nested model"
  rownames(mat) <- rep("", nrow(mat))
  #
  prmatrix(mat, quote = FALSE, right = TRUE)
  #
  invisible(NULL)
}
```
